# Safety and Efficacy of Disulfiram in Hospitalized Patients with Moderate COVID-19: A Randomized, Double-Blind, Placebo-Controlled Trial

**DOI:** 10.1101/2023.10.25.23297554

**Authors:** Augusto Cesar Mota, Valdir C. Sant’Ana Filho, Carolyn M Hendrickson, Rachel M. DeVay, Matt Donne, An M. Nguyen, Caroline Junqueira, Mark Marino, Munish Mehra, Ransi Somaratne, Christian Elabd, Ben Kamens, Wendy Cousin

## Abstract

**OBJECTIVES:** Disulfiram, a low-cost generic drug used for alcohol dependence, holds the potential to mitigate disease progression in patients with moderate COVID-19 by targeting inflammasomes. This study aimed to evaluate the clinical efficacy and safety of disulfiram when administered alongside standard of care for the treatment of hospitalized individuals with moderate COVID-19.

**DESIGN:** A randomized, double-blind, placebo-controlled trial.

**SETTING:** Conducted at four clinical sites in Brazil between December 2020 and August 2021.

**PARTICIPANTS:** 140 participants aged 35 and older with laboratory-confirmed SARS-CoV-2 infection, hospitalized for ≤5 days with moderate symptoms of COVID-19 were enrolled, 137 were randomized.

**INTERVENTION:** Participants were randomized in a 1:1 ratio to receive a daily dose of 500 mg of disulfiram (N=68) or placebo (N=69) for 14 days while receiving the current standard of care. Randomization was stratified by age and comorbidities (hypertension, diabetes, and BMI ≥35).

**MEASUREMENTS AND MAIN RESULTS:** The primary outcome, median time to clinical improvement [95% CI] did not significantly differ between groups (disulfiram: 3.5 [3.00, 4.00] days; placebo: 4 [3.00, 5.00] days; P=.73).

Key secondary outcomes, such as mean days (SD) on supplemental oxygen [disulfiram: 4.4 (6.61) days; placebo: 3.7 (5.80) days, P=.34], median (95% CI) time to hospital discharge [disulfiram: 6.0 (5.00, 8.00) days, placebo: 5.0 (4.00, 7.00)], proportion of participants discharged by day 8 [disulfiram (68%), placebo (63%), odds ratio: 0.801], and proportion of participants who clinically worsened [disulfiram (21%), placebo (19%), P=.79], did not reveal significant differences. While the incidence of adverse events was higher in the disulfiram group, serious adverse events and 28-day mortality were comparable between the two groups.

**Conclusions:** Although disulfiram was found to be safe in hospitalized patients with moderate COVID-19, it did not shorten the time to clinical improvement. These findings do not support the use of disulfiram alongside standard of care in this patient population.

**TRIAL REGISTRATION:** ClinicalTrials.gov Identifier: <u>NCT04594343</u>

**Key Points:** *Background:* Disulfiram has been proposed to mitigate disease progression in patients with COVID-19 by targeting the inflammasomes.

*Question:* Does disulfiram, a generic drug used for alcohol use disorder, reduce the time to clinical improvement or reduce the risk of severe disease in hospitalized patients with moderate COVID-19 and with comorbidities when added to the standard of care?

*Findings:* In this double-blind, placebo-controlled randomized clinical trial of adults hospitalized with moderate COVID-19 in Brazil, the addition of an oral disulfiram treatment to the standard of care was safe. Still, it did not decrease the time to clinical improvement.

*Meaning:* The study findings do not support the use of disulfiram in hospitalized patients with moderate COVID-19 in addition to the standard of care.

## Introduction

COVID-19 is a serious respiratory disease caused by the SARS-CoV-2 coronavirus that has caused substantial morbidity and mortality worldwide [1]. No treatment was available at study initiation and although vaccines and recently approved treatments have mitigated the risk of severe COVID-19, affordable and safe therapies remain needed [2,3]. This clinical trial aimed to evaluate disulfiram for the treatment of adult participants hospitalized with COVID-19.

Excessive innate immune response and inflammasome activation have been hypothesized as contributors to the systemic inflammation observed in COVID-19 patients [4]. COVID-19 can lead to acute respiratory distress syndrome (ARDS), multi-organ failure, and death due to an overwhelming innate immune response. Inflammatory cell death pathways, such as pyroptosis and neutrophil extracellular traps formation (NETosis), have been associated with hyperinflammation and tissue damage in COVID-19 patients [5–14]. Inflammasome-mediated pyroptosis is characterized by the formation of Gasdermin D (GSDMD) pores, releasing cytosolic contents such as LDH, HMGB1 and ATP, and proinflammatory cytokines including IL-1β and IL-18 into the extracellular space. Several studies have linked the activation of NLRP3 inflammasomes by rapid innate immune activation after SARS-CoV-2 infection [14–17]. NETosis can contain infections but also cause inflammation and microvascular thrombosis if dysregulated [6,18,19]. NETosis is NLRP3 and GSDMD-mediated[20] and biomarkers have been identified in severe COVID-19 patients, including in their lungs [18,21–24]. Targeting GSDMD, the common downstream effector of inflammasome-mediated pyroptosis and NETosis, could attenuate the excessive inflammatory response and improve SARS-CoV-2 infection outcomes.

Disulfiram, an FDA-approved drug used for alcohol dependence has shown inhibitory effects on GSDMD pore formation, pyroptosis, and NETosis both *in vitro* and *in vivo* [19,25–27]. Disulfiram inhibited NETosis in mouse and human neutrophils, as well as in animal models including the transfusion-related acute lung injury (TRALI) mouse model and SARS-CoV-2 infected golden hamsters. In hamsters, disulfiram also decreased lung inflammation, neutrophil infiltration, and perivascular fibrosis[26]. Observational studies have suggested potential benefits of disulfiram in reducing SARS-CoV-2 infection risk and disease severity in individuals with alcohol use disorder [28,29].

Given these findings, disulfiram affordability and established safety profile, a clinical trial was warranted to evaluate its potential as a COVID-19 treatment. This placebo-controlled study aimed to evaluate the safety and efficacy of disulfiram administration, in addition to standard of care, in hospitalized participants with moderate COVID-19.

## Methods

### Trial Design

This is a multicentered (4 sites), randomized, double-blind, placebo-controlled clinical trial conducted in Brazil (Salvador and Feira de Santana). Eligible participants, upon signing the informed consent form (ICF) were randomized (1:1) to receive disulfiram or placebo. Medication was administered orally once daily for 14 days, with two main visits on days 8 and 15, and a follow-up evaluation on day 28. The trial protocol and statistical analysis plan (**SAP**) are available in **Appendix 1** and **Appendix 2**.

### Ethics

The study received approval from national and local ethics committees (**details in Supplemental Methods**). It adhered to ICH E6 (R2) Good Clinical Practice and followed the Declaration of Helsinki of 1975, ensuring ethical and regulatory compliance. All participants signed an informed consent form (ICF). The trial also followed CONSORT guidelines and was registered on ClinicalTrials.gov (NCT04594343).

### Safety and Oversight

The Data and Safety Monitoring Board (DSMB) reviewed safety data after every 50 participants, as per their charter. Adverse events (AEs) were reported per protocol-defined procedures, and the severity of AEs and serious adverse events (SAEs) was assessed using the Division of AIDS Table for Grading the Severity of Adult and Pediatric Adverse Events, version 2.1.

### Study Population

The study enrolled moderate COVID-19 patients at risk of severe disease. Age was a major risk factor at the beginning of the pandemic[30]. Both genders aged ≥60, with laboratory-confirmed SARS-CoV-2 infection within seven days and hospitalized for up to five days were recruited. Participants aged 50-59 were eligible if they had at least one comorbidity: hypertension, diabetes, BMI ≥35. Moderate disease was defined as hospitalization without high-flow oxygen or mechanical ventilation, respiratory rate ≤30 per minute, and no intensive care unit (ICU)-level care (score 3 or 4 on a WHO 7-point ordinal scale, **Table S1**). Female participants of childbearing potential needed a negative hCG pregnancy test. Exclusion criteria included allergy to disulfiram, active hepatitis, elevated ALT or AST levels, malignancy, and severe chronic kidney disease. Participants were required to abstain from alcoholic substances within 24 hours prior to treatment and until 14 days after the last dose of study medication. With the emergence of new variants and beginning of vaccination, we amended the protocol to include participants aged 35 and older and only included moderate cases of COVID-19 requiring supplemental oxygen as explained in **Supplemental Methods**.

### Randomization, Stratification, and Masking

To minimize the impact of age and comorbidities on the treatment outcomes, randomization was stratified based on four different risk strata described in **Supplemental Methods and in Table S2**.

A random allocation sequence was generated by a statistician analyst using a SAS® Version 9.4 program, employing random permuted block sizes of 4 within each stratum. To ensure the study’s integrity, group assignments were kept blinded from patients, treating clinicians, trial personnel, and outcome assessors. More details on the masking process and randomization implementation can be found in **Supplemental Methods**.

### Data Collection and Monitoring

Clinical research data from source documentation including, but not limited to, WHO score, AEs, SAEs, concomitant medications, medical history, clinical laboratory data, and others were entered into a validated EDC system compliant with US FDA 21 CFR Part 11. AEs were coded using the MedDRA Dictionary (version 23.1), while concomitant medications were coded according to the WHO Drug Global Dictionary (version September 1, 2020, B3). Medical history was not coded or tabulated. Statistical programming to generate tables and listings was performed using SAS^®^ Version 9.4 by Tigermed.

### Intervention

Participants received capsules containing 500 mg of disulfiram or a matching placebo indistinguishable from the study drug. A compounding pharmacy prepared the capsules using 400 mg Antabuse tablets (Sanofi®, Belgium), since disulfiram was approved but not commercially available in Brazil. Those who experienced difficulty swallowing capsules were given the option to take the medication in syrup form, orally or via nasogastric tube. The treatment was administered by a healthcare professional to hospitalized participants. After discharge, participants received capsules to take home. Drug accountability was performed at day 8 and day 15 visits.

### Procedures

At baseline, participant data, including medical history, demographics, medications, vitals, and lab tests were documented, including SARS-CoV-2 confirmation. Baseline assessments included ECG, physical exam and chest CT. Throughout the study, vital signs, lab tests, WHO scores, and medication details were collected periodically. Hospitalized participants underwent daily monitoring, while discharged ones finished treatment at home, returning for assessments on days 8 and 15. A follow-up took place on day 28. Detailed procedures are available in **Supplemental Methods** and in the trial protocol **(Appendix 1)**.

### Outcome Measures

The primary endpoint was time to clinical improvement, measured from baseline to the first post-baseline assessment with a ≥1 point increase on the WHO ordinal scale (**Table S1**). Key secondary endpoints included days on supplemental oxygen; time of hospitalization; percentage of participants discharged by day 8; percentage of participants with clinical worsening (≥1 points on the WHO ordinal scale); duration of non-invasive ventilation or invasive mechanical ventilation (WHO score 5 or 6); and 28-day mortality. Endpoints changes made after trial commencement can be found in **Supplemental Methods.**

Subgroup analyses considered factors such as sex, age, baseline risk categories, comorbidities (hypertension, diabetes, BMI>35), baseline hyperinflammation state, clinical status, and parameters associated with increased mortality. Proinflammatory cytokines (interleukin (IL)-1β, IL-6, IL-18, tumor necrosis factor-alpha (TNF-α)) and cell-free DNA (cf-DNA), were quantified from plasma at baseline, day 8 and day 15 using multiplex Luminex assay (Thermo Fisher), and PicoGreen-based assay (Thermo Fisher).

### Sample Size Calculation

As there was no pre-existing data on the efficacy of disulfiram in COVID-19 patients or in pre-clinical studies, the sample size for this study was not determined through formal calculations. A sample size of 200 participants was considered adequate to assess the efficacy of disulfiram. Due to a decrease in the number of patients with COVID-19, the study enrollment was terminated early after randomizing 140 participants.

### Statistical Analysis

The primary and key secondary efficacy endpoints were tested sequentially to account for multiplicity and preserve the overall type I error at 0.05. No adjustments were made for multiple comparisons in testing other secondary or exploratory efficacy endpoints. The modified intent-to-treat (mITT) population was used for all efficacy analyses. It included all randomized participants who received at least one dose of the study drug and had their WHO score measured at baseline and at least one post-baseline. The **SAP** (**Appendix 1**) was finalized and signed prior to locking the database and unblinding the study. A summary of the statistical analysis in Supplemental Methods.

## Results

### Study Population

From December 2020 to August 2021, 168 signed ICF for eligibility evaluation. Of those, 28 participants were not randomized: 23 did not meet the eligibility criteria, 4 withdrew consent, and 1 was discharged before randomization. The remaining 140 participants underwent randomization to receive disulfiram (n=69) or placebo (n=71). However, 3 participants were excluded from the mITT population as they did not receive the intervention: 2 withdrew consent before taking the first dose (one from each arm), and 1 did not meet the criteria at baseline in the placebo arm.

Among the 137 participants receiving the intervention, 68 received disulfiram and 69 received placebo. Completion rates for day 15 and day 28 visits were high in both groups (day 15: 87.3% placebo and 91.3% disulfiram; day 28: 83.1% placebo and 91.3% disulfiram). Overall, 12 participants discontinued the treatment due to consent withdrawal (2 in disulfiram, 4 in placebo), death (2 in disulfiram, 1 in placebo), and self-reported treatment discontinuation (2 in placebo, 1 in disulfiram).

Between the end of the treatment period and the follow-up visit, 2 participants receiving placebo refused to return for visits, and one died. In the disulfiram group, one participant died during this period but is not listed on the chart because they withdrew consent before day 15. However, they are included in the count of participants who died due to any cause over 28-day period in the mITT population (as mentioned in **Figure 1, footnote d**).

**Figure 1.**
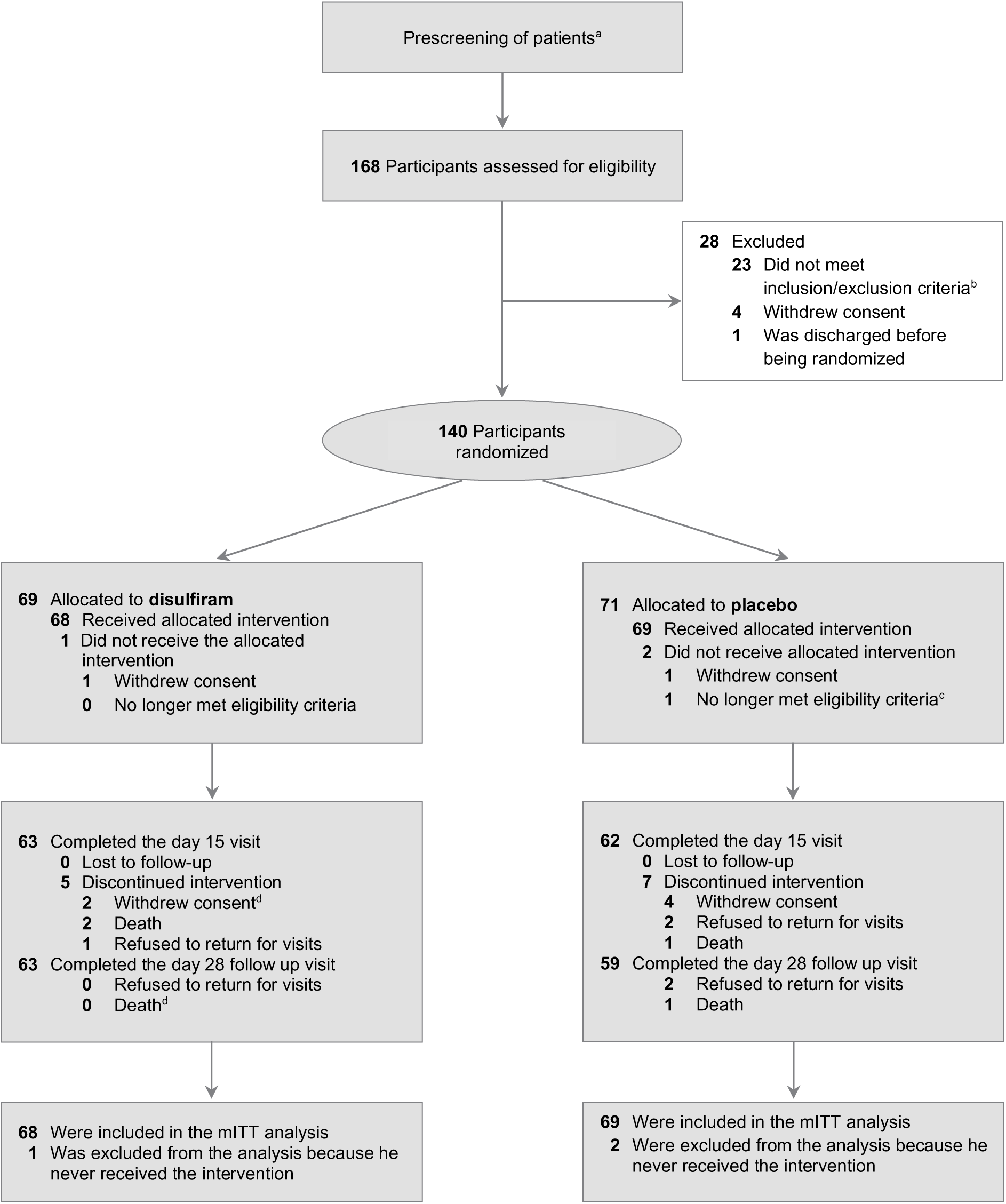
Flow Diagram of Participants. ^a^ Patients potentially eligible were informed by study investigators. The number of informed patients was not collected. ^b^ Out of the 23vwho didn’t meet inclusion/exclusion criteria, 13 tested negative for SARS-CoV-2, 3 had ALT or AST levels more than 3 times the upper limit of normal, 3 had a SpO2 lower than 93% on room air or using supplemental O2 via nasal cannula with a flow rate of up to 3 L/min, 2 needed invasive or non-invasive ventilation at screening or baseline, 2 did not need supplemental O2 via nasal cannula or equivalent (as per protocol amendment 1.3 inclusion/exclusion criteria, patients who didn’t need supplemental oxygen (WHO score of 3) were not eligible), 1 was admitted into the Intensive Care Unit (ICU) before baseline and 1 had a severe kidney dysfunction. ^c^ Negative SARS-CoV-2 PCR at baseline. ^d^ One of the two participants who withdrew consent before day 15 visit died at day 20 and is included in the participant who died due to any cause over 28 days period in the mITT population death at 28 day.

### Baseline Demographics and Characteristics

The demographics and baseline characteristics were generally well-balanced between groups (**Table 1**). Mean age and gender distribution were similar [mean age (SD): 61.6 (10.95) disulfiram, 60.7 (11.15) placebo; female participants: 45.6% disulfiram, 43.5% placebo]. Mean BMI (SD) was 28.5 (5.36) in disulfiram (11.8% BMI >35) and 29.4 (5.94) in placebo (14.5% BMI >35). Hypertension affected 61.8% of participants in the disulfiram group and 59.4% in the placebo group, diabetes affected 35.3% and 40.6%, respectively. The distribution of age and comorbidities is presented in **Table S3** and risk groups were well balanced. An imbalance was observed in race distribution: while most participants (66.2% disulfiram, 62.3% placebo) self-identified as Pardo (mixed race: White/Black; White/Indigenous; Black/Indigenous; White/Black/Indigenous), a higher percentage self-identified as White in placebo, and as Black in disulfiram (White: 5.9% disulfiram, 20.3% placebo; Black: 27.9% disulfiram, 17.4% placebo). Note that race reporting in Brazil differs from North American structures, posing limitations in traditional methods to capture and report race profiles due to a complex mixture of ethnic and racial groups and fluid self-identification. Blood cell count was generally well balanced except for participants with a neutrophil-to-lymphocyte ratio (NLR) ≥6.1, where the percentage was higher in placebo (46.4%) than in disulfiram (30.9%). Prior and concomitant medications usage was also well-balanced (data not shown), although it’s worth noting that 73% of enrolled participants were taking systemic corticosteroids prior to the study (73.5% disulfiram, 72.5% placebo) and 81% received them during the study (77.9% disulfiram, 84.1% placebo).

**Table 1.**
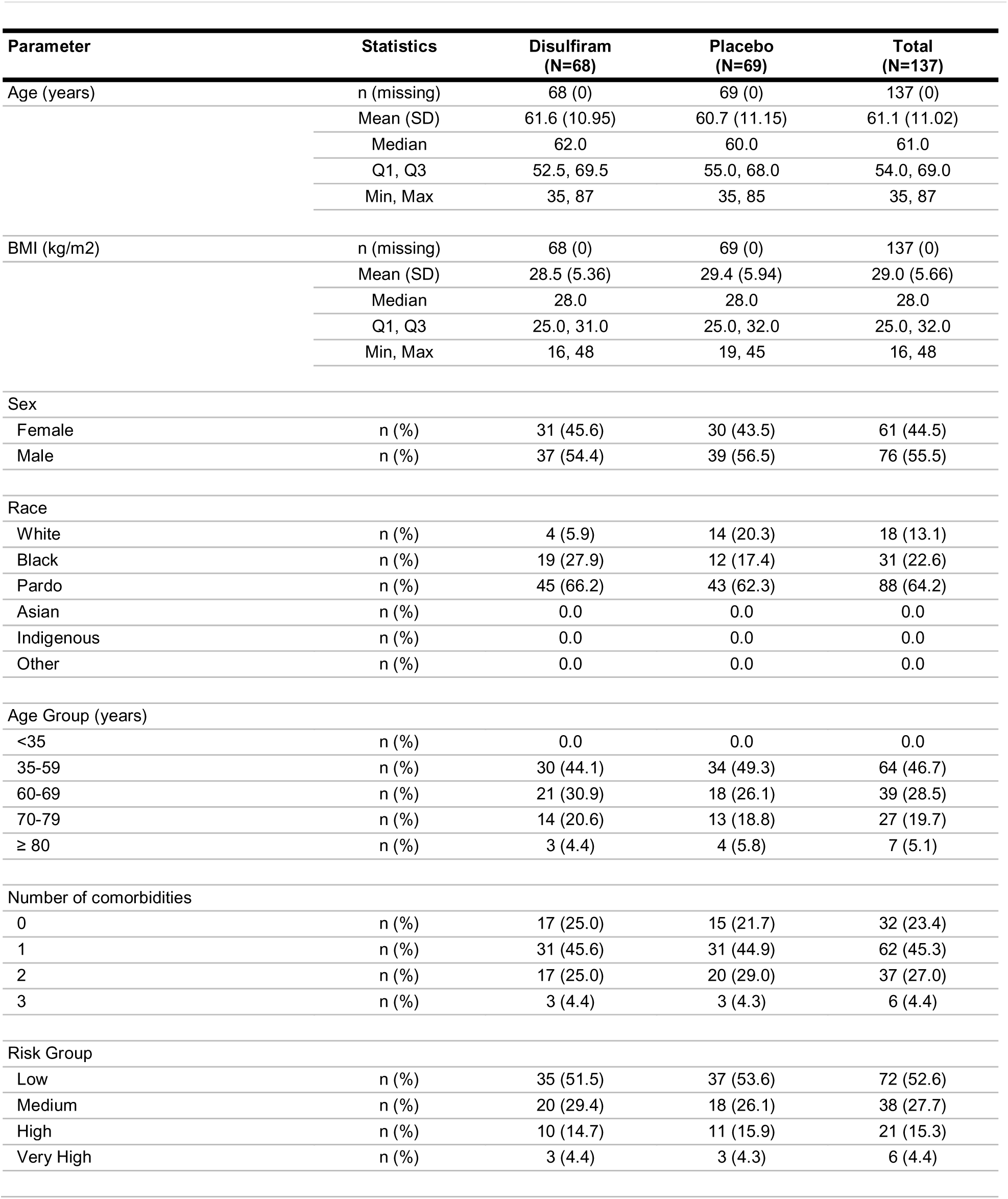
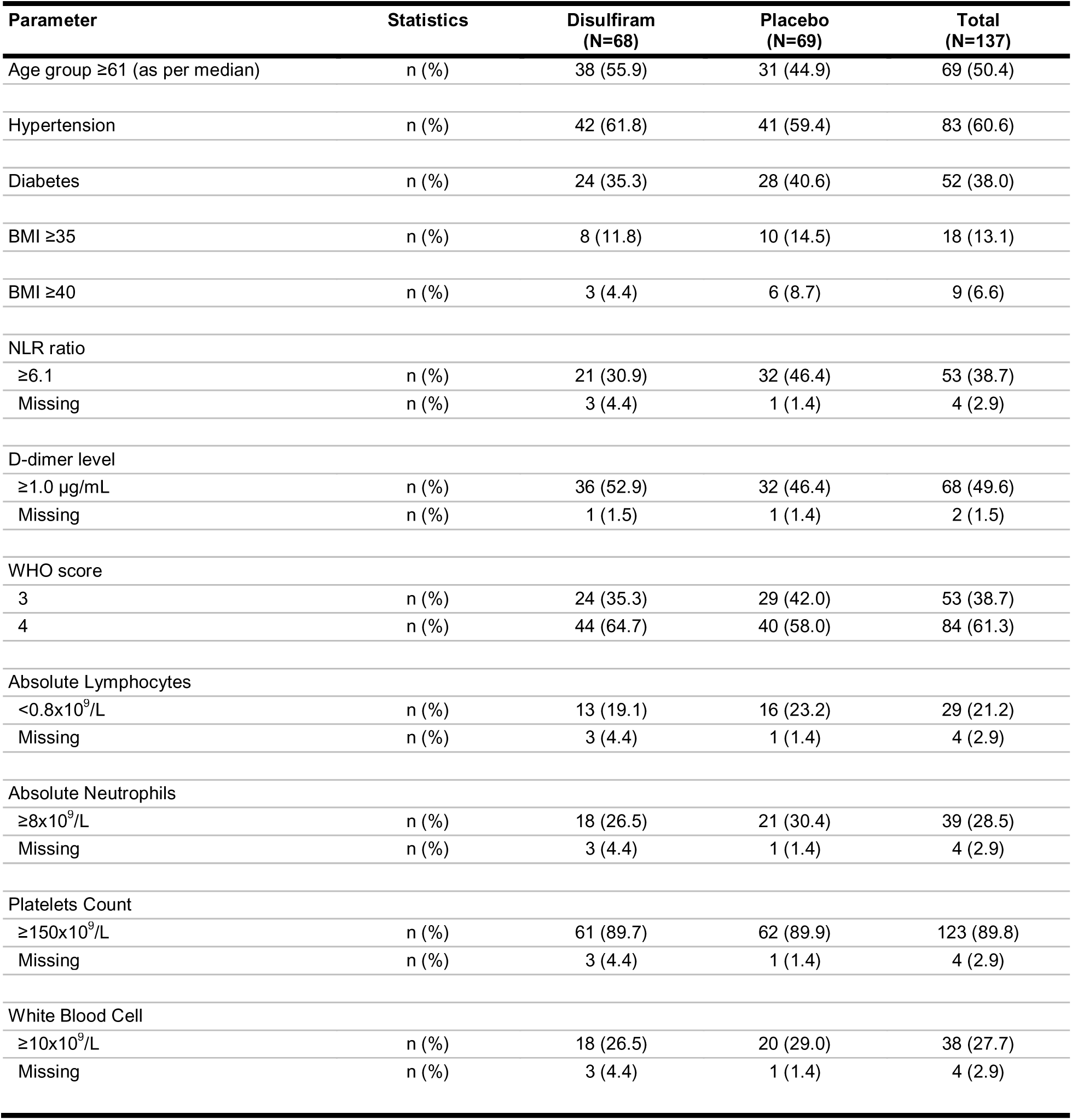
Summary of Demographics and Baseline Characteristics Safety Populations. Abbreviations: BMI: Body Mass Index; cm: centimeter; kg: kilogram; kg/m2: kilogram per square meter; SD: Standard Deviation; Q1: First Quartile (25th Percentile); Q3: Third Quartile (75th Percentile); Min: Minimum; Max: Maximum; L: Liter; pg/mL: picograms per milliliter; µg/mL: micrograms per milliliter; NLR: Absolute Neutrophil to Absolute Lymphocyte Ratio; WHO: World Health Organization. **Notes:**

| Parameter | Statistics | Disulfiram<br>(N=68) | Placebo<br>(N=69) | Total<br>(N=137) |
| --- | --- | --- | --- | --- |
| Age (years) | n (missing) | 68 (0) | 69 (0) | 137 (0) |
|  | Mean (SD) | 61.6 (10.95) | 60.7 (11.15) | 61.1 (11.02) |
|  | Median | 62.0 | 60.0 | 61.0 |
|  | Q1, Q3 | 52.5, 69.5 | 55.0, 68.0 | 54.0, 69.0 |
|  | Min, Max | 35, 87 | 35, 85 | 35, 87 |
| BMI (kg/m2) | n (missing) | 68 (0) | 69 (0) | 137 (0) |
|  | Mean (SD) | 28.5 (5.36) | 29.4 (5.94) | 29.0 (5.66) |
|  | Median | 28.0 | 28.0 | 28.0 |
|  | Q1, Q3 | 25.0, 31.0 | 25.0, 32.0 | 25.0, 32.0 |
|  | Min, Max | 16, 48 | 19, 45 | 16, 48 |
| Sex |  |  |  |  |
| Female | n (%) | 31 (45.6) | 30 (43.5) | 61 (44.5) |
| Male | n (%) | 37 (54.4) | 39 (56.5) | 76 (55.5) |
| Race |  |  |  |  |
| White | n (%) | 4 (5.9) | 14 (20.3) | 18 (13.1) |
| Black | n (%) | 19 (27.9) | 12 (17.4) | 31 (22.6) |
| Pardo | n (%) | 45 (66.2) | 43 (62.3) | 88 (64.2) |
| Asian | n (%) | 0.0 | 0.0 | 0.0 |
| Indigenous | n (%) | 0.0 | 0.0 | 0.0 |
| Other | n (%) | 0.0 | 0.0 | 0.0 |
| Age Group (years) |  |  |  |  |
| <35 | n (%) | 0.0 | 0.0 | 0.0 |
| 35-59 | n (%) | 30 (44.1) | 34 (49.3) | 64 (46.7) |
| 60-69 | n (%) | 21 (30.9) | 18 (26.1) | 39 (28.5) |
| 70-79 | n (%) | 14 (20.6) | 13 (18.8) | 27 (19.7) |
| ≥ 80 | n (%) | 3 (4.4) | 4 (5.8) | 7 (5.1) |
| Number of comorbidities |  |  |  |  |
| 0 | n (%) | 17 (25.0) | 15 (21.7) | 32 (23.4) |
| 1 | n (%) | 31 (45.6) | 31 (44.9) | 62 (45.3) |
| 2 | n (%) | 17 (25.0) | 20 (29.0) | 37 (27.0) |
| 3 | n (%) | 3 (4.4) | 3 (4.3) | 6 (4.4) |
| Risk Group |  |  |  |  |
| Low | n (%) | 35 (51.5) | 37 (53.6) | 72 (52.6) |
| Medium | n (%) | 20 (29.4) | 18 (26.1) | 38 (27.7) |
| High | n (%) | 10 (14.7) | 11 (15.9) | 21 (15.3) |
| Very High | n (%) | 3 (4.4) | 3 (4.3) | 6 (4.4) |
| Age group ≥61 (as per median) | n (%) | 38 (55.9) | 31 (44.9) | 69 (50.4) |
| Hypertension | n (%) | 42 (61.8) | 41 (59.4) | 83 (60.6) |
| Diabetes | n (%) | 24 (35.3) | 28 (40.6) | 52 (38.0) |
| BMI ≥35 | n (%) | 8 (11.8) | 10 (14.5) | 18 (13.1) |
| BMI ≥40 | n (%) | 3 (4.4) | 6 (8.7) | 9 (6.6) |
| NLR ratio |  |  |  |  |
| ≥6.1 | n (%) | 21 (30.9) | 32 (46.4) | 53 (38.7) |
| Missing | n (%) | 3 (4.4) | 1 (1.4) | 4 (2.9) |
| D-dimer level |  |  |  |  |
| ≥1.0 µg/mL | n (%) | 36 (52.9) | 32 (46.4) | 68 (49.6) |
| Missing | n (%) | 1 (1.5) | 1 (1.4) | 2 (1.5) |
| WHO score |  |  |  |  |
| 3 | n (%) | 24 (35.3) | 29 (42.0) | 53 (38.7) |
| 4 | n (%) | 44 (64.7) | 40 (58.0) | 84 (61.3) |
| Absolute Lymphocytes |  |  |  |  |
| <0.8x10<sup>9</sup>/L | n (%) | 13 (19.1) | 16 (23.2) | 29 (21.2) |
| Missing | n (%) | 3 (4.4) | 1 (1.4) | 4 (2.9) |
| Absolute Neutrophils |  |  |  |  |
| ≥8x10<sup>9</sup>/L | n (%) | 18 (26.5) | 21 (30.4) | 39 (28.5) |
| Missing | n (%) | 3 (4.4) | 1 (1.4) | 4 (2.9) |
| Platelets Count |  |  |  |  |
| ≥150x10<sup>9</sup>/L | n (%) | 61 (89.7) | 62 (89.9) | 123 (89.8) |
| Missing | n (%) | 3 (4.4) | 1 (1.4) | 4 (2.9) |
| White Blood Cell |  |  |  |  |
| ≥10x10<sup>9</sup>/L | n (%) | 18 (26.5) | 20 (29.0) | 38 (27.7) |
| Missing | n (%) | 3 (4.4) | 1 (1.4) | 4 (2.9) |

1. Percentages are based on the number of treated subjects per treatment group N.
2. A subject can belong to more than one race category.

### Primary Outcome

In the mITT population of 137 participants, 127 (92.7%) showed clinical improvement: 94.1% in the disulfiram arm and 91.3% in the placebo arm. Time to clinical improvement was similar between the two arms with a median of 3.5 days (95% CI 3.00, 4.00) for disulfiram and 4 days (95% CI 3.00, 5.00) for placebo, log-rank *P*=.73 (**Table 2** and **Figure S1**)

**Table 2.** Primary and key secondary efficacy endpoints over 28 days period, mITT Population. **Abbreviations**: CI: Confidence Interval; Max: Maximum; ICU: Intensive Care Unit; Min: Minimum; mITT: modified Intent-to-Treat; N: Number of subjects in mITT population in SD: Standard Deviation; SE: Standard Error; WHO: World Health Organization; NE: Not Estimable.each treatment group; Q1: First Quartile (25th Percentile); Q3: Third Quartile (75th Percentile); SD: Standard Deviation; SE: Standard Error; WHO: World Health Organization; NE: Not Estimable. **Footnotes:**

| Primary and Key Secondary Endpoints<br>Parameters | Disulfiram<br>(N=68) | Placebo<br>(N=69) |
| --- | --- | --- |
| **Time to clinical improvement (decrease of $\geq 1$ point on the WHO scale) over 28 days period** |  |  |
| Kaplan-Meier estimate of time to event (days) |  |  |
| Number n (%) of subjects with event (clinical improvement) | 64 (94.1) | 63 (91.3) |
| Number n (%) of subjects censored | 4 (5.9) | 6 (8.7) |
| Q1 (95% CI) | 2.0 (2.00, 3.00) | 2.0 (NE, NE) |
| Median (95% CI) | 3.5 (3.00, 4.00) | 4.0 (3.00, 5.00) |
| Q3 (95% CI) | 7.5 (5.00, 12.00) | 7.0 (5.00, 11.00) |
| Log-rank test p-value vs. Placebo <sup>1</sup> | 0.729 |  |
| **Mean number of days of supplemental oxygen (WHO score of 4, 5, 6, 7 or Death)** |  |  |
| n (missing) | 68 (0) | 69 (0) |
| Mean (SD) | 4.4 (6.61) | 3.7 (5.80) |
| Median | 2 | 1 |
| Q1, Q3 | 0.0, 6.0 | 0.0, 5.0 |
| Min, Max | 0, 28 | 0, 28 |
| LS Mean (SE) <sup>2</sup> | 6.45 (1.063) | 5.00 (1.057) |
| 95% CI LS Mean <sup>2</sup> | 4.35, 8.55 | 2.91, 7.10 |
| LS Mean Difference (SE) vs Placebo <sup>2</sup> | 1.45 (1.499) |  |
| 95% CI vs Placebo <sup>2</sup> | -1.52, 4.41 |  |
| p-value vs Placebo <sup>2</sup> | 0.337 |  |
| **Time from baseline to discharge from hospital over 28 days period** |  |  |
| Kaplan-Meier estimate of time to event (days) |  |  |
| Number n (%) of subjects with event (discharge from hospital) | 64 (94.1) | 63 (91.3) |
| Number n (%) of subjects censored | 4 (5.9) | 6 (8.7) |
| Q1 (95% CI) | 4.0 (3.00, 4.00) | 3.0 (2.00, 4.00) |
| Median (95% CI) | 6.0 (5.00, 8.00) | 5.0 (4.00, 7.00) |
| Q3 (95% CI) | 10.5 (8.00, 14.00) | 9.0 (7.00, 12.00) |
| Log-rank test p-value vs. Placebo <sup>1</sup> | 0.286 |  |
| **Percentage of subjects discharged by day 8 Visit** |  |  |
| n (missing) | 68 (0) | 69 (0) |
| Yes n (%) | 43 (63.2) | 47 (68.1) |
| No n (%) | 25 (36.8) | 22 (31.9) |
| Clopper-Pearson (Exact) 95% CI for Yes percentage | 50.7, 74.6 | 55.8, 78.8 |
| Odds Ratio <sup>3</sup> | 0.801 |  |
| 95% CI <sup>3</sup> | 0.388, 1.656 |  |
| p-value $\chi^2$ | 0.55 | |
| **Percentage of subjects that worsened (increase of $\geq 1$ point on the WHO scale) from baseline to anytime post-baseline** |  |  |
| n (missing) | 68 (0) | 69 (0) |
| Yes n (%) | 14 (20.6) | 13 (18.8) |
| No n (%) | 54 (79.4) | 56 (81.2) |
| Clopper-Pearson (Exact) 95% CI for Yes percentage | 11.7, 32.1 | 10.4, 30.1 |
| Odds Ratio <sup>3</sup> | 1.126 |  |
| 95% CI <sup>3</sup> | 0.478, 2.652 |  |
| p-value □ | 0.786 | |
| **Mean number of days of non-invasive ventilation or invasive mechanical ventilation (WHO score of 5, 6, 7 or Death)** |  |  |
| n (missing) | 68 (0) | 69 (0) |
| Mean (SD) | 1.9 (6.12) | 1.6 (5.31) |
| Median | 0 | 0 |
| Q1, Q3 | 0.0, 0.0 | 0.0, 0.0 |
| Min, Max | 0, 28 | 0, 28 |
| LS Mean (SE) <sup>2</sup> | 3.36 (0.994) | 3.18 (0.989) |
| 95% CI LS Mean <sup>2</sup> | 1.40, 5.33 | 1.23, 5.14 |
| LS Mean Difference (SE) vs Placebo <sup>2</sup> | 0.18 (1.402) |  |
| 95% CI vs Placebo <sup>2</sup> | -2.59, 2.96 |  |
| p-value vs Placebo <sup>2</sup> | 0.897 |  |
| **Mean number of days of subjects were in ICU** |  |  |
| n (missing) | 68 (0) | 69 (0) |
| Mean (SD) | 2.3 (6.32) | 1.8 (5.41) |
| Median | 0 | 0 |
| Q1, Q3 | 0.0, 0.0 | 0.0, 0.0 |
| Min, Max | 0, 28 | 0, 28 |
| LS Mean (SE) <sup>2</sup> | 4.31 (1.006) | 3.31 (1.001) |
| 95% CI LS Mean <sup>2</sup> | 2.32, 6.30 | 1.33, 5.29 |
| LS Mean Difference (SE) vs Placebo <sup>2</sup> | 1.00 (1.419) |  |
| 95% CI vs Placebo <sup>2</sup> | -1.81, 3.81 |  |
| p-value vs Placebo <sup>2</sup> | 0.482 |  |
| **Percentage of subjects on non-invasive ventilation or invasive mechanical ventilation (WHO score of 5, 6, 7 or Death)** |  |  |
| n (missing) | 68 (0) | 69 (0) |
| Yes n (%) | 11 (16.2) | 11 (15.9) |
| No n (%) | 57 (83.8) | 58 (84.1) |
| Clopper-Pearson (Exact) 95% CI for Yes percentage | 8.4, 27.1 | 8.2, 26.7 |
| Odds Ratio <sup>3</sup> | 1.031 |  |
| 95% CI <sup>3</sup> | 0.409, 2.599 |  |
| p-value □ | 0.949 | |
| **Percentage of subjects died due to any cause over 28 days period** |  |  |
| n (missing) | 68 (0) | 69 (0) |
| Yes n (%) | 3 (4.4) | 2 (2.9) |
| No n (%) | 65 (95.6) | 67 (97.1) |
| Clopper-Pearson (Exact) 95% CI for Yes percentage | 0.9, 12.4 | 0.4, 10.1 |
| p-value □ | 0.681 | |

1. p-value generated from log-rank test.
2. p-value and LS Mean statistics generated from ANCOVA model.
3. Odds ratio and 95% CI are calculated from the logistic regression model.

- p-value generated from Wald Chi-Square tests from the logistic regression model.
- p-value generated from Chi-Square/Fisher’s Exact test. **Notes:**

1. Baseline is defined as the last non-missing measure prior to treatment.
2. Percentages are based on the total number of subjects in each column with non-missing data at that or across visit.
3. In logistic regression model, the independent variables are treatment group and baseline risk category.
4. In ANCOVA model, mean number of days are the dependent variable, treatment as a fixed effect and adjusted for covariates baseline risk category and the interaction between treatment groups and baseline risk categories.
5. The time to improvement is defined as the time (in days) from baseline to the earliest day of improvement (decrease of at least one point from baseline) at any post baseline WHO assessment.
6. Deaths over 28 days are only included in analysis. For time to event analysis, deaths were censored at 28 days. For analysis of days endpoint, deaths were assigned a score of 28 days.

### Secondary Outcomes

All the analyses presented in **Table 2** were conducted on the mITT population (n=137). No significant differences were found between the two arms in any of the secondary endpoints.

Mean (SD) supplemental oxygen duration was 4.4 (6.61) days for disulfiram and 3.7 (5.80) days for placebo. Least square (LS) mean (SD) was 6.45 (1.063) days for disulfiram and 5.00 (1.057) days for placebo (*P*=.34).

A large proportion of participants (92.7%) were discharged over 28 days and no significant difference was observed in the time to discharge (**Table 2** and **Figure S2**). Median time to discharge was 6 days (95% CI 5.00, 8.00) for disulfiram and 5 days (95% CI 4.00, 7.00) for placebo, *P*=.29. By day 8 visit, 65.7% of participants were discharged (63.2% disulfiram, 68.1% placebo). The odds ratio was 0.801 (95% CI 0.388, 1.656; *P*=.55).

Percentage of participants who clinically worsened (increase ≥1 point on the WHO scale) was 20.6% for disulfiram and 18.8% for placebo. Odds ratio: 1.126 (95% CI .478, 2.653; *P*=.79).

In the mITT population, 22 participants (16%) received non-invasive/invasive ventilation, with 11 participants in each arm. Odds ratio was 1.031 (95% CI 0.409, 2.599; *P*=.95) Duration of non-invasive /invasive ventilation was similar in both arms [mean (SD) days: 1.9 (6.12) disulfiram, 1.6 (5.31) placebo; LS mean (SD): 3.36 (0.994) disulfiram, 3.18 (0.989) placebo, *P*=.90]. No difference was observed in the duration in ICU [mean (SD) days in the ICU: 2.3 (6.32) disulfiram, 1.8 (5.41) placebo; LS mean (SE): 4.31 (1.006) disulfiram, 3.31 (1.001) placebo, *P*=.48).

Regarding 28-day mortality, 3 participants died (4.4%) in disulfiram and 2 (2.9%) in placebo (*P*=.68).

### Subgroup Analysis

Pre-specified subgroup analyses were conducted to explore patterns and determine potential heterogeneity in the treatment effect. The table in **Figure 2** presents the number of participants in each subgroup and per treatment group. Subgroup analyses were performed on the primary endpoint and the number of participants with clinical improvement in each subgroup is noted in the table. The forest plot in **Figure 2** displays the Kaplan-Meier estimate of time to clinical improvement across subgroups. No statistically significant changes were observed, although there was a trend towards longer improvement times in disulfiram for the subgroup with NLR >6.1 (*P*=.06). A similar trend was observed in the absolute lymphocyte subgroups but was not statistically significant (*P*=.27). Importantly, most participants with severe lymphopenia at randomization were in the disulfiram group (7 out of 8). Overall, subgroup analyses on primary (**Figure 2**) and secondary endpoints (data not shown) revealed very few differences and no identifiable trends.

**Figure 2.**
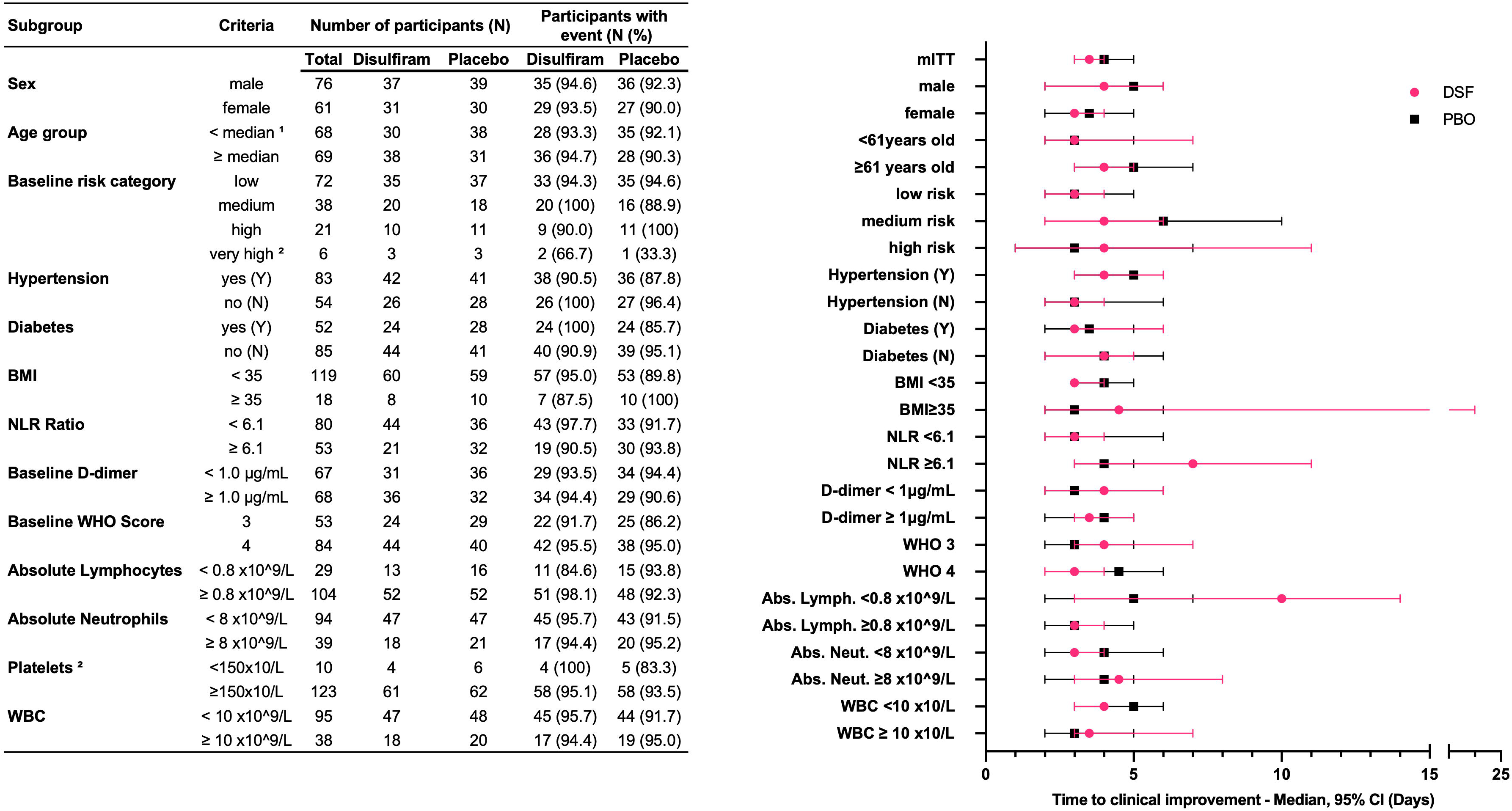
Forest plot of subgroup analyses for the primary endpoint. The table on the left represent the number of participants in the subgroup and the participant with events. The time to improvement is defined as the time (in days) from baseline to the earliest day of improvement (decrease of at least one point from baseline) at any post baseline WHO assessment. Forest plot on the right is a representation of Kaplan-Meier estimate of time to clinical improvement (days), median, 95% CI. **Abbreviations:** Abs.: absolute; CI: Confidence Interval; mITT: modified Intent-to-Treat; lymph.: lymphocytes; NLR: Neutrophil to lymphocyte ratio; WHO: World Health Organization **Footnotes:**

1. 61 is the median for the age group.
2. Not represented on the plot because of the small number of participants in the group. **Notes:**

1. 4 subjects have missing data for NLR ratio at baseline.
2. 2 subjects have missing data for D-dimer level at baseline.
3. 4 subjects have missing data for Absolute Lymphocytes at baseline.
4. 4 subjects have missing data for absolute neutrophils at baseline.
5. 4 subjects have missing data for platelets count at baseline.
6. 4 subjects have missing data for WBC at baseline.

### Exploratory Endpoints

In addition to the clinical endpoints, pro-inflammatory cytokines (IL-1β, IL-6, IL-18, TNF-α) and cf-DNA levels were measured in participants’ peripheral blood at baseline, day 8, and day 15. However, no significant changes from baseline were observed within or between the arms (data not shown).

### Safety

AEs were more frequent in the disulfiram group (67.6%) compared to placebo (37.7%). However, the reported events were common among the critically ill population under study. The incidence of SAEs was similar between groups (8.8% disulfiram, 7.2% placebo), and none of the SAEs were related to the study drug. AEs leading to discontinuation were low (1.5% disulfiram, 0% placebo). Five deaths occurred, 3 in disulfiram and 2 in placebo (**Table 2**). None of the deaths occurred by day 8 visit, 3 occurred during the treatment phase, and 2 during the follow-up period (**Figure 1**). One death in disulfiram was a participant who withdrew consent before day 15 visit and died at day 20.

The difference in AEs between groups was mainly driven by moderate lactate dehydrogenase (LDH) elevation, D-dimer increases, and dyspnea (**Table 3**). Hyperglycemic AEs were similar between arms. Hypertensive AEs occurred in 11 disulfiram participants and 2 placebo participants, with 9 disulfiram participants already on anti-hypertensive medications at baseline. None were reported as SAEs, although one participant in the placebo arm had an event of hypertensive crisis.

**Table 3.** Five most reported Adverse Events in the disulfiram group.

|  | **Disulfiram**<br>**N=68**<br>**Subjects n (%)** | **Placebo**<br>**N=69**<br>**Subjects n (%)** |
| --- | --- | --- |
| Elevated LDH | 11 (16) | 5 (7) |
| Hypertensive AEs | 11 (16) | 2 (3) |
| Dyspnea | 10 (15) | 7 (10) |
| Increased D-dimer | 10 (15) | 6 (9) |
| Hyperglycemia | 8 (12) | 7 (10) |

## Discussion

This randomized, double-blind, placebo-controlled trial tested the efficacy and safety of disulfiram in patients with COVID-19. The addition of disulfiram to standard of care did not improve clinical outcomes in moderate COVID-19 patients. No significant differences were observed in the analyses of primary and secondary outcomes in either the mITT population or in predefined subgroups.

Inhibition of inflammasome pathways and pyroptosis is being explored as potential treatments for COVID-19 [4,31–33]. However, other inhibitors such as dimethyl fumarate (GSDMD inhibitor) and DFV890 (NLRP3 inhibitor) have not shown significant improvements in clinical outcomes[34,35].

Systemic corticosteroids, particularly dexamethasone, are commonly used in hospitalized COVID-19 patients requiring supplemental oxygen to reduce inflammation and improve outcomes[36–38]. The RECOVERY trial showed that a cumulative dose of 60 mg of dexamethasone reduced mortality in patients requiring mechanical ventilation and supplemental oxygen[38]. In the present study, 73% of the safety population received systemic corticosteroids prior to randomization (73.5% disulfiram, 72.5% placebo) and 81% during the study (77.9% disulfiram, 84.1% placebo). Dexamethasone was the most frequently administered corticosteroid (>90%) but some participants received prednisolone, methylprednisolone, prednisone, or hydrocortisone. To standardize the comparison, all corticosteroid doses were converted into dexamethasone equivalent doses disregarding their mineralocorticoid effect since it is not their primary mechanism of action in treating COVID-19. In the safety population, 41.61% of participants (48.5% disulfiram, 34.8% placebo) received a dexamethasone equivalent cumulative dose ≥60 mg (dose tested in the RECOVERY trial[38]) and 78.1% received a dexamethasone equivalent cumulative dose ≥20 mg (76.5% disulfiram vs 79.7% placebo). While 20 mg represents a third of the CDC recommended dose, the lowest efficacious dose has yet to be defined. Corticosteroids use may have blunted the anti-inflammatory effects of disulfiram, potentially preventing the study from showing any beneficial effect.

In addition, both the use of corticosteroids and the improved standard of care may have led to improved clinical outcomes[39] and therefore decrease the power of the study, in particular for endpoints looking at disease progression. As of August 2020, the rate of progression to severe disease or death for hospitalized COVID-19 patients was over 20%, reaching up to 40% in high-risk cohorts (advanced age and comorbidities) [40–43]. In this study, only 16.1% (15.9% in placebo) transitioned to severe COVID-19.

Disulfiram is considered safe for treating alcohol dependence. In this study, more AEs were reported in disulfiram, but no safety concerns related to the study drug were identified. Biomarkers levels associated with COVID-19, including LDH and D-dimer, were elevated in both treatment groups[44–47]. Hypertensive AEs were more common in disulfiram, but a large proportion of the study population had pre-existing hypertension (61.8% disulfiram, 59.4% placebo), and half of the study population was taking anti-hypertensive medication at baseline, including 9 of the 11 disulfiram-treated participants with hypertensive AEs. Baseline heart rate and blood pressures were comparable across treatment arms. However, in the disulfiram group, we noted modest elevations in mean systolic and diastolic blood pressures between baseline and day 8 visit. Given the small sample size, concurrent illnesses, corticosteroids use, and the high prevalence of hypertension at baseline, the clinical significance of this observation remains uncertain. Notably, hypertension has not been reported as a common adverse reaction to disulfiram. Overall, this study did not demonstrate any unique safety concerns associated with the use of disulfiram in this population. Further research is needed to fully understand the relationship between disulfiram, hypertension, and potential interactions with other COVID-19 medications.

Elevated NLR and severe lymphopenia are both markers associated with severe infection or inflammation. In COVID-19 patients, they have been identified as independent risk factors for mortality and linked to poorer prognosis[48–51]. Subgroup analysis based on elevated NLR showed increased time to clinical improvement, particularly in disulfiram (*P*=.06), possibly due to an imbalance in severe lymphopenia at baseline (7 disulfiram,1 placebo).

The safety profile of disulfiram in this population aligned with its established profile for treating alcohol dependence. However, it did not show significant benefit in clinical outcomes for moderate COVID-19. Overall, study findings do not support the use of disulfiram in hospitalized patients with moderate COVID-19, in addition to the standard of care.

## Supporting information

Appendix 1: protocol

Appendix 2: SAP

CONSORT checklist

## Data Availability

All data produced in the present study are available upon reasonable request to the authors.

## Acknowledgments

The clinical trial was made possible through generous donations to the Spring Research Foundation, a non-profit organization that supported this study. The authors declare no conflicts of interest. The authors express their gratitude to study participants who volunteered and contributed to the scientific knowledge and public health goals of the research. We acknowledge the contributions of the research staff, including investigators, co-investigators, research coordinators, and data managers. Our thanks go to Alvaro Machado for his pharmacy-related support, and to all staff members at ETICA Institute, the local IRB, and CONEP for their efficient handling of regulatory processes. CSSI LifeSciences provided resources, infrastructure, and data management services for the study, and we are especially thankful to Janice Cattano, Senior Director of clinical operations at CSSI, for her invaluable support and collaboration throughout the study. We also thank Daniela Dorta, onsite clinical monitor, and Helena Gomez, remote clinical monitor, for ensuring data quality and completeness, which were essential for the successful completion of the study. Tigermed provided expertise in statistical analysis and programming of the Electronic Data Capture system. We are grateful to their staff for their support, which was key in ensuring accurate and efficient data collection and data analysis. We acknowledge the members of the DSMB for their oversight of the safety and efficacy data: Dr. Dan Jorgensen (physician), Dr. Kevin Lye (physician), and Terri Sampo (biostatistician). Their valuable contributions were essential for ensuring the validity and ethical conduct of the trial. We acknowledge Maria Raquel Venturin Cosate, Gisele Santos Gonçalves and Santuza Maria Ribeiro Teixeira at CT Vacinas, for performing the immunological experiments. We also thank the entire Spring Discovery team, and in particular Dr. Lauren Nicolaisen and Dr. Colin Fuller, for their support, insightful discussions, and posthoc analysis. Finally, we express our gratitude to Dr. Judy Lieberman and Dr. Hao Wu, who provided critical support and advice in various aspects of the study.

## Abbreviations

AEs: adverse events
ARDS: acute respiratory distress syndrome
BMI: body mass index
cf-DNA: cell-free DNA
CONSORT: consolidated standards of reporting trials
DAIDS: division of AIDS
DSMB: data and safety monitoring board
GSDMD: gasdermin D
ICF: informed consent form
ICU: intensive care unit
IL: interleukin
LDH: lactate dehydrogenase
LS: least square
mITT: modified intention-to-treat
NETosis: neutrophil extracellular traps formation
+NLR: neutrophil-to-lymphocyte ratio
SAEs: serious adverse events
SAP: statistical analysis plan
SE: standard error
TNF: tumor necrosis factor alpha
WHO: World Health Organization

