## Supplemental Methods for "Safety and Efficacy of Disulfiram in Hospitalized Patients with Moderate COVID-19: A Randomized, Double-Blind, Placebo-Controlled Trial"

### Ethics

The study entitled “A Randomized, Double-Blind, Placebo-Controlled Safety and Clinical Outcomes Study of Disulfiram in Subjects With Moderate COVID-19” received approval from the National Research Ethics Committee (Comissão Nacional de Ética em Pesquisa (CONEP), approval number: 4.408.019 on 11/18/2020) and from local Research Ethics Committees (Comitês de Ética em Pesquisa (CEP)): Ensino e Terapia de Inovacao Clinica AMO (ETICA), approval number: 4.280.370 on 09/16/2020; Hospital Bahia, approval number: 4.472.182 on 12/17/2020; Hospital Portugues, approval number: 4.592.906 on 03/16/2021.

### Randomization, Stratification, and Masking

The four strata were designed to assess the risk of poor clinical outcomes based on age, hypertension, diabetes, and BMI. Hypertension and diabetes were adjudicated based on the current use of any prescription medication to treat high blood pressure, or any prescription of oral hypoglycemics or insulin injections, respectively. Additionally, a BMI of 35 or higher was also considered a risk factor. The four strata were defined as low risk, medium risk, high risk, and very high risk, with expected worse clinical outcomes in each of the higher risk groups as described in **Table S2.**

A random allocation sequence was generated by a statistician analyst using a SAS® Version 9.4 program, employing random permuted block sizes of 4 within each stratum. Kit numbers were used to implement the random allocation sequence. Disulfiram and placebo were provided in capsule form and had identical appearances. Unblinded pharmacists prepacked them into identical bottles and labeled them with kit numbers. These labeled bottles were stored in a central pharmacy that was shared among the four sites. The allocation sequence was concealed by assigning kit numbers via the centralized electronic system once participants were enrolled.

The only individuals with access to unblinded information were: a statistician analyst, who was not directly involved in the study but responsible for generating the allocation sequence and providing treatment codes to unblinded pharmacists; two unblinded pharmacists responsible for labeling the kits; an independent clinical research associate (CRA), separate from the study monitors who supervised the compounding of the capsules and the labeling of the kits; and a statistician who prepared the data for DSMB.

### Procedures

At screening and baseline, the medical history, demographics, medications, vitals, weight, height, and peripheral oxygen saturation (SpO2) of participants were recorded. In addition, a physical exam was conducted, and a blood sample was collected for complete blood cell count (CBC) and serum chemistry, including liver and kidney function. Female participants of childbearing potential underwent a hCG pregnancy test, and a PCR or rapid antigen test was performed to confirm SARS-CoV-2 infection unless a positive result was obtained within the last 7 days.

At baseline, day 8 visit, and day 15 visit, vital signs, SpO2, and concomitant medications were recorded, and blood samples were collected to evaluate CBC, serum chemistry, D-dimer, and biomarkers. Additionally, the WHO score was assessed, a physical exam was conducted, and a 12-lead ECG was performed at baseline. A chest CT was performed at baseline for all participants and at day 15 visit for those still hospitalized. AEs were recorded at day 8 and day 15 visits for all participants. For hospitalized participants, vital signs, SpO2, WHO score, concomitant medications, and AEs were recorded daily. If participants were discharged from the hospital prior to the end-of-treatment visit (day 15), they continued to take the investigational drug at home to complete the 14-day treatment period and returned to the site for day 15 visit. A follow-up visit was performed on day 28, either in person or remotely; concomitant medications and AEs were recorded, and the WHO score was assessed.

More details are available in the trial protocol **in Appendix 1**.

### Statistical Analysis

The baseline value for efficacy and safety data were defined as the last non-missing observed data prior to the first dose of study drug.

The primary efficacy endpoint was the time to clinical improvement of one or more points on the WHO 7-point ordinal scale using the mITT population. Improvement was defined as a decrease of at least one point on the WHO scale compared to the baseline value. The time to improvement was measured in days from baseline to the earliest day of improvement at any post-baseline WHO assessment. The hypothesis test for the difference between disulfiram and placebo in the primary efficacy endpoint of time to improvement through day 28 was performed using a log-rank test.

Participants who were lost to follow-up or discontinued from the study prior to day 28 visit and before the event was observed, were censored at their last assessment. Participants who died were assumed to have been censored at day 28 for the purpose of analyzing time to improvement, with death assigned the worst rank.

Subgroup analyses were conducted to evaluate the homogeneity of treatment effect across demographic, comorbidity, and other baseline characteristic subgroups on the primary and key secondary endpoints as detailed in the SAP (**Appendix 2**).

The hypothesis testing to compare the mean number of days on supplemental oxygen between two treatment groups was tested using an analysis of covariance (ANCOVA) model with treatment as a fixed effect, adjusted for covariates including baseline risk category and the interaction between treatment groups and baseline risk categories. Death at any time point was assigned a score of 28 days. Significance testing was based on least square (LS) Means. LS means (95% CI of LS means), standard error (SE), LS mean vs. placebo, 95% CI and p-value from above mentioned ANCOVA model was presented.

Time from baseline to discharge was analyzed in a similar manner to the primary efficacy endpoint, as described above. The time to discharge was defined as the duration (in days) from baseline to the participant's discharge from the hospital.

The hypothesis testing to compare the percentage of participants who were discharged by day 8 between the two treatment groups was conducted using logistic regression. The independent variables in the model were the treatment group and baseline risk category. Along with the aforementioned analysis, the frequency and percentage of participants were presented by treatment group together with the two-sided 95% Clopper-Pearson confidence interval.

The mean number of days on non-invasive ventilation, high-flow oxygen devices, or invasive mechanical ventilation over a 28-day period was assessed using WHO scores of 5, 6, or 7. The same approach as the first secondary endpoint was used for the above and other analyses that compared mean number of days.

The Chi-square test was used to compare the proportions of participants who died from any cause between the groups. In cases where the expected cell frequency was less than 5, Fisher's exact test was used for the analysis, and the corresponding confidence interval for the difference in proportion was calculated.

The safety population was used for all safety analyses, and it consisted of all subjects who were randomized and received at least one dose of the study drug.

### Important changes after trial commencement

The protocol amendments resulted in important changes, which are summarized below.

#### Study Population – Eligibility criteria

Throughout the enrollment period, the elderly remained at risk, but we observed a shift in the age distribution of hospitalized patients. The rate of hospitalization increased among younger adults, likely due to the vaccination rollout targeting the elderly and the emergence of a COVID-19 variant affecting younger individuals in Brazil. Consequently, hospitalization rates for older adults declined while rates for adults under 50 years of age increased during the trial. The standard of care also improved, resulting in better clinical outcomes and reduced reliance on mechanical ventilation[1]. Participants with milder symptoms (WHO 3) were discharged within a few days, and fewer participants than expected died or needed mechanical ventilation[2–5].

To ensure the enrollment of the population most affected by COVID-19 during the study, the protocol was amended to include individuals aged 35 years and older regardless of their comorbidities (comorbidities were still used to define the strata). To enhance the population with those likely to benefit from the investigational product, we limited enrolment to moderate cases of COVID-19 requiring supplemental oxygen (WHO Score of 4) to enhance the population with those likely to benefit from the investigational product. We also excluded fully vaccinated participants.

#### Outcome measure

During the clinical trial, several factors prompted changes to the endpoints. The widespread use of corticosteroids by most patients at baseline had the potential to impact their baseline cytokine levels in particularly the levels of IL-18, which was the initial primary endpoint (change from baseline to day 8 for the cytokine IL-18). To minimize any confounding factors and ensure the accuracy of treatment evaluation we modified the primary endpoint to assess the time to clinical improvement, as defined by an improvement of ≥1 point in the WHO score. Furthermore, the standard of care for COVID-19 patients improved during the trial resulting in overall better clinical outcomes, reduced reliance on mechanical ventilation[1], and a lower rate of disease progression to severe stages than initially anticipated[2–5], potentially affecting the statistical power for endpoints related to disease progression. Consequently, secondary endpoints were reevaluated and reordered to align with the changing dynamics of the study population.

3. Docherty AB, Harrison EM, Green CA, Hardwick HE, Pius R, Norman L, et al. Features of 20 133 UK patients in hospital with covid-19 using the ISARIC WHO Clinical Characterisation

Protocol: prospective observational cohort study. Bmj. 2020;369: m1985. doi:10.1136/bmj.m1985

4. Garg S, Kim L, Whitaker M, O'Halloran A, Cummings C, Holstein R, et al. Hospitalization Rates and Characteristics of Patients Hospitalized with Laboratory-Confirmed Coronavirus Disease 2019 — COVID-NET, 14 States, March 1–30, 2020. Morbidity Mortal Wkly Rep. 2020;69: 458–464. doi:10.15585/mmwr.mm6915e3

5. Wiersinga WJ, Rhodes A, Cheng AC, Peacock SJ, Prescott HC. Pathophysiology, Transmission, Diagnosis, and Treatment of Coronavirus Disease 2019 (COVID-19). Jama. 2020;324: 782–793. doi:10.1001/jama.2020.12839
