## Supplementary material for "Safety and Efficacy of Disulfiram in Hospitalized Patients with Moderate COVID-19: A Randomized, Double-Blind, Placebo-Controlled Trial": Figure S1

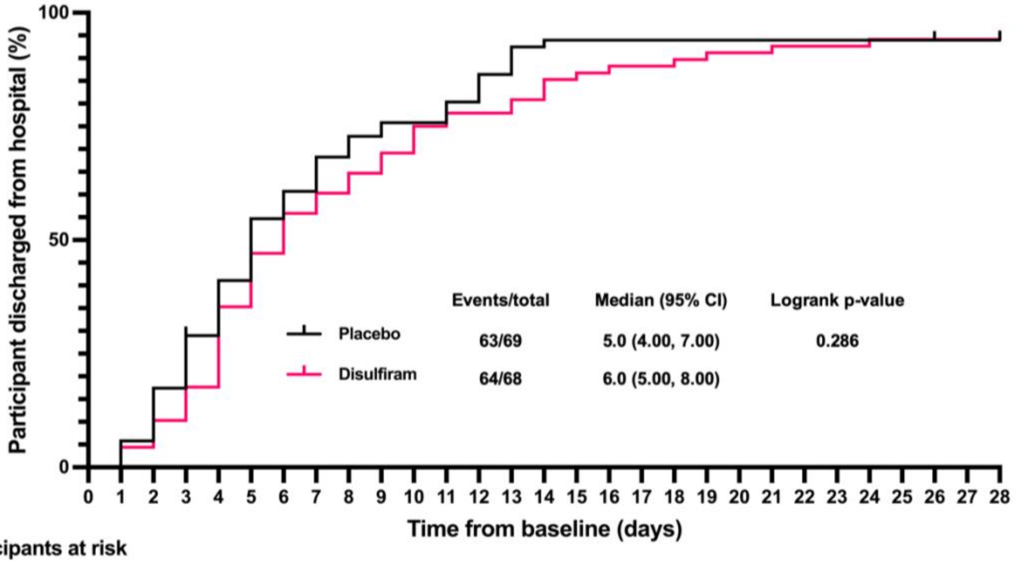


**Participants at risk**

| Days elapsed | 0 | 1 | 2 | 3 | 4 | 5 | 6 | 7 | 8 | 9 | 10 | 11 | 12 | 13 | 14 | 15 | 16 | 18 | 19 | 21 | 24 | 26 | 28 |
|---|---|---|---|---|---|---|---|---|---|---|---|---|---|---|---|---|---|---|---|---|---|---|---|
| Placebo | 69 | 69 | 65 | 57 | 47 | 39 | 30 | 26 | 21 | 18 | | 16 | 13 | 9 | 5 | | | | | | | 4 | 3 |
| Disulfiram | 68 | 68 | 65 | 61 | 56 | 44 | 36 | 30 | 27 | 24 | 21 | 17 | | 15 | 13 | 10 | 9 | 8 | 7 | 6 | 5 | | 4 |

### Figure S1. Time to clinical improvement

Kaplan-Meier plot representing the time to clinical improvement over 28 days period by treatment group in the mITT population. Clinical improvement is defined as the time (in days) from baseline to the earliest day of improvement (decrease of at least one point from baseline) at any post-baseline WHO assessment. The ticks represent censored participants only.

**Abbreviations:** CI: Confidence Interval; HR: Hazard Ratio; mITT: modified Intent-to-Treat; WHO: World Health Organization.
