## Supplementary material for "Safety and Efficacy of Disulfiram in Hospitalized Patients with Moderate COVID-19: A Randomized, Double-Blind, Placebo-Controlled Trial": Figure S2

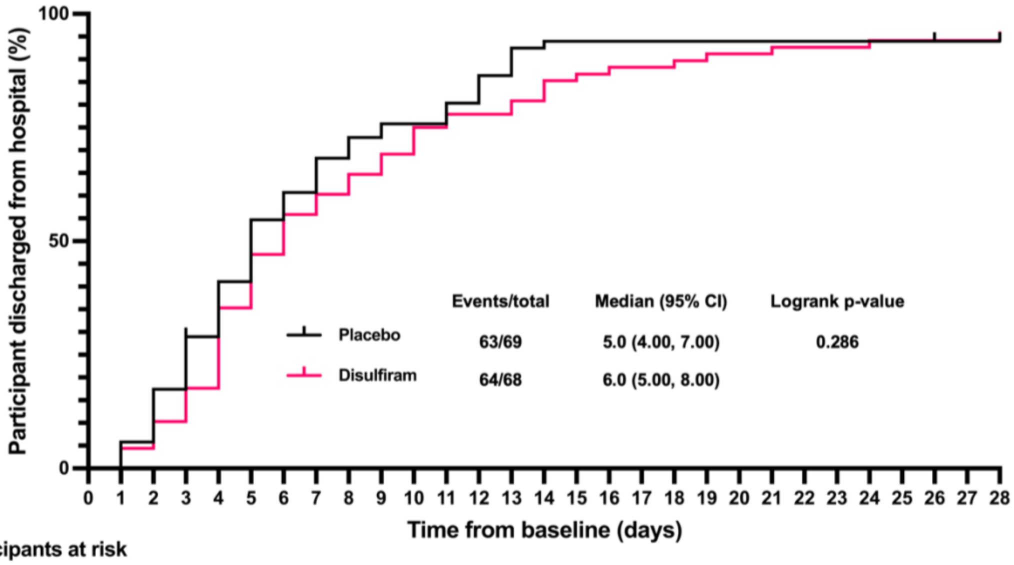


**Participants at risk**

| Days elapsed | 0 | 1 | 2 | 3 | 4 | 5 | 6 | 7 | 8 | 9 | 10 | 11 | 12 | 13 | 14 | 15 | 16 | 18 | 19 | 21 | 24 | 26 | 28 |
|---|---|---|---|---|---|---|---|---|---|---|---|---|---|---|---|---|---|---|---|---|---|---|---|
| Placebo | 69 | 69 | 65 | 57 | 47 | 39 | 30 | 26 | 21 | 18 | | 16 | 13 | 9 | 5 | | | | | | | 4 | 3 |
| Disulfiram | 68 | 68 | 65 | 61 | 56 | 44 | 36 | 30 | 27 | 24 | 21 | 17 | | 15 | 13 | 10 | 9 | 8 | 7 | 6 | 5 | | 4 |

### Figure S2. Time to discharge from the hospital

Kaplan-Meier plot representing the time to discharge from the hospital over 28 days period by treatment group in the mITT Population. The time to discharge is defined as the time (in days) from baseline to when the subject is first discharged from the hospital. The ticks represent censored participants only.

**Abbreviations:** CI: Confidence Interval; mITT: modified Intent-to-Treat; WHO: World Health Organization.
