## Supplementary material for "Safety and Efficacy of Disulfiram in Hospitalized Patients with Moderate COVID-19: A Randomized, Double-Blind, Placebo-Controlled Trial": Table S1

| WHO score | Descriptor | Covid stage |
|---|---|---|
| 1 | Not hospitalized, no limitations on activities | Mild |
| 2 | Not hospitalized, limitation on activities | |
| 3 | Hospitalized, not requiring supplemental oxygen | Moderate |
| 4 | Hospitalized, requiring supplemental oxygen | |
| 5 | Hospitalized, on non-invasive ventilation or high flow oxygen devices | Severe |
| 6 | Hospitalized, on invasive mechanical ventilation or ECMO | |
| 7 | Death | |

### Table S1. WHO 7-point ordinal clinical progression scale

**Abbreviations:** WHO: World Health Organization. ECMO: extracorporeal membrane oxygenation
