## Supplementary material for "Safety and Efficacy of Disulfiram in Hospitalized Patients with Moderate COVID-19: A Randomized, Double-Blind, Placebo-Controlled Trial": Table S2

| | Comorbidities | |
|---|---|---|
| **Age** | 0 | 1 or more |
| 35-59 | Low | Low |
| 60-69 | Low | Medium |
| 70-79 | Medium | High |
| ≥ 80 | High | Very High |

### Table S2. Tiered risk schema stratification

Comorbidities include hypertension (on any prescription anti-hypertension medication), diabetes (on oral or insulin injections), and BMI ≥ 35.
