## Supplementary material for "Safety and Efficacy of Disulfiram in Hospitalized Patients with Moderate COVID-19: A Randomized, Double-Blind, Placebo-Controlled Trial": Table S3

| | Disulfiram (N=68) | | | | Placebo (N=69) | | | |
|---|---|---|---|---|---|---|---|---|
| **Comorbidities** | **0 n (%)** | **1 n (%)** | **2 n (%)** | **3 n (%)** | **0 n (%)** | **1 n (%)** | **2 n (%)** | **3 n (%)** |
| **Age group (Years)** | | | | | | | | |
| **<35** | 0 | 0 | 0 | 0 | 0 | 0 | 0 | 0 |
| **35 - 59** | 8 (11.8) | 16 (23.5) | 4 (5.9) | 2 (2.9) | 8 (11.6) | 18 (26.1) | 7 (10.1) | 1 (1.4) |
| **60 - 69** | 5 (7.4) | 9 (13.2) | 6 (8.8) | 1 (1.5) | 3 (4.3) | 8 (11.6) | 6 (8.7) | 1 (1.4) |
| **70 - 79** | 4 (5.9) | 5 (7.4) | 5 (7.4) | 0 | 3 (4.3) | 3 (4.3) | 6 (8.7) | 1 (1.4) |
| **≥ 80** | 0 | 1 (1.5) | 2 (2.9) | 0 | 1 (1.4) | 2 (2.9) | 1 (1.4) | 0 |

### Table S3. Age and Comorbidities Distribution in the Safety Populations

Comorbidities include hypertension (on any prescription anti-hypertension medication), diabetes (on oral or insulin injections), and BMI ≥ 35.

**Abbreviation:** N: Number of subjects in the safety population in each treatment group.

**Note:** Percentages are based on the number of treated subjects per treatment group N.
