## Appendix 1: protocol for "Safety and Efficacy of Disulfiram in Hospitalized Patients with Moderate COVID-19: A Randomized, Double-Blind, Placebo-Controlled Trial"

### A Randomized, Double-blind, Placebo-controlled Safety and Clinical Outcomes Study of Disulfiram in Subjects with Moderate COVID-19

| | |
|---|---|
| Protocol Number: | SPR-001-201 |
| Short Title: | SPRING Study |
| Original Protocol Date: | 03 September 2020 |
| Version: | 1.0 |
| Version: | 1.1 |
| Date of Revision | 20 October 2020 |
| Version: | 1.2 |
| Amendment Date: | 07 December 2020 |
| Version: | 1.3 |
| Amendment Date: | 21 May 2021 |
| Version: | 1.4 |
| Amendment Date: | 09 July 2021 |

**CONFIDENTIALITY STATEMENT**
The information contained in this document and all information provided to you related to the SPRING Study is confidential and proprietary information and, except as may be required by federal, state, or local laws or regulations, may not be disclosed to others without prior written permission. The Principal Investigator may disclose such information to supervised individuals working on this protocol, provided such individuals agree to maintain the confidentiality of such information.

#### STATEMENT OF COMPLIANCE

| | |
|---|---|
| Study Title | A Randomized, Double-blind, Placebo-controlled Safety and Clinical Outcomes Study of Disulfiram in Subjects with Moderate COVID-19 |
| Protocol Number | SPR-001-201 |
| Original Protocol Date of Issue: | 03 September 2020 |
| Version 1.1 | 20 Oct 2020 |
| Version 1.2 | 07 December 2020 |
| Version 1.3 | 21 May 2021 |
| Version 1.4 | 09 July 2021 |
| Investigator Name and Address: | Augusto Mota, MD, PhD<br>Rua Altino Serbeto de Barros, 119<br>14º Andar Sala 1408<br>Pituba Salvador-Ba<br>CEP 41830-492 |

The study will be carried out in accordance with the following, as applicable:

- All National and Local Regulations and guidelines for the conduct of clinical research to ensure the safety and well-being of study subjects and the scientific integrity of the trial.

I, the undersigned, have read and approve this protocol and agree to conduct the study according to the protocol.

I agree to comply with the current ICH and GCP guidelines.

I agree to conduct the study in person or to supervise the study.

I agree to ensure that all who assist me in the conduct of the study have access to the study protocol and any amendments and are aware of their obligations.

________________________________________
Name/Title

________________________________________ ____________________
Investigator Signature Date

SUMMARY OF CHANGES

Amendment 1: The protocol has been amended to incorporate administrative changes and correct typographical errors. The details of the changes are summarized in the table below.

| Version | Date | Page | Change | Reason for Change |
|---|---|---|---|---|
| Original | 03 Sept 2020 | N/A | Original Version | New Document |
| Version 1.1 | October 20, 2020 | In | Revised according to CONEP recommendations | New Document |
| Version 1.2 | 02 December 2020 | Page 2, Cover | Added new version number and date of new version | Clarification |
| Version 1.2 | 02 December 2020 | Page 12, Protocol Summary, Number of Sites | Revised from 4 sites to 8 sites | To enhance enrollment |
| Version 1.2 | 02 December 2020 | Page 13, Protocol Summary, Secondary Endpoints | Move: To assess the percentage of subjects that are not admitted to the ICU from other secondary endpoints to number 6 in secondary endpoints. | Typographical error with the placement of the endpoint in the list. |
| Version 1.2 | 02 December 2020 | Page 13, Section 1.1 Synopsis, Secondary Outcomes | Added: Evaluate the percentage of individuals who are not admitted to the ICU of other secondary outcomes for number 6 in secondary outcomes. | Typographical error with the end point being placed in the list. |
| Version 1.2 | 02 December 2020 | Page 14, Section 1.1 Synopsis, Inclusion criteria | Revise inclusion criteria 1 for subjects aged 50 to < 60 with at least one of the following co-morbidities: BMI ≥ 35 or hypertension or diabetes. | |
| Version 1.2 | 02 December 2020 | Page 14, Section 1.1 Synopsis, Inclusion criteria | Revise inclusion criteria 9 to include the use of rapid antigen test and COVID testing within 7 days of screening. | |
| Version 1.2 | 02 December 2020 | Page 14, Section 1.1 Synopsis, Inclusion criteria | Added to item 7, after "ambient air": "or using supplementary O2 via nasal cannula with flow of up to 3 l/min. | Clarification |
| Version 1.2 | 02 December 2020 | Page 14, Section 1.1 Synopsis, Inclusion criteria | Added to item 8, after "3 days": "counting only days if week". | Clarification |

| | | | | |
|---|---|---|---|---|
| Version 1.2 | 02 December 2020 | Page 15, Protocol Summary Baseline Day 1 | Add 12-lead ECG (if subject had ECG already performed for this hospitalization, if can be used). | Typographical error of omission. Needs to match body of the protocol |
| Version 1.2 | 02 December 2020 | Page 17, Protocol Summary, Day 7 Visit | Remove bullet 2 Physical Exam | Typographical error. Should match body of the protocol |
| Version 1.2 | 02 December 2020 | Page 17, Protocol Summary, Day 7 Visit | Add LDH to serum chemistry | Typographical error of omission. |
| Version 1.2 | 02 December 2020 | Page 18, Protocol Summary Day 8-13 | Remove sentence: "The information can be obtained remotely." | Typographical error. Needs to match Schedule of Assessments. |
| Version 1.2 | 02 December 2020 | Page 18, Protocol Summary Day 8-13 | Remove sentence: "The subject will be assessed per the WHO Ordinal Scale." | Typographical error. This information will be included in the study procedure list. |
| Version 1.2 | 02 December 2020 | Page 18, Protocol Summary Day 8-13 | Add the following information: If the subject is in the hospital, the procedures to be performed in consultations from Day 8 to Day 13 of the study are:<br>1. Concomitant meds,<br>2. vital signs: blood pressure, heart rate, respiratory rate, axillary temperature<br>3. Oxygen saturation will be measured by pulse oximetry (SpO2<br>4. The subject will be assessed per the WHO Ordinal Scale.<br>5. it will be noted if the subject had been admitted to the ICU, requires invasive mechanical ventilation or non-invasive ventilation or supplemental oxygen (should any of these outcomes have occurred, the duration in days of such events must be recorded).<br>6. If a CT scan of the chest was performed | Typographical error. Needs to match Schedule of Assessments. |

| | | | | |
|---|---|---|---|---|
| | | | as part of standard of care, the data will be captured.<br>7. Record adverse events. | |
| Version 1.2 | 02 December 2020 | Page 18, Protocol summary, Day 14 visit | Remove Physical Exam | Typographical error. Needs to match body of the protocol. |
| Version 1.2 | 02 December 2020 | Page 18, Protocol summary, Day 14 visit | Add LDH to serum chemistry | Typographical error of omission. |
| Version 1.2 | 02 December 2020 | Page 18, Protocol summary, Day 14 visit | Add “the subject is hospitalized” and “will be”<br><br>Remove: as part of standard of care, the data will be captured. | Typographical error. Needs to match body of the protocol. |
| Version 1.2 | 02 December 2020 | Page 18, Protocol summary, Day 14 visit | Add the following to study procedure: It will be noted if the subject had been admitted to the ICU, requires invasive mechanical ventilation or non-invasive ventilation or supplemental oxygen (should any of these outcomes have occurred, the duration in days of such events must be recorded). | Typographical error. Needs to match body of the protocol. |
| Version 1.2 | 02 December 2020 | Page 19 synopsis, Day 28 visit | Remove: Oxygen saturation will be measured by pulse oximetry (sPO2) | Typographical error. Needs to match body of the protocol. |
| Version 1.2 | 02 December 2020 | Page 22, footnote 2 Schedule of Assessments | Remove:” Consultations between Day 2 and Day 6 of the Study will only occur if the participant is still in the hospital.” | Typographical error. |
| Version 1.2 | 02 December 2020 | Page 22, footnote 2 Schedule of Assessments | Change footnote 2 to: Visits only if hospitalized. | Typographical error. |
| Version 1.2 | 02 December 2020 | Page 21 Schedule of Assessment | Add supplemental Oxygen use assessment to screening and Day 1 | |
| Version 1.2 | 02 December 2020 | Page 21 Schedule of Assessment | Add rapid antigen to SARS-CoV-2 testing | |

| | | | | |
|---|---|---|---|---|
| Version 1.2 | 02 December 2020 | Page 21 Schedule of Assessment | Visit Day 28 remove vital signs and SpO2 | Typographical error. Needs to match body of the protocol. |
| Version 1.2 | 02 December 2020 | Page 32, Section 5.1 Inclusion criteria | Added to item 7, after "ambient air": "or using supplementary O2 via nasal cannula with flow of up to 3 l/min. | Clarification |
| Version 1.2 | 02 December 2020 | Page 33, Section 5.1 Inclusion criteria | Added to item8, after "less than 3 days": "counting only weekdays. | Clarification |
| Version 1.2 | 02 December 2020 | Page 34, Section 5.2 | Revise bullet 9 to add "or adenocarcinoma of the prostate with low or very low risk categories by NCCN criteria." | Typographical omission error. |
| Version 1.2 | 02 December 2020 | Page 36, Section 6.1.1, Dose Administration Route | Add, "During the hospitalization, study medication will be dispensed daily by study staff."<br><br>Add "or enteral if required"<br><br>Add "contents of the" | Provide additional clarification. |
| Version 1.2 | 02 December 2020 | Page 36, Section 6.1.1 | Remove section on administration for subjects on invasive ventilation and replace with: If the subject is unable to swallow the capsule, the capsule can be dissolved in a liquid syrup. See the pharmacy manual for specific instructions. | Study treatment will be dissolved into syrup rather than a separate liquid formulation. |
| Version 1.2 | 02 December 2020 | Page 37, Section 6.1.4 Disulfiram | Change to 15°C and 30°C | Typographical Error |
| Version 1.2 | 02 December 2020 | Page 37, Section 6.1.4 Placebo to Match | Change to 15°C and 30°C | Typographical Error |
| Version 1.2 | 02 December 2020 | Page 40, Section 9.1, screening assessments | Change "Date of Birth" to "Age"<br>Add bullet CBC and serum chemistry. | Typographical omission error. |
| Version 1.2 | 02 December 2020 | Page 41, Section 9.2 baseline visit | Add serum chemistry to label blood test | Typographical omission error. |

| | | | | |
|---|---|---|---|---|
| Version 1.2 | 02 December 2020 | Page 41 Section 9.2, baseline visit | Add, During the hospitalization, study medication will be dispensed daily by study staff. | Provide additional clarification. |
| Version 1.2 | 02 December 2020 | Page 42, Section 9.5 visit Day 8 – 13 | Delete “The subject will be evaluated by the WHO Ordinal Scale. Information can be obtained remotely. | Typographical error. Needs to match Schedule of Assessments. |
| Version 1.2 | 02 December 2020 | Page 42, Section 9.5 visit Day 8 – 13 | Add the following information: If the subject is in the hospital, the procedures to be performed in consultations from Day 8 to Day 13 of the study are:<br>8. Concomitant meds,<br>9. vital signs: blood pressure, heart rate, respiratory rate, axillary temperature<br>10. Oxygen saturation will be measured by pulse oximetry (SpO2<br>11. The subject will be assessed per the WHO Ordinal Scale.<br>12. it will be noted if the subject had been admitted to the ICU, requires invasive mechanical ventilation or non-invasive ventilation or supplemental oxygen (should any of these outcomes have occurred, the duration in days of such events must be recorded).<br>13. If a CT scan of the chest was performed as part of standard of care, the data will be captured.<br>14. Record adverse events. | Typographical error. Needs to match Schedule of Assessments. |

6.1 Study Products and Administration ... 36
6.1.1 Investigational Therapeutic and Matching Placebo ... 36
6.1.2 Handling/Accountability/Destruction ... 37
6.1.3 Formulation, Appearance, Packaging, and Labeling ... 38
6.1.4 Product Storage ... 38
6.1.5 Preparation ... 38
7 Measures to Minimize Bias: Randomization and Blinding ... 38
7.1 Study Intervention Compliance ... 39
7.2 Concomitant Therapy ... 39
7.2.1 Rescue Medicine ... 39
7.2.2 Non-Research Standard of Care ... 39
8 Study Intervention discontinuation and Subject Discontinuation/Withdrawal ... 39
8.1 Halting Criteria and Discontinuation of Study Intervention ... 39
8.1.1 Study Halting for Safety ... 39
8.1.2 Withdrawal from Randomized Treatment or from the Study ... 40
8.1.3 Discontinuation of Study Drug ... 40
8.1.4 Withdrawal of Subjects from the Study ... 40
8.1.5 Lost to Follow-up ... 41
9 Study Assessments and Procedures ... 41
9.1 Screening (Day -2 to 1) ... 41
9.2 Baseline (Day 1) ... 42
9.3 Day 2-7 Visits ... 43
9.4 Day 8 Visit (± 2 days) ... 43
9.5 Day 9-14 Visits ... 43
9.6 Day 15 Visit (± 3 days) ... 44
9.7 Day 28 Visit (± 7 days) ... 44
9.8 Early Termination Visit ... 45
9.8.1 Efficacy Assessments ... 45
9.8.2 Exploratory Assessments ... 45
9.9 Safety and Other Assessments ... 45
9.9.1 Procedures to be Followed in the Event of Abnormal Laboratory Test Values or Abnormal Clinical Findings ... 45
9.10 Adverse Events and Serious Adverse Events ... 46
9.10.1 Definition of Adverse Event (AE) ... 46

### 1 PROTOCOL SUMMARY

#### 1.1 SYNOPSIS

| | |
|---|---|
| **Study Title** | A Randomized, Double-blind, Placebo-controlled Safety and Clinical Outcomes Study of Disulfiram in Subjects with Moderate COVID-19 |
| **Study Short Name** | SPRING Study |
| **Study Design** | Prospective, randomized, double-blind, placebo-controlled study |
| **Number of Sites** | Up to 8 sites |
| **Location** | Brazil |
| **Laboratories** | Routine lab:<br>Laboratório Labchecap<br>Hospital da Bahia<br>Avenida Prof. Magalhães Neto, 1541 Térreo<br>Salvador - BA<br>41810-011<br>+55 71 3345 8200<br><br>Specialty lab:<br>CT VACINAS<br>Rua Professor José Vieira de Mendonça, 770 - Sala 206<br>Engenho Nogueira<br>Belo Horizonte – MG<br>31310-260, Brasil<br>+55 31 4401-1113 |
| **Indication** | Hospitalized subjects with moderate COVID-19 |
| **Investigational Product** | Disulfiram |
| **Dose Regimen** | Arm 1: Disulfiram 500 mg po daily x 14 days<br>Arm 2: Placebo po daily x 14 days |
| **Number of Subjects and Randomization** | Up to 200 Subjects, Randomized 1:1 |
| **Objective** | The overall objective of the study is to evaluate the clinical outcomes, safety, and the effect of disulfiram on select biomarkers as compared to placebo in hospitalized subjects with moderate COVID-19. |

| Endpoints | **Primary Endpoint**<br>The primary endpoint is to assess the time to clinical improvement, defined as the time from baseline to the first post-baseline assessment with an improvement in WHO score of ≥1 point.<br><br>**Secondary Endpoints**<br>The key secondary endpoints are:<br><br>1. To assess the mean number of days of supplemental oxygen (WHO Score ≥4).<br>2. To assess the time from baseline to discharge from the hospital.<br>3. To assess the percentage of subjects that are discharged by Day 8.<br>4. To assess the percentage of subjects that worsened 1 or more points on the WHO Ordinal Scale, from baseline to any post baseline assessment through Day 28.<br>5. To assess the mean number of days of non-invasive ventilation or high flow oxygen devices or invasive mechanical ventilation (WHO Score 5 or 6) over the 28-day period.<br>6. To assess the mean number of days subjects were in the Intensive Care Unit (ICU).<br>7. To assess the percentage of subjects that were on non-invasive ventilation or high flow oxygen devices or invasive mechanical ventilation (WHO Score 5 or 6) over the 28-day period.<br>8. To assess the 28-day mortality.<br><br>Other secondary endpoints are:<br>1. To assess the mean change and percent change from baseline to Day 8 and Day 15 for Cytokine IL-18.<br>2. To assess the percentage of subjects requiring supplemental oxygen (WHO Score ≥4) by Day 8, 15, and 28.<br>3. To assess the percentage of subjects that are discharged by Day 15 and Day 28.<br>4. To assess the percentage of subjects that worsened 1 or more points on the WHO Ordinal Scale from baseline through Day 8 and Day 15.<br>5. To assess the percentage of subjects admitted to the Intensive Care Unit.<br>6. To assess the percentage of subjects that improved 1 or more points on the WHO Ordinal Scale from baseline to Day 8, 15, and 28.<br>7. To assess the change and percent change in neutrophil count from baseline to Day 8 and 15.<br>8. To assess the change and percent change in total lymphocyte count from baseline to Day 8 and 15.<br>9. To assess the change and percent change from baseline to Day 8 and 15 for neutrophil-derived circulating free DNA (cf-DNA/NETs).<br>10. To assess the mean change and percent change from baseline to Day 8 and 15 for: |
|---|---|

| | |
|---|---|
| | a. Cytokine TNF-α<br>b. Cytokine IL-1β<br>c. Cytokine IL-1RA<br>d. Cytokine IL-6<br>e. Cytokine IL-8<br>f. Cytokine IL-10<br>g. Lactate dehydrogenase (LDH)<br>h. D-dimer<br>11. To assess the association between baseline and worst post-baseline WHO score. |
| **Safety** | Incidence of Adverse Events (AEs) and Serious Adverse Events (SAEs). Vital Signs and Laboratory data will be collected during the study period. |
| **Patient Selection** | **Inclusion Criteria**<br><br>Subjects may be enrolled in the study only if all the inclusion criteria are met.<br><br>1. Male and female subjects, age 35 or older.<br>2. Female subjects of childbearing potential must have a negative hCG (in urine or blood) pregnancy test. Non-childbearing potential is defined as postmenopausal, with amenorrhea for ≥ 12 months or ≥ 45 days post procedure for irreversible sterilization by hysterectomy, bilateral oophorectomy, or bilateral salpingectomy.<br>3. An International Ethics Committee (IEC) approved informed consent is signed and dated prior to any study-related activities.<br>4. Willing to abstain from any alcohol or alcoholic substances of any kind (including medications) within 24 hours prior to treatment and for 14 days after treatment concludes.<br>5. Have the ability to understand the requirements of the study and is willing to comply with all study procedures and visits.<br>6. Respiratory rate: ≤ 30 per minute.<br>7. Use supplemental $O_2$ via nasal cannula or equivalent.<br>8. Currently hospitalized ≤ 5 days.<br>9. PCR test or rapid antigen test confirming SARS-CoV-2. (If subject had PCR test performed within 7 days of screening, this can be used.) If a test comes back negative, a second test can be performed.<br>10. In the opinion of the investigator, able to participate in the study. |
| | **Exclusion Criteria**<br><br>Subjects may not be enrolled in the study if any of the exclusion criteria apply.<br><br>1. Admission into the Intensive Care Unit (ICU).<br>2. Clinically active Hepatitis. |

3. ALT or AST > 3 times the upper limit of normal.
4. Need for invasive or non-invasive ventilation at screening or baseline visit.
5. Stage 4 severe chronic kidney disease or requiring dialysis or estimated GFR < 30.
6. Known allergy to disulfiram.
7. Treatment with any of the medications listed below within 7 days prior to the baseline visit[1]:

| | | | | |
|---|---|---|---|---|
| Amprenavir | Dronabinol | Hydantoins | Metronidazole | Ritonavir |
| Benznidazole | Dyphylline | Idelalisib | Naltrexone | Sertraline |
| Chloral Hydrate | Ethanol | Immuno-modulatory drugs | Paclitaxel | Tinidazole |
| Cocaine | Ethotoin | Ixabepilone | Phenytoin | Tipranavir |
| Cyclosporine | Fosphenytoin | Lithium | Pimozide | Tranylcypromine |
| Dasabuvir | Guaifenesin | Mesoridazine | Pirfenidone | |

8. Participation in any other interventional trial within 30 days prior to enrollment.
9. Active malignancy (excluding basal cell carcinoma, squamous cell carcinoma, in situ cervical cancer or adenocarcinoma of the prostate with low or very low risk categories by NCCN criteria).
10. Any surgical or medical condition which in the opinion of the investigator may interfere with participation in the study or which may affect the outcome of the study.
11. Fully vaccinated for COVID-19 (number of doses as per manufacturer recommendation.

**Study Assessments and Procedures**

The study is designed to evaluate standard of care with either disulfiram or placebo.

**Screening Visit (Day -2 to 1)**

If willing to participate in the study, the subject will sign the informed consent. If the subject meets all the inclusion criteria and none of the exclusion criteria, they can be enrolled in the study. At this time, a unique subject identification number will be assigned to the patient to guarantee his anonymity during and after his participation to the study

The following assessments will be performed:

- Demographics including date of birth, sex, and race
- Concomitant medications
- Medical history
- Weight / height
- Vital signs – heart rate, respiratory rate, blood pressure, and temperature
- Oxygen saturation will be measured by pulse oximetry ($SpO_2$)

- It will be noted if the subject requires supplemental oxygen
- Physical exam
- Clinical labs including CBC, serum chemistry
- ALT, AST, and estimated GFR (if subject had tests performed within 5 days of screening, these results can be used)
- hCG pregnancy test (either blood or urine) in female subjects of childbearing potential.
- PCR or rapid antigen test confirming SARS-CoV-2 (if subject had PCR test performed within 7 days of screening, this can be used)

**Baseline (Day 1)**

The eligibility of enrolled subjects will be confirmed. If the results of the PCR or rapid antigen test confirming SARS-CoV-2 and ALT, AST, pregnancy test if applicable, and estimated GFR are available at screening, the screening and baseline visit can be combined.

The following procedures will be performed in this consultation on Day 1

1. Recording of the concomitant medications, doses, and forms of use.
2. Physical exam
3. Vital signs: blood pressure, heart rate, respiratory rate, axillary temperature
4. 12-lead ECG (if subject had ECG already performed for this hospitalization, it can be used),
5. Oxygen saturation will be measured by pulse oximetry (SpO2),
6. Chest CT scan. If the subject had a chest CT scan within 3 days of enrollment, this can be used, and a repeat scan is not needed.
7. Laboratory evaluations
    a. CBC
    b. serum chemistry (including liver function, kidney function, LDH)
    c. D-dimer levels
    d. cytokine panel (IL-1β, IL-1RA, IL-6, IL-8, IL-10, IL-18, TNF-α)
    e. Neutrophil extracellular traps (cf-DNA/NETs)
8. Chest CT scan (if subject had CT scan performed within 3 days of enrollment this can be used).
9. Determine the WHO Ordinal score.
   The subject will be assessed per the WHO Ordinal Scale. The WHO Ordinal Scale is an assessment of the current clinical status. The scale is as follows:

1. Not hospitalized, no limitations on activities
2. Not hospitalized, limitations on activities
3. Hospitalized, not requiring supplemental oxygen

4. Hospitalized, requiring supplemental oxygen
5. Hospitalized, on non-invasive ventilation or high flow oxygen devices
6. Hospitalized, on invasive mechanical ventilation or ECMO
7. Death

After collection of all medical assessments and information, treatment will be randomly assigned. Dispense the 7-day study medication to the subject. Treatment will be orally administered.
Patients will be randomized to one of two arms of the study: experimental drug, Disulfiram, or placebo drug. The pharmacist responsible for the study will be the only one to know which of the two arms of the study the patient was randomized to. After randomization, the pharmacy team will dispense the first supply of either the experimental drug, Disulfiram, or the placebo drug, according to the result of the randomization. The first supply will have enough study treatment for the first 7 (seven) days. This first supply will have two additional doses so that there is no interruption of treatment if the visit on Day 8 of the study, for whatever reason, occurs exceptionally on Day 9 or Day 10.

**Day 2-7 Visits**

If the subject is in the hospital, the procedures to be performed in consultations from Day 2 to Day 7 of the study are:

1. Recording of concomitant meds, doses, and forms of use.
2. Vital signs: blood pressure, heart rate, respiratory rate, axillary temperature.
3. Oxygen saturation will be measured by pulse oximetry (SpO2).
4. The subject will be assessed per the WHO Ordinal Scale, identical to that performed in the consultation on Day 1 of the study.
5. It will be noted if the subject had been admitted to the ICU, requires invasive mechanical ventilation or non-invasive ventilation or supplemental oxygen (should any of these outcomes have occurred, the duration in days of such events must be recorded).
6. If a CT scan of the chest was performed as part of standard of care, the data will be captured.
7. If the subject is unable to swallow the capsule, the contents of the capsule can be dissolved in a liquid syrup. This applies for the entire duration of treatment
8. Record adverse events.

**Day 8 Visit ±2 days**

This consultation will, whenever possible, be held on Day 8 of the study. However, for whatever reason, it can be performed from two days before (Day 6 of the study) or two days after (Day10 of the study) Day 8.

| | |
|---|---|
|  | The following procedures will be performed in this consultation on Day 8<br><br>1. Recording of the concomitant medications, doses, and forms of use.<br>2. Vital signs: blood pressure, heart rate, respiratory rate, axillary temperature<br>3. Oxygen saturation will be measured by pulse oximetry (SpO2),<br>4. Laboratory evaluation<br> a. CBC<br> b. serum chemistry (including liver function, kidney function, to be repeated only if the available results are out of date) LDH<br> c. D-dimer levels<br> d. cytokine panel -IL-1β, IL-1RA, IL-6, IL-8, IL-10, IL-18, TNF-α<br> e. neutrophil extracellular traps (cf-DNA/NETs)<br>5. Determine the WHO Ordinal score.<br>6. If a CT scan of the chest was performed as part of standard of care, the data will be captured<br>7. It will be noted if the subject had been admitted to the ICU, requires invasive mechanical ventilation or non-invasive ventilation or supplemental oxygen (should any of these outcomes have occurred, the duration in days of such events must be recorded).<br>8. Any study drug remaining from Days 1 to 7 will be collected.<br>9. The study treatment for Days 8 to 14 will be dispensed, and the drug accountability log updated. This new supply will have either the experimental drug, Disulfiram, or the placebo drug (without active substance), according to the initial randomization.<br>10. Record adverse events.<br><br>**Day 9-14 Visits**<br><br>If the subject is in the hospital, the procedures to be performed in consultations from Day 9 to Day 14 of the study are:<br>1. Concomitant meds<br>2. Vital signs: blood pressure, heart rate, respiratory rate, axillary temperature<br>3. Oxygen saturation will be measured by pulse oximetry (SpO2)<br>4. The subject will be assessed per the WHO Ordinal Scale.<br>5. It will be noted if the subject had been admitted to the ICU, requires invasive mechanical ventilation or non-invasive ventilation or supplemental oxygen (should any of these outcomes have occurred, the duration in days of such events must be recorded).<br>6. If a CT scan of the chest was performed as part of standard of care, the data will be captured.<br>7. Record adverse events. |

| | |
|---|---|
|  | **Day 15 Visit ±3 days**<br><br>This consultation will, whenever possible, be held on Day 15 of the study. However, for whatever reason, it can be done from three days before (Day 12 of the study) to three days after (Day18 of the study) Day 15.<br><br>The following procedures will be performed in this consultation on Day 15:<br><br>1. Recording of the concomitant medications, doses, and forms of use.<br>2. Vital signs: blood pressure, heart rate, respiratory rate, axillary temperature<br>3. Oxygen saturation will be measured by pulse oximetry (SpO2),<br>4. Laboratory evaluation<br> a. CBC<br> b. serum chemistry (including liver function, kidney function, to be repeated only if the available results are out of date) LDH<br> c. D-dimer levels<br> d. Cytokine panel IL-1β, IL-1RA, IL-6, IL-8, IL-10, IL-18, TNF-α<br> e. Dosage of neutrophil extracellular traps (cf-DNA/NETs)<br>5. Determine the WHO Ordinal score<br>6. If the subject is hospitalized, a CT scan of the chest will be performed.<br>7. It will be noted if the subject had been admitted to the ICU, requires invasive mechanical ventilation or non-invasive ventilation or supplemental oxygen (should any of these outcomes have occurred, the duration in days of such events must be recorded).<br>8. Any study drug remaining will be collected, and the drug accountability log updated.<br>9. No new study drug supplies will be provided. On that day, the treatment will have ended<br>10. Record adverse events.<br><br>**Day 28 Visit ±7 days**<br><br>This consultation will, whenever possible, be held on Day 28 of the study. However, for whatever reason, it can be performed from seven days before (Day 21 of the study) to seven days after (Day 35 of the study) Day 28. This visit can be conducted remotely.<br><br>The following procedures will be performed in this consultation on Day 28: |

| | |
|---|---|
| | 1. Recording of the concomitant medications, doses, and forms of use.<br>2. Determine the WHO Ordinal score<br>3. It will be noted if the subject had been admitted to the ICU, requires invasive mechanical ventilation or non-invasive ventilation or supplemental oxygen (should any of these outcomes have occurred, the duration in days of such events must be recorded).<br>4. Record adverse events. |
| **Sample Size Estimation and Statistical Analysis** | **Sample Size Estimation**<br><br>Since the efficacy of disulfiram on COVID-19 infected subjects is not known, the sample size for this study is not based on any formal calculations. This study will provide adequate data to assess potential benefits as measured by primary and key secondary clinical outcomes.<br><br>**Statistical Analyses**<br><br>Continuous data will be summarized by presenting the number of subjects with available data, number of subjects with missing data, mean, standard deviation, minimum value, maximum value and median.<br><br>Categorical data will be summarized by presenting the number and percentage of subjects in each category based on the total number of subjects with available data.<br><br>Subject disposition, demographics, and other baseline characteristics will be summarized for each arm.<br><br>Clinical efficacy outcomes will be evaluated by changes between the treatment arms, and where possible the relationships between them.<br><br>In order to control overall Type I error at 0.05, the primary endpoint will be tested first followed by each of the key secondary efficacy endpoints hierarchically in the order above.<br><br>Hypothesis testing will be performed to evaluate the difference in Kaplan-Meier (KM) curves of time to improvement from Baseline to Day 28 between treatment groups to assess statistical significance.<br><br>Hypothesis testing will be performed on continuous clinical outcomes for post baseline visits and also to compare changes in biomarkers from baseline to post-baseline visits between treatment groups to assess the difference. |

| | |
|---|---|
| | Hypothesis testing will also be performed on categorical data to evaluate percentage of responders between the treated and placebo groups.<br><br>Key safety data including any Deaths, Serious Adverse Events, Adverse Events leading to discontinuation of study medication or discontinuation from the study, Vital Signs, Laboratory data, etc. will be summarized for each arm.<br><br>Details of above will be included in the Statistical Analysis Plan which will be finalized prior to locking the database and unblinding the study. |

#### 1.2 Study Phase

Phase 2.

#### 1.3 Study Population

Hospitalized male and female subjects ≥ 35 years of age with moderate COVID-19.

#### 1.4 Sites

Up to 8 sites.

#### 1.5 Study Duration

The study may last for up to 12 months.

##### 1.5.1 Participant Duration

An individual subject will complete the study within 37 days, from screening (Day -2 to 1) to follow-up on Day 28 ±7 days.

##### 1.5.2 Safety

Given the severity of illness in COVID-19, there are no pre-specified study stopping rules for safety. Serious Adverse Events will be shared with the Data and Safety Monitoring Board (DSMB), operating under a charter. The DSMB will review the data to assess whether the study should be closed to future enrollment, and to determine whether blinding must be broken. While the DSMB is reviewing the data, recruitment may be temporarily halted until a decision is rendered.

#### 1.6 Schedule of Assessments

**Table 1. Schedule of Assessments**

| | | Treatment | | | | | Follow-up |
|---|---|---|---|---|---|---|---|
| **Activity** | **Screening Day -2 to 1[1]** | **Baseline Day 1** | **Days 2-7[2]** | **Day 8 ±2 days** | **Days 9-14[2]** | **Day 15 ±3 days** | **Day 28[12] ±7 days** |
| **Informed Consent** | x | | | | | | |
| **Confirmation Eligibility** | | x | | | | | |

| | | Treatment | | | | | Follow-up |
|---|---|---|---|---|---|---|---|
| **Activity** | **Screening Day -2 to 1[1]** | **Baseline Day 1** | **Days 2-7[2]** | **Day 8 ±2 days** | **Days 9-14[2]** | **Day 15 ±3 days** | **Day 28[12] ±7 days** |
| **Demographics & Medical History** | x | | | | | | |
| **Concomitant Meds** | x | x | x | x | x | x | x |
| **Physical Exam** | x | x | | | | | |
| **Weight and Height** | x | | | | | | |
| **Vital Signs** | x | x | x | x | x | x | |
| **SpO2 (pulse oximetry)** | x | x | x | x | x | x | |
| **12 Lead ECG** | | x[3] | | | | | |
| **Supplemental Oxygen Use Assessment** | x | x | x | x | x | x | x |
| **Need for Invasive or Non-invasive Ventilation Assessment** | | | x | x | x | x | x |
| **ICU Admission Assessment** | | | x | x | x | x | x |
| **CBC and Serum Chemistry** | x | x | | x | | x | |
| **ALT, AST** | x[4] | | | | | | |
| **Estimated GFR** | x | | | | | | |
| **SARS-CoV-2 PCR or rapid antigen** | x[5] | | | | | | |
| **hCG Test** | x [6] | | | | | | |
| **CT Scan** | | x [7] | x [8] | x [8] | x [8] | x [9] | |
| **Cytokine levels[10]** | | x | | x | | x | |
| **LDH** | | x | | x | | x | |
| **cf-DNA/NETs[11]** | | x | | x | | x | |
| **D-dimer** | | x | | x | | x | |
| **WHO Ordinal Assessment** | | x | x | x | x | x | x |
| **Dispense Study Drug** | | x | | x | | | |
| **Study Drug Accountability** | | | | x | | x | |
| **AE** | | | x | x | x | x | x |

[1] The screening and baseline visit can be combined if all eligibility criteria are met.
[2] Visits only if hospitalized.
[3] If the subject had ECG already performed for this hospitalization, it can be used.
[4] If ALT and AST levels are available from this hospitalization, they do not need to be repeated.
[5] SARS-CoV-2 PCR or rapid antigen results from a test performed within 7 days of screening can be used.
[6] hCG pregnancy test, either urine or blood to be performed in female subjects of childbearing potential
[7] If subject had CT scan performed within 3 days of enrollment this can be used.
[8] If CT scan occurs as standard of care, capture in the source documents.
[9] The Day 15 CT scan only occurs if the subject is in the hospital on this visit day.
[10] Cytokine levels (Cytokine IL-18, Cytokine TNF-α, Cytokine IL-1β, Cytokine IL-6, Cytokine IL-10, Cytokine IL-1RA, and Cytokine IL-8)
[11] Circulating neutrophil DNA / neutrophil extracellular traps (cf-DNA/NETs)
[12] May be conducted remotely.

#### 1.7 Study Schema

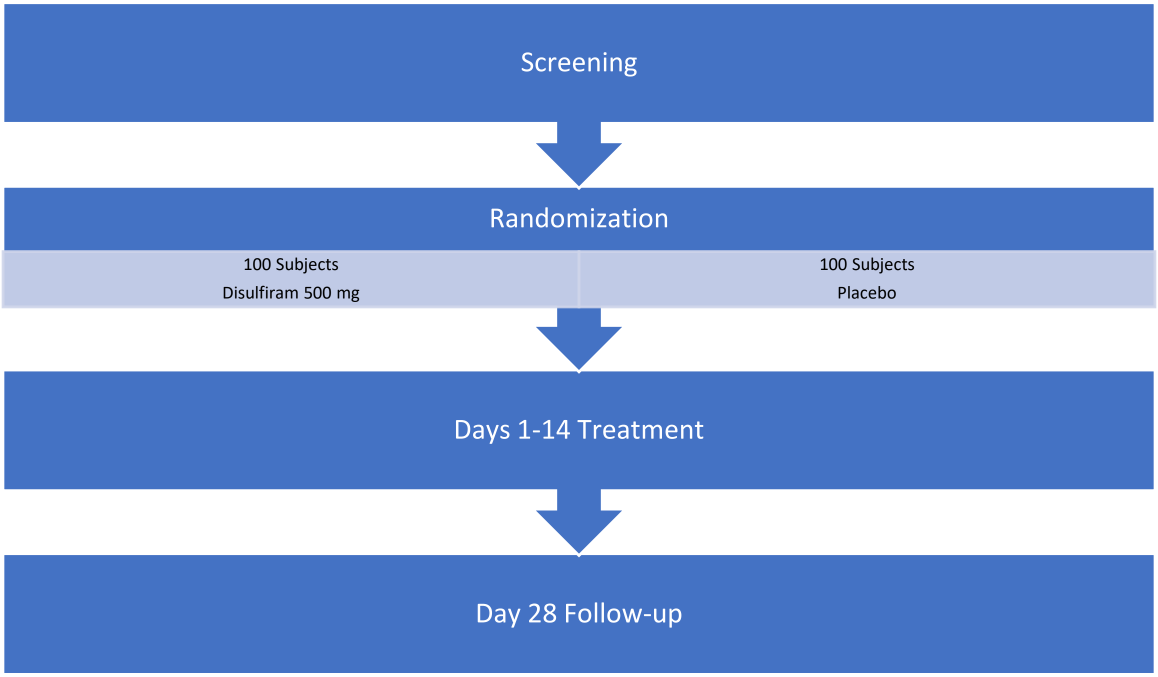

### 2 Introduction

#### 2.1 Study Rationale

COVID-19 is a respiratory disease caused by a novel coronavirus (SARS-CoV-2) and causes substantial morbidity and mortality[1]. At the start of the study there was no vaccine to prevent COVID-19 or infection with SARS-CoV-2 or therapeutic agent to treat COVID-19. Despite the start of the vaccination program in the past months, COVID-19 is still a potentially life-threatening disease in Brazil, for which no unequivocally proven therapy has emerged. This clinical trial is designed to evaluate disulfiram for the treatment of adult subjects hospitalized with COVID-19.

#### 2.2 Background

##### 2.2.1 Purpose of Study

Coronaviruses (CoVs) are positive-sense single-stranded enveloped RNA viruses, many of which are commonly found in humans and cause only mild symptoms. Over the past two decades, emerging pathogenic CoVs capable of causing life-threatening disease in humans and animals have been identified, namely Severe Acute Respiratory Syndrome coronavirus (SARS-CoV) and Middle East Respiratory Syndrome coronavirus (MERS-CoV).

Coronavirus Disease 2019 (COVID-19) emerged in Wuhan, China in late 2019 and is caused by Severe Acute Respiratory Syndrome coronavirus 2 (SARS-CoV-2), which is a highly infectious zoonotic virus.[2] Clinically, individuals with COVID-19 present with a myriad of symptoms, clinical course, and outcome. In order to develop effective treatments against COVID-19, it is necessary to understand the nature of the disease and the immunological response that occurs in these varying presentations. While the immune response - to pathogens has been well categorized (Type-1 Immunity, Type-2 Immunity, and Type-3 Immunity) and despite the numerous analyses that have been carried out to date, the correlation between an individual's immune response to a SARS-CoV-2 infection and their clinical outcome remain unclear.

Analyses of COVID-19 immunologic features have been carried out to identify differences between moderately and severely affected patients through the detection of immune biomarkers. Specifically, there has been interest in understanding the differing levels of cytokines, chemokines, and additional immune markers in these patient groups. A "core COVID-19 signature" of these markers was noted to exist in both moderately and severely affected patients. As reported by Lucas, et al.,[2] the majority of cytokines linked to this "signature" include IL-1α, IL-1β, IL-6, IL-10, IL-18, and TNF-α. These markers show an increased positive correlation in patients with severe disease and demonstrate a broad inflammatory response across type-1, type-2, and type-3 cytokines. Further, Lucas reported that individuals characterized with highly elevated proinflammatory cytokines were noted to develop a more severe disease course with many dying of COVID-19.[2]

McGonagle et al. similarly reported that an overactive immune response may be driving the COVID-19 related adult respiratory distress syndrome (ARDS).[3] McGonagle proposes that a loss of "front line" antiviral defense mechanisms may trigger a "second wave" of a more tissue-aggressive immunity, including exaggerated production of IL-6 with a secondary cytokine storm supervening with increased tissue damage.

Specific cell death pathways such as pyroptosis and netosis have been linked with the hyperinflammation and subsequent tissue damage observed in COVID-19 patients. The inflammasome-mediated pyroptosis is a type of programmed cell death characterized by gasdermin D-mediated membrane rupture, and spontaneous release of cytosolic contents including proinflammatory cytokines such as IL-1β and IL-18 and biomarkers such as the enzyme lactate dehydrogenase (LDH) into the extracellular spaces.[4, 5] These biomarkers have been identified in COVID-19 patients and correlate with disease severity. Release of such factors may prompt an immediate reaction from surrounding immune cells, attracting circulating immune cells such as monocytes and neutrophils and thus inducing a pyroptotic chain reaction that can ultimately lead to high levels of circulating proinflammatory cytokines defined as the "cytokine storm". Netosis is a unique form of cell death involving neutrophils that is characterized by the release of neutrophil extracellular traps (NETs) to the extracellular space in order to contain infections.[6] When dysregulated, NETs have the potential to propagate inflammation and create microvascular thrombosis. A feed-forward loop has been described between pyroptosis and netosis. Similarly to pyroptosis, NETs formation is gasdermin D-mediated.[7] Neutrophil infiltration in the lungs and the presence of cell free DNA (a biomarker of netosis) in the circulation have been observed in severe COVID-19 patients[8] and could be the origin of the fluid accumulation observed in the lungs of these patients.

Based upon these accounts that suggest a correlation between cytokine levels and disease course, significant interest has arisen in the medical and scientific communities to further understand how the inflammasome-mediated pyroptosis pathway and netosis may link to patient outcome, and how to predict or interrupt the process that appears to trigger vascular insults and tissue pathologies in severe COVID-19 patients. The elderly and those with comorbidities are reported as being the most susceptible to COVID-19, which may be due to a higher basal state of inflammation ("inflammaging") and a primed inflammasome pathway.[9] We believe that disulfiram could target the root cause of hyperinflammation, weakening the cytokine storm and therefore reducing the risk of progression to severe illness.

The Emergency Committee on COVID-19, convened by the WHO Director-General under the International Health Regulations (2005) (IHR), held its fourth meeting on 31 July 2020. In its statement following the meeting, it expressed "appreciation for WHO and partners' COVID-19 pandemic response efforts and highlighted the anticipated lengthy duration of this COVID-19 pandemic, noting the importance of sustained community, national, regional, and global response efforts."[10] Global efforts to evaluate novel antivirals and therapeutic strategies to treat COVID-19 have intensified. There is an understood urgent public health need for the rapid development of protective, predictive, and therapeutic interventions. In particular, the COVID-19 pandemic has brought focus to the need for therapeutics for the immune aging population.[11]

##### 2.2.2 Potential Therapeutics

Other than remdesivir, there are no drugs or other therapeutics to treat COVID-19. Current clinical management includes infection prevention and control measures and supportive care, including supplemental oxygen and mechanical ventilatory support, when indicated.

Disulfiram, an alcohol antagonist drug, has been used by clinicians since the 1950s to aid in the treatment of chronic alcoholism and is commercially approved for use in various countries, including the US. During normal human alcohol metabolism, ethanol is first converted into acetaldehyde and then to acetic acid. Disulfiram blocks the initial conversion process, resulting in a rapid increase of acetaldehyde, which is toxic and produces highly unpleasant side effects. This response is designed to deter alcohol intake.[12]

Disulfiram also has a potential for limiting the hyperinflammatory response associated with COVID-19. It has been documented during the COVID-19 pandemic that seemingly mild cases of COVID-19 can rapidly transition to severe cases that involve the lower lungs. This phenomenon may be related to the genesis of a "cytokine storm", characterized by an overproduction of immune cells and their activating compounds. This onslaught results in lung inflammation and fluid buildup that can lead to respiratory distress and serve as a site for secondary bacterial pneumonia, which increases the risk of mortality.[13] While this development of a “cytokine storm” is not fully understood, it is now believed to have been a major contributor to the mortality experienced in the 1918-1920 Spanish Flu and the worldwide H1N1 (swine flu) and H5N1 (bird flu) epidemics and pandemics, and is an urgent area of focus during the ongoing COVID-19 pandemic.

On a molecular level, the entry of an invading virus into a cell triggers inflammation and a cascade of events, including particular cell death pathways such as pyroptosis and netosis. Important cytokines, such as IL-1β and IL-18, are released during pyroptotic cell death.[5] Subsequently, a wave of local inflammation occurs that involves the increased secretion of proinflammatory cytokines and chemokines, such as IL-6, into the bloodstream. This secretion process attracts immune cells and for some patients, the response is dysfunctional and progresses to a cytokine storm and exacerbated netosis leading to fluid buildup in the lungs.[14] Individuals for whom this process has been fatal have been noted to have an increased level of cytokines, such as IL-6.[15] Research has assessed ways in which to interrupt the pyroptosis and netosis processes, including the use of disulfiram as a blocking mechanism to prohibit gasdermin D pore formation.[16] It is believed that disulfiram blocks the cellular gasdermin-D-NT fragments from creating pores in the cell wall. This mediation should be associated with a lessened increase of circulating cytokines in the bloodstream and may ultimately prevent the patient’s condition from progressing to a more severe state.

In addition to the potential immuno-inflammatory effect, disulfiram also has a potential for exerting a functional interference with the virus. With the emergence of illnesses such as Severe Acute Respiratory Syndrome coronavirus (SARS-CoV) in 2002, and the highly pathogenic human MERS-CoV outbreak ten years later with a fatality rate of 35%,[16] an urgent need developed to identify effective antiviral drugs for use in patients that were infected by these coronavirus pathogens. Disulfiram was studied by Lin, et al.[16] who reported that disulfiram can inhibit the papain-like proteases of MERS-CoV and SARS-CoV; is a noncompetitive inhibitor of MERS-CoV papain-like protease; is a competitive inhibitor of SARS-CoV papain-like protease; and is a slow-binding inhibitor that forms a covalent adduct at the active site of SARS-CoV papain-like protease. In addition to certain proteases being resistant to mutations, these proteases can play a major role in replication, which means that its inhibition slows down or even completely stops viral reproduction inside the body.

#### 2.3 Risk/Benefit

##### 2.3.1 Known Potential Benefits

This is a proof of concept, exploratory trial and for that reason, it is not possible to anticipate the potential benefit of disulfiram in the clinical outcomes of patients with moderate COVID-19. However, there is potential benefit to society from their participation in this study resulting from insights gained about the therapeutic agent under study. While there may not be benefits for an individual subject, there may be benefits to society if a safe, efficacious therapeutic agent can be identified during this global COVID-19 outbreak.

##### 2.3.2 Known Potential Risks

The potential risks of participating in this trial are those associated with having blood drawn, undergoing standard CT scan, possible reactions to disulfiram, and breach of confidentiality. Drawing blood may cause transient discomfort and fainting. Fainting is usually transient and managed by having the subject lie down and elevate his/her legs. Bruising at the blood collection sites may occur but can be prevented or lessened by applying pressure to the blood draw site for a few minutes after the blood is taken.

Disulfiram is a therapeutic drug used in adults for the management of ethanol abuse. The dosage in this adult and elderly population is typically 500 mg po once daily for up to two weeks, then reduced to 250 mg po once daily. With this dosing, there are labeled guidelines warning that the drug should be used with extreme caution in individuals with hepatic impairment as hepatic impairment and failure resulting in transplantation or death has been reported.[1]

As reported in the drug labeling, hypersensitivity and hematological disease have been reported during the administration of disulfiram. Hepatic disease or insufficiency may be exacerbated. Hepatic toxicity including hepatic failure resulting in transplantation or death have been reported. Severe and sometimes fatal hepatitis associated with disulfiram therapy may develop even after many months of therapy. Hepatic toxicity has occurred in patients with or without prior history of abnormal liver function. Disulfiram therapy can exacerbate certain conditions, such as diabetes mellitus, hypothyroidism, seizure disorder, cerebral damage, abnormal EEGs, renal disease, and multiple drug dependence, so the drug should be used with caution, if at all, in patients with these conditions. Disulfiram is contraindicated for use in patients with severe cardiac disease (e.g., coronary occlusion that usually results in acute myocardial infarction) or psychosis.

> **Disulfiram Labeling Warning Statement:**
> *WARNING: Disulfiram should never be administered to patients with ethanol intoxication and should never be administered to patients without their knowledge. Relatives of patients receiving the drug should be instructed about the precautions and risks associated with its use. Patients should be warned to avoid any ethanol ingestion including ethanol-containing foods or medications (e.g., vinegar, cough syrup, and sauces) while receiving disulfiram because a disulfiram-ethanol reaction can occur. The application of alcohol-containing linaments or lotions also may precipitate this reaction, and patients should be warned that reactions can occur up to 14 days following cessation of therapy. Disulfiram plus alcohol, even small amounts, produce flushing, throbbing in head and neck, throbbing headache, respiratory difficulty, nausea, copious vomiting, sweating, thirst, chest pain, palpitation, dyspnea, hyperventilation, tachycardia, hypotension, syncope, marked uneasiness, weakness, vertigo, blurred vision, and confusion. In severe reactions there may be respiratory depression, cardiovascular collapse, arrhythmias, myocardial infarction, acute congestive heart failure, unconsciousness, convulsions, and death. The intensity of the*

*reaction varies with each individual but is generally proportional to the amounts of disulfiram and alcohol ingested. Mild reactions may occur in the sensitive individual when the blood alcohol concentration is increased to as little as 5 to 10 mg/100 mL. Symptoms are fully developed at 50 mg/100 mL, and unconsciousness usually results when the blood alcohol level reaches 125 to 150 mg per 100 mL. The duration of the reaction varies from 30 to 60 minutes, to several hours in the more severe cases, or as long as there is alcohol in the blood.*

Adverse reactions associated with disulfiram have been well characterized and may include:

**Table 2. Adverse Reactions with Disulfiram**

| | |
|---|---|
| **Severe** | Heart failure / Delayed / Incidence not known<br>Seizures / Delayed / Incidence not known<br>Optic neuritis / Delayed / Incidence not known<br>Hepatic failure / Delayed / Incidence not known<br>Acute generalized exanthematous pustulosis (AGEP) / Delayed / Incidence not known |
| **Moderate** | Sinus tachycardia / Rapid / Incidence not known<br>Blurred vision / Early / Incidence not known<br>Respiratory depression / Rapid / Incidence not known<br>Dyspnea / Early / Incidence not known<br>Chest pain (unspecified) / Early / Incidence not known<br>Hypotension / Rapid / Incidence not known<br>Confusion / Early / Incidence not known<br>Palpitations / Early / Incidence not known<br>Atopic dermatitis / Delayed / Incidence not known<br>Peripheral neuropathy / Delayed / Incidence not known<br>Neuritis / Delayed / Incidence not known<br>Hepatitis / Delayed / Incidence not known<br>Cholestasis / Delayed / Incidence not known<br>Jaundice / Delayed / Incidence not known<br>Elevated hepatic enzymes / Delayed / Incidence not known<br>Impotence (erectile dysfunction) / Delayed / Incidence not known<br>Psychosis / Early / Incidence not known |
| **Mild** | Flushing / Rapid / Incidence not known<br>Weakness / Early / Incidence not known<br>Nausea / Early / Incidence not known<br>Vertigo / Early / Incidence not known<br>Syncope / Early / Incidence not known<br>Headache / Early / Incidence not known<br>Vomiting / Early / Incidence not known<br>Hyperhidrosis / Delayed / Incidence not known<br>Acne vulgaris / Delayed / Incidence not known |

| | |
|---|---|
| | Drowsiness / Early / Incidence not known<br>Dysgeusia / Early / Incidence not known<br>Fatigue / Early / Incidence not known |

Subjects with underlying chronic liver disease, as evidenced by a screening ALT (Alanine Transaminase) or AST (Aspartate aminotransferase) >3 times the upper limit of normal, will not be eligible for study enrollment. Any observed liver function-related abnormalities or possibly related AEs should be treated appropriately and followed to resolution. There are no labeled dose adjustments or specific warnings related to renal function; however, subjects with Stage 4 severe chronic kidney disease or requiring dialysis (estimated GFR < 30) are not eligible for study enrollment. Disulfiram should not be used with other drugs that have documented interactions with disulfiram. The study exclusion criteria list the drugs known to have documented interactions and subjects taking these drugs will not be eligible for study enrollment.

##### 2.3.3 Risks to Privacy

Subjects will be asked to provide Protected Health Information (PHI). All attempts will be made to keep this information confidential within the limits of the law. However, there is a chance that unauthorized persons will see the subject's PHI. Hard copies of study records will be kept in a locked file cabinet or maintained in a locked room at the participating clinical site. Electronic files will be password protected.

Only people who are involved in the conduct, oversight, or auditing of this study will be allowed access to the PHI that is collected.

Any publications from this study will not use information that will identify subjects by name. Organizations that may inspect and/or copy research records maintained at the participating site for quality assurance and data analysis include groups such as the IEC, designee, and the pertinent regulatory authorities.

##### 2.3.4 Assessment of Potential Risks and Benefits

Disulfiram is a generally well-documented medication. There are significant liver toxicities that have been reported. By excluding those with significant underlying liver and renal disease during the study, the risk to subjects can be reduced.

### 3 Objectives and Endpoints

The overall objective of the study is to evaluate the clinical outcomes, safety, and the effect of disulfiram on select biomarkers as compared to placebo in hospitalized subjects with moderate COVID-19.

| OBJECTIVES | ENDPOINTS (OUTCOME MEASURES) |
|---|---|
| **Primary** | |
| The primary objective is to assess the effect of disulfiram on clinical improvement. | To assess the time to clinical improvement, defined as the time from baseline to first post-baseline assessment with an improvement in WHO score of ≥1 point. |
| **Secondary** | |
| To assess the effect of disulfiram on clinical outcomes. | Key secondary:<br>1. To assess the mean number of days of supplemental oxygen (WHO Score ≥4).<br>2. To assess the time from baseline to discharge from the hospital.<br>3. To assess the percentage of subjects that are discharged by Day 8.<br>4. To assess the percentage of subjects that worsened 1 or more points on the WHO Ordinal Scale from baseline to any post baseline assessment through Day 28.<br>5. To assess the mean number of days of non-invasive ventilation or high flow oxygen devices or invasive mechanical ventilation (WHO Ordinal Scale 5 or 6) over the 28-day period.<br>6. To assess the mean number of days subjects were in the Intensive Care Unit (ICU).<br>7. To assess the percentage of subjects that were on non-invasive or high flow oxygen devices or invasive mechanical ventilation (WHO Ordinal Scale 5 or 6) over the 28-day period.<br>8. To assess the 28-day mortality. |
| To assess the effect of disulfiram on other measures of efficacy and various biomarkers. | Other Secondary:<br>1. To assess the mean change and percent change from baseline to Day 8 and Day 15 for Cytokine IL-18.<br>2. To assess the percentage of subjects requiring supplemental oxygen (WHO Score ≥4) by Day 8, 15, and 28.<br>3. To assess the percentage of subjects that are discharged by Day 15 and Day 28<br>4. To assess the percentage of subjects that worsened 1 or more points on the WHO Ordinal Scale from baseline through Day 8 and Day 15. |

| OBJECTIVES | ENDPOINTS (OUTCOME MEASURES) |
|---|---|
| | 5. To assess the percentage of subjects admitted to the Intensive Care Unit.<br>6. To assess the percentage of subjects that improved 1 or more points on the WHO Ordinal Scale from baseline to Day 8, 15, and 28.<br>7. To assess the change and percent change in neutrophil count from baseline to Day 8 and 15.<br>8. To assess the change and percent change in total lymphocyte count from baseline to Day 8 and 15.<br>9. To assess the change and percent change from baseline to Day 8 and 15 for neutrophil-derived circulating free DNA (cf-DNA/NETs).<br>10. To assess the mean change and percent change from baseline to Day 8 and 15 for:<br>a. Cytokine TNF-α<br>b. Cytokine IL-1β<br>c. Cytokine IL-1RA<br>d. Cytokine IL-6<br>e. Cytokine IL-8<br>f. Cytokine IL-10<br>g. Lactate dehydrogenase (LDH)<br>h. D-dimer<br>11. To assess the association between baseline and worst post-baseline WHO score. |
| Safety | Incidence of Adverse Events (AEs) and Serious Adverse Events (SAEs). Vital Signs and Laboratory data will be collected during the study period. |

### 4 Study Design

#### 4.1 Overall Design

This is a prospective, randomized, double-blind, placebo-controlled study to evaluate the safety and clinical outcomes of disulfiram administered to subjects diagnosed with moderate COVID-19. The multi-center study will be conducted in up to 8 clinical sites in Brazil. The study is a 2-arm comparison between disulfiram and placebo. Eligible subjects will be enrolled and randomized (1:1) to either receive disulfiram (study drug) or placebo, orally (po) once daily for fourteen (14) days. Given the severity of illness in COVID-19, there are no pre-specified study stopping rules for safety. The DSMB, which will operate under a charter, will review safety data.

#### 4.2 Scientific Rationale for Study Design

After the severe acute respiratory syndrome (SARS) coronavirus was identified in 2002 and caused a large global outbreak, there was an increased interest in the development of specific therapeutic agents. SARS-CoV case-patients were treated with corticosteroids, Type 1 IFN agents, convalescent plasma, ribavirin, and lopinavir or ritonavir, and, except for ribavirin, many of these agents have in vitro pre-clinical data that support their efficacy. Since the SARS outbreak, new therapeutic agents targeting viral entry proteins, proteases, polymerases, and methyltransferases have been tested; however, none have been shown to be efficacious in clinical trials.[17]

The ongoing COVID-19 pandemic has demonstrated increased risk to those with an aging immune system.

This study is designed to assess the safety and clinical outcomes of disulfiram as a potential therapeutic agent for patients with moderate COVID-19. The use of disulfiram and its impact on COVID-19 subjects will be assessed by analyzing the levels of signaling molecules in the blood of both active treatment and placebo subjects. These signaling molecules are known to regulate immunity, inflammation, and hematopoiesis and have been documented to be elevated in severe COVID-19 patients.[2] Based on analyses reported by Lucas, et. al.,[2] there appears to be a broad misfiring of immune components in COVID-19 patients, with early predictive markers and distinct correlations between types of immune responses among moderate and severe disease outcomes. The results suggest that the COVID-19 late-stage disease course may be driven primarily by host immune responses to SARS-CoV-2. As such, research for an effective cytokine blocking treatment and the impact on COVID-19 patients has become a priority. As reported by Lucas et al., cytokines, such as IL-18, were found to be elevated in patients with severe disease when compared to patients with moderate disease, at most time points that were analyzed. In concert with this potential relationship between cytokine levels and COVID-19 disease progression, the SPRING study is designed to assess the levels of Cytokine IL-18 in patients receiving disulfiram to assess the impact of the study drug on the patient’s cytokine levels.

#### 4.3 Justification for Dose

The dose of disulfiram used in the SPRING Study will be the same labeled dose for the drug’s use in the management of ethanol abuse, 500 mg administered orally, daily for two weeks.

Disulfiram has been studied in various areas of medicine, including oncology (i.e., breast cancer and melanoma), Lyme Disease, cocaine abuse, HIV (Human Immunodeficiency Virus), and others. In these studies, the dose of disulfiram administered to subjects ranged from 250 mg to 500 mg daily for up to 12 weeks duration.

The use of disulfiram in COVID-19 subjects is noted in ongoing clinical studies, including the “DISCO” study[18]. The DISCO study is being conducted in a total of sixty (60) COVID-19 PCR (+) subjects. As outlined in the study summary information available, subjects eligible to participate in the DISCO study will be administered 2,000 mg of disulfiram per day for three consecutive days and will be assessed for the impact on COVID-19 symptom severity, viral load, and biomarkers of inflammation over a total of 31 days.[18]

### 5 Study Population

Up to 200 subjects hospitalized with moderate COVID-19 who meet all eligibility criteria will be enrolled at up to 8 clinical sites in Brazil.

The estimated time from screening (Day -2 to 1) to end of study (Day 28 ± 7 days) for an individual subject is approximately 37 days.

Information regarding this trial may be provided to potential subjects who have previously participated in other trials conducted at the sites and to medical care providers who have cases of COVID-19 admitted to their hospital or in the referral area. Other forms and/or mechanisms of recruitment may also be used. The IEC will approve the recruitment process and all materials prior to use.

Subject Inclusion and Exclusion Criteria must be confirmed by a study clinician named on the delegation log.

Due to the evolving nature of COVID-19 disease, with the emergence of variant strains, the changes in the standard of care and the availability of vaccines, the inclusion criteria are not optimal anymore to target the population at risk to evolve to a severe disease.

At the start of the pandemic, age was the major risk factor associated with disease severity. Elderly is still a population at risk, but it seems like the new strains are also affecting younger people and the number of hospitalizations of adults under 50 years of age keeps increasing.

Also, as the vaccination schedule started from older adults to younger, the rate of hospitalized older adults is decreasing drastically and soon most of the population over 50 years of age will be vaccinated.

As the standard of care has also evolved since we first wrote this protocol, the clinical outcome of the patient has improved, and the overall use of mechanical ventilation has decreased. Over the course of the clinical trial, we have observed that subjects hospitalized with milder symptoms were discharged from the hospital in less than a week, and the number of subjects that either died or required mechanical ventilation was a lot lower than anticipated.

Taking all this into consideration, to enrich the subject population with those most likely to gain benefit from the investigational product, the inclusion criteria have been amended.

#### 5.1 Inclusion Criteria

Subjects may be enrolled in the study only if all the inclusion criteria are met.

1. Male and female subjects, age 35 or older.

2. Female subjects of childbearing potential must have a negative hCG pregnancy test (in urine or blood). Non-childbearing potential is defined as postmenopausal, with amenorrhea for ≥ 12 months; or ≥ 45 days post procedure for irreversible sterilization by hysterectomy, bilateral oophorectomy, or bilateral salpingectomy.

3. An International Ethics Committee (IEC) approved informed consent is signed and dated prior to any study-related activities.

4. Willing to abstain from any alcohol or alcoholic substances of any kind (including medications) within 24 hours prior to treatment and for 14 days after treatment concludes.

5. Have the ability to understand the requirements of the study and is willing to comply with all study procedures and visits.

6. Respiratory rate: ≤ 30 per minute.

7. Use supplemental $O_2$ via nasal cannula or equivalent.

8. Currently hospitalized ≤ 5 days.

9. PCR test or rapid antigen test confirming SARS-CoV-2. (If the subject had the PCR test performed within 7 days of screening, this can be used.) If the test is negative, a second test can be performed.

10. In the opinion of the investigator, able to participate in the study.

#### 5.2 Exclusion Criteria

Subjects may not be enrolled in the study if any of the exclusion criteria apply.

1. Admission into the Intensive Care Unit (ICU).
2. Clinically active Hepatitis.
3. ALT or AST > 3 times the upper limit of normal.
4. Need for invasive or non-invasive mechanical ventilation at screening or baseline visit.
5. Stage 4 severe chronic kidney disease or requiring dialysis or estimated GFR < 30.
6. Known allergy to disulfiram.

7. Treatment with any of the medications listed below within 7 days prior to the baseline visit[1]:

| | | | | |
|---|---|---|---|---|
| Amprenavir | Dronabinol | Hydantoins | Metronidazole | Ritonavir |
| Benznidazole | Dyphylline | Idelalisib | Naltrexone | Sertraline |
| Chloral Hydrate | Ethanol | Immunomodulatory drugs | Paclitaxel | Tinidazole |
| Cocaine | Ethotoin | Ixabepilone | Phenytoin | Tipranavir |
| Cyclosporine | Fosphenytoin | Lithium | Pimozide | Tranylcypromine |
| Dasabuvir | Guaifenesin | Mesoridazine | Pirfenidone | |

8. Participation in any other interventional trial within 30 days prior to the screening visit.
9. Active malignancy excluding basal cell carcinoma and squamous cell carcinoma, in situ cervical cancer or adenocarcinoma of the prostate with low or very low risk categories by NCCN criteria.
10. Any surgical or medical condition which in the opinion of the investigator may interfere with participation in the study or which may affect the outcome of the study.
11. Fully vaccinated for COVID-19 (number of doses as per manufacturer recommendation.

#### 5.3 Lifestyle Considerations

During this study, subjects are required to refrain from drinking alcohol or partaking in any alcoholic substances of any kind, including food (salad dressing, vinegars) and medications (for example cough syrup), within 24 hours prior to treatment and for 14 days after treatment concludes. Subjects should be cautioned about the use of hand sanitizers containing alcohol, alcohol swabs, or other products that contain alcohol.

#### 5.4 Screen Failures

After the screening evaluations have been completed, the investigator or designee is to review the inclusion/exclusion criteria and determine the subject's eligibility for the study.

The reason for ineligibility will be collected on screen failures. Subjects who are found to be ineligible will be told the reason for ineligibility.

Individuals who do not meet the criteria for participation in this study (screen failure) because of an abnormal laboratory finding may be rescreened once.

#### 5.5 Strategies for Recruitment and Retention

It is anticipated that patients with moderate COVID-19 will present to participating hospitals. Subject recruitment materials may be developed.

Patients that are confirmed to have PCR positive for SARS-CoV-2 will be assessed for eligibility.

Screening will begin with a brief discussion with the potential subject about the study with study staff. Information about the study will be presented to potential subjects (or legally authorized representative) and questions will be asked to determine potential eligibility. Some subjects will be excluded based on medical history. Screening procedures can begin only after signed informed consent is obtained.

### 6 STUDY PRODUCT

#### 6.1 STUDY PRODUCTS AND ADMINISTRATION

##### 6.1.1 Investigational Therapeutic and Matching Placebo

*Study Product Description*

Disulfiram (bis(diethylthiocarbamoyl) disulfide) is an alcohol antagonist drug. Disulfiram is designed to inhibit the enzyme acetaldehyde dehydrogenase. The drug was also found to be an inhibitor of papain-like proteases of MERS-CoV and SARS-CoV, suggesting a potential benefit. Specifically, this drug has recently been noted to be an inhibitor of gasdermin D. As such, disulfiram may have an antiviral effect and anti-inflammatory effect for COVID-19 subjects.

The supplied matching placebo will be given using the same dosing schedule.

At the initial study visit (Day 1 of the study) the patient will receive a supply of either the study drug, Disulfiram, or the placebo drug, depending on which group it has been randomized. This first supply will be sufficient for the first 7 days of treatment with 2 spare doses in case of a contamination or the loss of a dose in the first week. On the Day 8 visit, any remaining study drug will be collected, and the drug accountability will be performed, and the remaining study treatment identical to the first one (with the experimental drug, Disulfiram, or with the placebo drug) will be dispensed to the subject with sufficient quantity for the next 7 (seven) days, with two spare doses for eventual contamination or lost doses during the second week. On Day 15, any remaining study drug will be collected, and drug accountability performed.

*Dosing and Administration*

Subjects will be randomized to receive either the active product (disulfiram) or placebo.

Disulfiram will be dosed 500 mg daily for a total of 14 days of treatment. A matching placebo will be given at an equal volume using the same dosing schedule. Missed doses are not made up. There are no dose adjustments.

*Dose Administration Route*

The study treatment is intended for oral administration or enteral if required.

Study drug will be dispensed to the subjects. The subject will be responsible for administration daily and will be instructed to preserve the spare doses for counting at the end of the first and second weeks of treatment. During hospitalization, study medication will be dispensed daily by study staff.

If the subject is unable to swallow the capsule, the contents of the capsule can be dissolved in a liquid syrup. See the pharmacy manual for specific instructions.

*Dose Modifications*

Not applicable.

##### 6.1.2 Handling/Accountability/Destruction

*Acquisition*

The central pharmacy will be responsible for blinding and randomization of the study treatment. The PI or designee will notify the central pharmacy when a subject is eligible for treatment. The central pharmacy will deliver the first seven-day study treatment to the PI or designee to provide to the study subject. The Day 8 to 14 study treatment will be stored in the satellite pharmacy with the subject number, and it will be provided on the Day 8 visit to the PI or designee to provide to the study subject.

*Accountability*

The PI or designee is responsible for study product distribution and has ultimate responsibility for study product accountability, including maintaining complete records and documentation of study product receipt, accountability, dispensation, and final disposition of the study product(s).

All study product(s), whether administered or not, must be documented on the appropriate study product accountability log.

*Destruction*

After the study treatment period has ended, or as appropriate over the course of the study after study product accountability has been performed, disposition of unused and used active and placebo drug should occur as noted.

Study Product

- At the conclusion of the study, study product should be destroyed on-site following applicable site procedures or by the site's selected destruction vendor. Follow the site's standard operating procedure (SOP) for the destruction of study product when destroying used and unused items.
- A certificate of destruction should be provided to the Investigator if using an outside vendor. If following the site's SOP make a notation on the drug accountability log. Either should be retained in the Regulatory Binder once completed.

##### 6.1.3 Formulation, Appearance, Packaging, and Labeling

Disulfiram is a white to off-white, odorless, and almost tasteless powder.

The oral administration contains a total of 500mg disulfiram.

**Placebo to Match**
The supplied matching placebo is identical in physical appearance, taste, and odor to the study drug to retain blinding during the study.

Each of the study products will be labeled with a statement "Caution: Drug Limited by Regulatory Authority to Investigational Use."

##### 6.1.4 Product Storage

**Disulfiram**
Study product should be stored at room temperature between 15°C and 30°C.

**Placebo to Match**
Placebo should be stored between 15°C and 30°C.

##### 6.1.5 Preparation

Not applicable.

### 7 Measures to Minimize Bias: Randomization and Blinding

In order to minimize bias due to key baseline characteristics that can impact clinical outcomes the randomization will be stratified 1:1 to placebo or investigational product within each of the following 4 strata defined as Low Risk, Medium Risk, High Risk and Very High Risk, with expected worse clinical outcomes in each of the higher risk groups:

| | **Comorbidities:** hypertension (on any prescription anti-hypertension medication), diabetes (on oral or insulin injections for diabetes), BMI ≥ 35 | | | |
|---|---|---|---|---|
| **Age** | 0 | 1 | 2 | 3 |
| 35-59 | Low | Low | Low | Low |
| 60-69 | Low | Medium | Medium | Medium |
| 70-79 | Medium | High | High | High |
| ≥ 80 | High | Very High | Very High | Very High |

There is no requirement for a minimum number of subjects in each strata.

The study will randomize participants 1:1 to placebo or investigational product.

The randomization will be prepared and incorporated into the randomization module of the EDC system.

The PI or designee will notify the central pharmacy when a subject is eligible for randomization.

The central pharmacy will be responsible for providing medication for the subject with a randomization number on the dispensed medication.

Only required central pharmacy personnel will be unblinded to treatments allocated to each randomization number. All other site personnel, individuals from the CRO will remain blinded during the study until the database is locked and the study is unblinded.

#### 7.1 Study Intervention Compliance

Each dose of study product will be self-administered by the study subject or caregiver. During hospitalization, study medication will be dispensed daily only by study staff. The ICU team where there are patients participating in the study will also be informed and trained so that the patient is not exposed to any alcohol content substance, even in small concentrations.

#### 7.2 Concomitant Therapy

Standard of care.

##### 7.2.1 Rescue Medicine

Not Applicable.

##### 7.2.2 Non-Research Standard of Care

Not Applicable.

### 8 Study Intervention Discontinuation and Subject Discontinuation/Withdrawal

#### 8.1 Halting Criteria and Discontinuation of Study Intervention

##### 8.1.1 Study Halting for Safety

There are no pre-specified study stopping rules for safety. The DSMB, which will operate under a charter, will review safety data on a monthly basis and could recommend:

- The continuity of the study until the pre-defined number of patients has been reached
- The temporary interruption of the study for a better analysis of safety aspects
- The definitive discontinuation of the study for security reasons.

##### 8.1.2 Withdrawal from Randomized Treatment or from the Study

Subjects are free to withdraw from participation in the study at any time upon request, without any consequence. Subjects should be listed as having withdrawn consent only when they no longer wish to participate in the study and no longer authorize the Investigators to make efforts to continue to obtain their outcome data. Every effort should be made to encourage subjects to remain in the study for the duration of their planned outcome assessments. Subjects should be educated on the continued scientific importance of their data, even if they discontinue study drug. In the case of a subject's becoming lost to follow-up, attempts to contact the subject should be made and documented in the subject's source documents.

##### 8.1.3 Discontinuation of Study Drug

A subject in this clinical study may discontinue study drug for any of the following reasons:

- Request from the patient himself or his legal guardian in the event that it is impossible for the patient to do so to discontinue study drug.
- Occurrence of any medical condition or circumstance that exposes the subject to substantial risk and/or does not allow the subject to adhere to the requirements of the protocol.
- Any serious adverse event (SAE), clinically significant adverse event, severe laboratory abnormality, intercurrent illness, or other medical condition that indicates to the Investigator that continued participation is not in the best interest of the subject.
- Subject fails to comply with protocol requirements or study-related procedures.

Unless the subject withdraws consent, those who discontinue study drug early should remain in the study for further acquisition of endpoint measurements. The reason for subject discontinuation of study drug should be documented in the source documents and eCRF.

##### 8.1.4 Withdrawal of Subjects from the Study

A subject may be removed from the study for the following reasons post initial dosing; however, whenever possible the subject should be followed for safety evaluations per protocol:

- Subject withdraws consent or requests discontinuation from the study for any reason
- Death of the subject
- Termination of the study
- Lost to follow-up

Subjects who withdraw from this study or are lost to follow-up after signing the informed consent form (ICF) and administration of the study product, will not be replaced. The reason for subject discontinuation from the study will be recorded on the appropriate eCRF.

###### 8.1.5 Lost to Follow-up

A subject will be considered lost to follow-up if he or she fails to appear for a follow-up assessment and cannot be contacted with good effort. These efforts will be documented in the subject's record.

### 9 Study Assessments and Procedures

#### 9.1 Screening (Day -2 to 1)

The study and study procedures will be explained in detail to the subject and / or his/her legal representative(s). If willing to participate in the study, the subject will sign the informed consent. A unique subject number will be assigned, comprised of a two-digit site number and a three-digit sequential number (i.e., 01-001, 01-002, etc.) The inclusion and exclusion criteria will be reviewed. If the subject meets all the inclusion criteria and none of the exclusion criteria, they are eligible to participate in the study. Subjects that do not meet the inclusion/exclusion criteria are considered a screen failure.

The investigator will maintain a screening log to record details of all subjects screened and to confirm eligibility or record reasons for screen failure, as applicable.

The following information will be collected, and assessments performed for each subject:

- Age
- Sex
- Race (self-declared)
- Height (cm)
- Weight (kg)
- BMI (calculated by the site)
- Concomitant medications
- COVID vaccine (manufacturer, number of doses)
- Medical history- Including presence of hypertension or diabetes.
- Vital signs – heart rate, respiratory rate, blood pressure, and temperature
- Physical examination
- $SpO_2$ (All $SpO_2$ measurements in the protocol will be via pulse oximeter). It will be noted if the subject requires supplemental oxygen.
- CBC and serum chemistry
- ALT, AST and estimated GFR – If the subject had these tests performed within 5 days of screening, these results can be used.
- hCG pregnancy test (either blood or urine) in female subjects of childbearing potential. Non-childbearing potential is defined as postmenopausal, with amenorrhea for ≥ 12 months or ≥ 45 days post procedure for irreversible sterilization by hysterectomy, bilateral oophorectomy, or bilateral salpingectomy.

- PCR or rapid antigen test confirming SARS-CoV-2 - If the subject had a PCR test performed within 7 days of screening, these results can be used.

The overall eligibility of the subject to participate in the study will be assessed once all screening values are available.  Study subjects who qualify can have the screening and the baseline visit take place on the same day.

#### 9.2 Baseline (Day 1)

The eligibility of the subject to participate in the study will be confirmed. After collection of all medical assessments and information, the treatment arm will be randomly assigned.

The following assessments are performed for each subject:

- Recording of the concomitant medications, doses, and forms of use.
- Vital Signs – heart rate, respiratory rate, blood pressure, and temperature
- Physical examination
- 12 Lead ECG (If the subject had an ECG performed as part of the current hospitalization, this can be used.)
- Oxygen saturation will be measured by pulse oximetry ($SpO_2$).
- Chest CT scan – if the subject had a chest CT scan within 3 days of enrollment, this can be used, and a repeat scan is not needed.
- Laboratory blood tests - On day 1 of the study basic tests will be collected, which include the complete blood count and serum chemistry tests to assess liver and kidney function.  You will only need to collect these basic exams if your initial consultation occurs on a different day than the screening assessment. In addition to these basic tests, other blood tests will be performed to quantify the inflammatory reaction in your body, caused by COVID-19. Those tests includeCytokine Levels: IL-18, IL-6, TNF-α, IL-1β, IL-10, IL-8, IL-1RA; LDH; D-dimer
- Neutrophil DNA marker - cf-DNA/NET
- Determine the WHO Ordinal Scale score.

The WHO Ordinal Scale is an assessment of the current clinical status.  The scale is as follows:

1. Not hospitalized, no limitations on activities
2. Not hospitalized, limitations on activities
3. Hospitalized, not requiring supplemental oxygen
4. Hospitalized, requiring supplemental oxygen
5. Hospitalized, on non-invasive ventilation or high flow oxygen devices
6. Hospitalized, on invasive mechanical ventilation or ECMO
7. Death

Dispense study medication to the subject and update the drug accountability log.

The patient will be instructed on how to use the study medication. During hospitalization, study medication will be dispensed daily by study staff.

#### 9.3 Day 2-7 Visits

The following assessments are performed (only if subject is hospitalized subject):

- Concomitant medications
- Vital Signs – heart rate, respiratory rate, blood pressure, and temperature
- $SpO_2$
- Determine the WHO Ordinal Scale score.
- If a chest CT scan was performed as part of standard of care, capture the results.
- Note if the subject has been admitted to the ICU, requires invasive mechanical ventilation or non-invasive ventilation or supplemental oxygen.
- If the subject is unable to swallow the capsule, the contents of the capsule can be dissolved in a liquid syrup.  This applies for the entire duration of treatment.
- Record adverse events.

#### 9.4 Day 8 Visit (± 2 days)

This consultation will, whenever possible, be held on Day 8 of the study. However, for whatever reason, it can be performed from two days before (Day 6 of the study) to two days after (Day 10 of the study) Day 8.

The following assessments are performed for each subject:

- Recording of the concomitant medications, doses, and forms of use
- Vital Signs – heart rate, respiratory rate, blood pressure, and temperature
- Oxygen saturation will be measured by pulse oximetry ($SpO_2$)
- Laboratory blood tests – CBC and serum chemistry; cytokine levels: IL-18, IL-6, TNF-α, IL-1β, IL-10, IL-8, IL-1RA; D-dimer, LDH.
- Neutrophil DNA marker - cf-DNA/NET
- Determine the WHO Ordinal Scale score
- Note if the subject has been admitted to the ICU, requires invasive mechanical ventilation or non-invasive ventilation or supplemental oxygen.
- If a chest CT scan was performed as part of standard of care, record the results.
- Collect the remaining study treatment and update the drug accountability log.
- A new supply will be provided for the next 7 (seven) days of treatment (Day 8 to day 14 of the study). This new supply will have either the experimental drug, Disulfiram, or the placebo drug (without active substance), according to the initial randomization
- Record adverse events.

#### 9.5 Day 9-14 Visits

The following assessments are performed for each hospitalized subject:

- Concomitant medications
- Vital Signs – heart rate, respiratory rate, blood pressure, and temperature
- $SpO_2$
- Determine the WHO Ordinal Scale score.
- If a chest CT scan was performed as part of standard of care, capture the results.
- Note if the subject has been admitted to the ICU, requires invasive mechanical ventilation or non-invasive ventilation or supplemental oxygen.
- Record adverse events.

#### 9.6 Day 15 Visit (± 3 days)

This consultation will, whenever possible, be held on Day 15 of the study. However, for whatever reason, it can be done from three days before (Day 12 of the study) to three days after (Day 18 of the study) Day 15.

The following assessments are performed for each subject:

- Recording of the concomitant medications, doses, and forms of use.
- Vital Signs – heart rate, respiratory rate, blood pressure, and temperature
- Oxygen saturation will be measured by pulse oximetry $SpO_2$
- Laboratory blood tests – CBC and serum chemistry; cytokine levels: IL-18, IL-6, TNF-α, IL-1β, IL-10, IL-8, IL-1RA; D-dimer, LDH.
- Neutrophil DNA marker - cf-DNA/NET
- Chest CT scan (only for subjects that are in the hospital).
- Determine the WHO Ordinal Scale score.
- Note if the subject has been admitted to the ICU, requires invasive mechanical ventilation or non-invasive ventilation or supplemental oxygen.
- If a chest CT scan was performed as part of standard of care, record the results.
- Collect the remaining study treatment and update the drug accountability log.
- No new supplies will be provided. Treatment will have ended on Day 14 of the study.
- Record adverse events.

#### 9.7 Day 28 Visit (± 7 days)

This consultation will, whenever possible, be held on Day 28 of the study. However, for any reason, it can be performed from seven days before (Day 21 of the study) to seven days after (Day 35 of the study) Day 28. In addition, if for some reason the patient is unable to attend a face-to-face consultation within that time frame, this consultation can be done remotely.

The following assessments are performed for each subject:

- Recording of the concomitant medications
- Determine the WHO Ordinal Scale.

- Note if the subject has been admitted to the ICU for the preceding time period of day 15 to day 28, requires invasive mechanical ventilation or non-invasive ventilation or supplemental oxygen.
- Record adverse events.

#### 9.8 Early Termination Visit

If the subject is discontinued prior to the Day 15 visit, the assessments conducted on the ET visit will be the same as the Day 15 visit.

##### 9.8.1 Efficacy Assessments

**WHO Ordinal Scale**
The WHO Ordinal Scale is an assessment of the clinical status. Each day recorded the worse score for the time period will be recorded. The scale is as follows:

1. Not hospitalized, no limitations on activities
2. Not hospitalized, limitation on activities
3. Hospitalized, not requiring supplemental oxygen
4. Hospitalized, requiring supplemental oxygen
5. Hospitalized, on non-invasive ventilation or high flow oxygen devices
6. Hospitalized, on invasive mechanical ventilation or ECMO
7. Death

##### 9.8.2 Exploratory Assessments

Not applicable.

#### 9.9 Safety and Other Assessments

##### 9.9.1 Procedures to be Followed in the Event of Abnormal Laboratory Test Values or Abnormal Clinical Findings

Study procedures are specified in the Schedule of Assessments (SOA). A study physician licensed to make medical diagnoses and listed will be responsible for all trial-related medical decisions.

- Physical examination:
  A symptom-directed (targeted) physical examination will be performed to evaluate any possible adverse event. No physical exam is needed for routine visits.

- Clinical laboratory evaluations:
  - Fasting is not required before collection of laboratory samples.
  - Blood will be collected at the time points indicated in the SOA.
  - The procedure for sample collection and processing is outlined in the Laboratory Manual.

**Venipuncture Volumes**

At the screening: up to 38 mL

Day 1: up to 38 mL if the patient is eligible already in the screening consultation, there will be no need to repeat the basic exams.

Day 8: up to 38 mL

Day 15: up to 38 mL

#### 9.10 Adverse Events and Serious Adverse Events

##### 9.10.1 Definition of Adverse Event (AE)

Adverse Event means any untoward medical occurrence associated with the use of an intervention in humans, whether or not considered intervention related. An AE can therefore be any unfavorable and unintended sign (including an abnormal laboratory finding), symptom, or disease temporally associated with the use of the investigational product.

Any medical condition that is present at the time that the subject is screened will be considered as baseline and not reported as an AE. However, if the severity of any pre-existing medical condition increases, it should be recorded as an AE.

Given the nature of severity of the underlying illness, subjects will have many symptoms and abnormalities in vital signs and laboratory values. All Grade 3 and 4 AEs will be captured as AEs in this trial in the CRF.

##### 9.10.2 Definition of Serious Adverse Event (SAE)

An SAE is defined as “An AE or suspected adverse reaction is considered serious if, in the view of the investigator it results in any of the following outcomes:

- Death
- a life-threatening AE
- inpatient hospitalization or prolongation of existing hospitalization
- a persistent or significant incapacity or substantial disruption of the ability to conduct normal life functions
- or a congenital anomaly/birth defect

Important medical events that may not result in death, be life-threatening, or require hospitalization may be considered serious when, based upon appropriate medical judgment, they may jeopardize the subject and may require medical or surgical intervention to prevent one of the outcomes listed in this definition. Examples of such medical events include allergic bronchospasm requiring intensive treatment in an emergency room or at home, blood dyscrasias or convulsions that do not result in inpatient hospitalization, or the development of drug dependency or drug abuse.

“Life-threatening” refers to an AE that at occurrence represents an immediate risk of death to a subject. An event that may cause death if it occurs in a more severe form is not considered life-threatening. Similarly, a hospital admission for an elective procedure is not considered a SAE.

All SAEs, as with any AE, will be assessed for severity and relationship to study intervention.

All SAEs will be recorded on the appropriate SAE CRF.

All SAEs will be followed through to resolution or stabilization by a licensed study physician.

All SAEs will be reviewed and evaluated and will be sent to the IEC.

##### 9.10.3 Suspected Unexpected Serious Adverse Reactions (SUSAR)

A SUSAR is any SAE where a causal relationship with the study product is at least reasonably possible but is not listed in the Investigator Brochure (IB), Package Insert, and/or Summary of Product Characteristics.

##### 9.10.4 Classification of an Adverse Event

The determination of seriousness, severity, and causality will be made by an on-site investigator who is qualified (licensed) to diagnose AE information, provide a medical evaluation of AEs, and classify AEs based upon medical judgment. This includes but is not limited to physicians, physician assistants, and nurse practitioners.

**Severity of Adverse Events**
All AEs and SAEs will be assessed for severity, according to the Division of AIDS (DAIDS) Table for Grading the Severity of Adult and Pediatric Adverse Events, version 2.1 (July 2017).

For AEs not included in the protocol-defined grading system, the following guidelines will be used to describe severity.

- Mild (Grade 1): Events that are usually transient and may require only minimal or no treatment or therapeutic intervention and generally do not interfere with the subject’s usual activities of daily living.
- Moderate (Grade 2): Events that are usually alleviated with additional specific therapeutic intervention. The event interferes with usual activities of daily living, causing discomfort but poses no significant or permanent risk of harm to the research subject.
- Severe (Grade 3): Events interrupt usual activities of daily living, or significantly affects clinical status, or may require intensive therapeutic intervention. Severe events are usually incapacitating.
- Severe (Grade 4): Events that are potentially life-threatening.

AEs characterized as intermittent require documentation of onset and duration of each episode. The start and stop Duration of each reported AE will be recorded on the appropriate CRF. Changes in the severity of an AE will be documented to allow an assessment of the duration of the event at each level of intensity.

**Relationship to Study Intervention**
For each reported adverse reaction, the PI or designee must assess the relationship of the event to the study product using the following guideline:

- Related – The AE is known to occur with the study intervention, there is a reasonable possibility that the study intervention caused the AE, or there is a temporal relationship between the study intervention and event. Reasonable possibility means that there is evidence to suggest a causal relationship between the study intervention and the AE.
- Not Related – There is not a reasonable possibility that the administration of the study intervention caused the event, there is no temporal relationship between the study intervention and event onset, or an alternate etiology has been established.

##### 9.10.5 Time Period and Frequency for Event Assessment and Follow-up

For this study, all Grade 3 and 4 AEs and all SAEs occurring from the time the informed consent is signed through the Day 28 (end of study) visit will be documented, recorded, and reported.

**Investigators Reporting of AEs**
Information on all AEs should be recorded on the appropriate CRF. All clearly related signs, symptoms, and results of diagnostic procedures performed because of an AE should be grouped together and recorded as a single diagnosis. If the AE is a laboratory abnormality that is part of a clinical condition or syndrome, it should be recorded as the syndrome or diagnosis rather than the individual laboratory abnormality. Each AE will also be described in terms of duration (start and stop date), severity, association with the study product, action(s) taken, and outcome.

##### 9.10.6 Serious Adverse Event Reporting

**Investigators Reporting of SAEs**
Any AE that meets a protocol-defined criterion as a SAE must be submitted immediately (within 24 hours of site awareness) on an SAE form to the IEC system, through the quickest and most efficient channel of communication.

Other supporting documentation of the event may be requested by the designated IEC and should be provided as soon as possible.

At any time after completion of the study, if the site PI or appropriate sub-investigator becomes aware of an SAE, the site PI or appropriate sub-investigator will report the event to the designated IEC.

**Regulatory Reporting of SAEs**
Any unexpected fatal or life-threatening suspected adverse reaction will be reported to the regulatory authority as soon as possible, but in no case later than 7 calendar days after the investigator's initial receipt of the information. Upon request from regulatory authority, the investigator will submit any additional data or information that the agency deems necessary, as soon as possible, but in no case later than 15 calendar days after receiving the request.

Sites may have additional local reporting requirements (to the IEC and/or national regulatory authority).

##### 9.10.7 Reporting Events to Subjects

Subjects will be informed of any severe AEs or SAEs that occur as part of their participation in this trial.

##### 9.10.8 Reporting of Pregnancy

Pregnancy is not an AE. However, any pregnancy that occurs during study participation should be reported to the IRB on the appropriate CRF. Pregnancy should be followed to outcome.

#### 9.11 Unanticipated Problems

##### 9.11.1 Definition of Unanticipated Problems (UP)

An Unanticipated Problem is any event, incident, experience, or outcome that meets the following criteria:

- Unexpected in terms of nature, severity, or frequency given (a) the research procedures that are described in the protocol-related documents, such as the CEP/CONEP -approved research protocol and informed consent document; and (b) the characteristics of the subject population being studied;
- Related or possibly related to participation in the research ("possibly related" means there is a reasonable possibility that the incident, experience, or outcome may have been caused by the procedures involved in the research); and
- Suggests that the research places subjects or others at a greater risk of harm (including physical, psychological, economic, or social harm) than was previously known or recognized.

##### 9.11.2 Unanticipated Problem Reporting

To satisfy the requirement for prompt reporting, unanticipated problems (UP) will be reported using the following timeline:

- UPs that are SAEs will be reported to the CEP/CONEP within 24 hours of the investigator becoming aware of the event per the above describe SAE reporting process.

- Any other UP will be reported to the CEP/CONEP within 3 days of the investigator becoming aware of the problem.

##### 9.11.3 Reporting Unanticipated Problems to Subjects

Subjects will be informed of any unanticipated problems that occur as part of their participation in this trial.

### 10 Statistical Considerations

This protocol section summarizes the planned statistical analyses. A separate Statistical Analysis Plan (SAP) will be finalized prior to the database being locked and the study unblinded.

The SAP will serve as a complement to the study protocol and will supersede the protocol in case of differences. Any changes to the SAP will be documented prior to database lock.

#### 10.1 Statistical Hypothesis

The null hypothesis for the primary endpoint assumes no difference in Kaplan-Meier curves of time to improvement in the active treatment group (disulfiram) compared to the placebo group.

The alternative hypothesis is that there is a difference in KM curves between time to improvement in the active treatment group compared to the placebo group. Both hypotheses are represented below:

H0: The two time to improvement KM curves are identical (or St1 = St2)
HA: The two time to improvement KM curves are not identical (or St1 ≠ St2)

Where St1 is the KM curve for time to improvement of ≥1 point in WHO score from Baseline to Day 28 visit for subjects randomized to Disulfiram (Active) treatment group and St2 is the KM curve for time to improvement of ≥1 point in WHO score from Baseline to Day 28 visit for subjects randomized to the placebo group.

#### 10.2 Sample Size Estimation

Since the efficacy of disulfiram on COVID-19 infected patients is not known, the sample size for this study is not based on any formal calculations.

This study will provide adequate data to assess potential benefits as measured clinical improvement and key secondary clinical outcomes and various biomarkers.

#### 10.3 Population for Analyses

| | |
|---|---|
| **Modified Intent-to-Treat (mITT) population** | The mITT population will include all randomized subjects who receive a dose of study drug and have WHO score measured both at the baseline visit and at least one post-baseline timepoint. The subjects will be grouped as randomized irrespective of the treatment received. The mITT population will be used for all efficacy analyses. |
| **Per Protocol (PP) population** | The per protocol population is a subset of the mITT population and includes all subjects who did not have any important protocol deviations significantly impacting efficacy assessment, the participant's rights, or their safety. For this population, subjects will be grouped based on treatment they received. The per protocol population will be used for sensitivity analysis of the primary efficacy endpoint and may also be used for selected secondary endpoints. |
| **Safety Population (SP)** | The safety population will consist of all participants randomized and receiving at least one dose of study drug. The participants will be grouped according to the treatment actually received. The safety population will be used for safety analyses. |

#### 10.4 Statistical Analyses

##### 10.4.1 General Approach

All data collected in the study will be presented in Individual Data listings. All listings will include the Subject ID and treatment group. Where applicable, listings will also include visit number, visit date, and days relative to the initiation of treatment. Individual Subject Data Listings, as a minimum, are sorted by Subject ID and Visit, where applicable.

When missing or partial dates are imputed, the listings will display both the missing or partial dates as well as the imputed dates. Any data derived for analyses will also be included with the raw data in the listings and flagged as derived data.

Summary Tables will present data to compare the two treatment groups.

Continuous variables (e.g., age) are summarized using descriptive statistics (the number of participants with available data, the mean, standard deviation (SD), first quartile (Q1), median, third quartile (Q3), minimum and maximum). Categorical variables (e.g., race) are summarized using counts and percentages. Percentages are calculated using the total participants per treatment group with available data (overall or within relevant subgroup when appropriate). Summaries for discrete variables will include frequencies and percentages. Denominators used for computation of percentages will be indicated in footnotes on tables.

The baseline value for efficacy and safety is the last non-missing observed data prior to the first dose of study drug.

All analyses will be conducted using SAS 9.4 or higher.

##### 10.4.2 Subject Disposition

The number of subjects signing informed consent and subjects failing screening will be summarized. For subjects randomized, the number of subjects treated, completed through Day 15 and 28 and discontinued from the study will be tabulated by treatment group. A subject is considered to have completed the study if they complete the Day 28 visit. For subjects who discontinue the number of percentage of subjects that discontinue by reason for discontinuations will be tabulated.

##### 10.4.3 Baseline Descriptive Statistics

Demographics (Age, Sex, Race, etc.), and other baseline characteristics will be summarized by treatment group.

##### 10.4.4 Prior and Concomitant Medications

All medications (e.g., both prior and concomitant) will be coded using the WHO Drug Dictionary, Version September 1, 2020, B3. The incidence of medications will be summarized by therapeutic class and preferred term. Participants are counted only once in each therapeutic class category, and only once in each preferred term category. Prior medications will include all medications taken prior to a participant taking the first dose of study drug treatment. Any medication given at least once on or after the date of first day of study drug treatment will be defined as a concomitant medication.

##### 10.4.5 Protocol Deviations

As per ICH E3 guideline Section 10.2, all important deviations related to study inclusion or exclusion criteria, conduct of the trial, participant management or participant assessment will be summarized by site and grouped into different categories, such as:

- those who entered the study even though they did not satisfy the entry criteria;
- those who developed withdrawal criteria during the study but were not withdrawn;
- those who received the wrong treatment or incorrect dose;
- those who received an excluded concomitant treatment.

Protocol Deviations will be categorized as important or not.

A summary Table will compare important protocol deviations between the two treatment groups.

##### 10.4.6 Analysis of the Primary Efficacy Endpoint

The primary efficacy endpoint will be analyzed using a log rank test by comparing Kaplan-Meier (KM) curves of time to improvement of ≥ 1 point in WHO Score from baseline through Day 28 between the active treatment group (disulfiram) and the placebo group. Handling of missing data will be defined in the SAP and finalized prior to database lock, if required.

##### 10.4.7 Analysis of the Secondary Endpoint(s)

In order to control for multiplicity, testing of the statistical hypothesis for secondary efficacy endpoints will be conducted hierarchically following test of the primary efficacy endpoint. Each test will be at a type I error level ≤0.05, two-sided. As soon as a test yields a p-value > 0.05 no further tests following it will be considered statistically significant adjusting for multiplicity, however p-values will be presented and considered nominal (unadjusted for multiplicity).

Continuous secondary efficacy endpoints will be analyzed either similar to the primary efficacy endpoint as described above or using an analysis of covariance (ANCOVA) adjusted for key prognostics variables for post baseline visits between treatment groups to assess the difference.

Categorical endpoints will compare the percentage of responders between the active and placebo groups using logistic regression adjusted for key prognostic variables. If there are less than 5 subjects in any cell, a Fisher's exact test will be used.

The exact model for ANCOVA and logistics regression will be defined in the SAP and finalized prior to database lock.

##### 10.4.8 Safety Analyses

The safety analysis will be conducted on the safety population. AEs will be coded according to the Medical Dictionary for Regulatory Activities (MedDRA) dictionary Version 23.1. The Treatment Emergent Adverse Events (TEAEs) will be summarized by treatment group, system organ class and preferred term. Further summaries will be done by seriousness, severity, and relationship to study drug. Vital signs, and laboratory tests at baseline at each post-baseline visit, and changes from baseline to each of the post-baseline visits will be summarized with descriptive statistics. Shift tables for laboratory tests based on a classification of values as low, normal, or high with respect to the reference range will be summarized.

All safety data will be summarized with descriptive statistics with no hypothesis testing.

##### 10.4.9 Planned Interim and Early Analyses

None planned.

###### 10.4.10 Subgroup Analyses

Subgroup analyses for the primary and key secondary outcomes will evaluate the treatment effect across the following subgroups: age, sex, and comorbidities. A forest plot will display confidence intervals across subgroups. Interaction tests will be conducted to determine whether the effect of treatment varies by subgroup. Additional subgroups might be added in addition to above and will be defined in SAP and finalized prior to database lock.

###### 10.4.11 Exploratory Analyses

Exploratory analyses will be conducted as deemed appropriate.

### 11 Supporting Documentation and Operational Considerations

#### 11.1 Regulatory, Ethical, and Study Oversight Considerations

This study will be conducted in conformity with the principles set forth in The Belmont Report: Ethical Principles and Guidelines for the Protection of Human Subjects of Research.

The CEP/CONEP will review and approve this protocol, associated informed consent document, recruitment material, and handouts or surveys intended for the subjects, prior to the recruitment, screening, and enrollment of subjects. The local IEC may have additional local regulations.

Any amendments to the protocol or consent materials will be approved by the CEP/CONEP before they are implemented. CEP/CONEP review and approval will occur at least annually throughout the duration of the study. The investigator will notify the CEP/CONEP of deviations from the protocol and SAEs, as applicable to the CEP/CONEP policy.

The Investigator must receive the documentation that verifies CEP/CONEP approval for this protocol, informed consent documents, and associated documents prior to the recruitment, screening, and enrollment of subjects, and any CEP/CONEP approvals for continuing review or amendments.

##### 11.1.1 Informed Consent Process

Informed consent is a process that is initiated prior to an individual agreeing to participate in a study and continuing throughout the individual’s study participation. Investigators or designated research staff will obtain a subject’s informed consent in accordance with national and local regulatory requirements.

Subjects will receive a concise and focused presentation of key information about the clinical trial, verbally and with a written consent form. The key information about the study will be organized and presented in lay terminology and language that facilitates understanding why one might or might not want to participate.

ICFs will be CEP/CONEP approved, and subjects will be asked to read and review the consent form. Subjects (or legally authorize representatives) must sign the ICF prior to starting any study procedures being done specifically for this trial. Once signed, a copy of the ICF will be given to the subject for their records.

New information will be communicated by the site PI to subjects who consent to participate in this trial in accordance with IEC requirements. The informed consent document will be updated, and subjects will be re-consented per IEC requirements, if necessary.

**Other Informed Consent Procedures**
Subjects will be asked for consent to collect additional blood, the use of residual specimens, and samples for secondary research. Extra blood will be drawn for secondary research during each visit when a study blood samples are obtained.

The stored samples will be labeled to maintain confidentiality.

Samples designated for secondary research use may be used for understanding the SARS-CoV-2 infection, the immune response to this infection, and the effect of therapeutics on these factors.

Samples will not be sold for commercial profit. Although the results of any future research may be patentable or have commercial profit, subjects will have no legal or financial interest in any commercial development resulting from any future research.

There are no direct benefits to the subject for extra specimens collected or from the secondary research. No results from secondary research will be entered into the subject's medical record. Incidental findings will not be shared with the subject, including medically actionable incidental findings, unless required by law.

Subjects may withdraw permission to use samples for secondary use at any time. They will need to contact the study site and the samples will be removed from the study repository after this study is completed and documentation will be completed that outlines the reason for withdrawal of permission for secondary use of samples.

##### 11.1.2 Study Termination and Closure

In Section 8, Study Intervention Discontinuation and Subject Discontinuation/Withdrawal, describes the temporary halting of the study.

This study may be prematurely terminated if there is sufficient reasonable cause, including but not limited to:

- Determination of unexpected, significant, or unacceptable risk to subjects
- Insufficient compliance to protocol requirements

- Data that are not sufficiently complete and/or not evaluable
- Regulatory authorities

If the study is prematurely terminated, the site PI will promptly inform study subjects and the CEP/CONEP, as applicable. The site PI will assure appropriate follow-up for the subjects, as necessary.

##### 11.1.3 Confidentiality and Privacy

Subject confidentiality is strictly held in trust by the participating investigators, their staff, and the sponsor(s), if applicable, and their agents. This confidentiality is extended to cover clinical information relating to subjects, test results of biological samples and genetic tests, and all other information generated during participation in the study. No identifiable information concerning subjects in the study will be released to any unauthorized third party. Subject confidentiality will be maintained when study results are published or discussed in conferences.

The study monitor, other authorized representatives, representatives of the IEC, and/or regulatory agencies may inspect all documents and records required to be maintained by the investigator, including but not limited to, medical records (office, clinic, or hospital) and pharmacy records for the subjects in this study. The clinical study site will permit access to such records.

All source records including electronic data will be stored in secured systems in accordance with institutional policies and federal regulations.

All study data and research specimens that leave the site (including any electronic transmission of data) will be identified only by a coded number that is linked to a subject through a code key maintained at the clinical site. Names or readily identifying information will not be released unless the Investigator approves and it aligns with the consent form, or according to laws for required reporting.

##### 11.1.4 Secondary Use of Stored Specimens and Data

Secondary Human Subject Research is the re-use of identifiable data or identifiable biospecimens that were collected from some other ‘‘primary’’ or ‘‘initial’’ activity, such as the data and samples collected in this protocol. Out of the 38 mL of blood drawn, 5 mL of blood will be separated into peripheral blood mononuclear cells (PBMCs) and plasma and will be stored in a biorepository for up to 10 years for possible further analysis to measure Anti-SARS-CoV-2 Antibody, Disulfiram, and Disulfiram metabolites. In case these samples are used, they will be used exclusively for research purposes to better characterize biomarkers associated with either improvement or worsening of COVID, and better understand the mechanism by which disulfiram helps to improve COVID infection. These possible future tests will be run in Brazil whenever possible, but in case of this not being possible, they will be run in the United States.

Each sample will be labeled only with a unique subject number to protect subject confidentiality. Secondary research with coded samples and data may occur, however, subject confidentiality will be maintained as described for this protocol. An IEC review of the secondary research using coded specimens is required.

The subject's decision can be changed at any time by notifying the study doctors or nurses in writing. If the subject subsequently changes his/her decision, the samples will be destroyed if the samples have not been used for research or released for a specific research project.

##### 11.1.5 Data Sharing and Secondary Research

Data from this study may be used for secondary research. All the individual subject data collected during the study will be made available after de-identification. The SAP and Analytic Code will also be made available. This data will be available immediately following publication, with no end date.

#### 11.2 Key Roles and Study Governance

The study is physician-initiated study. Decisions related to the study will be made by the involved physician.

##### 11.2.1 Safety Oversight

**Protocol Oversight**
Given the severity of illness in COVID-19, there are no pre-specified study stopping rules for safety. Serious Adverse Events will be shared with the Data and Safety Monitoring Board (DSMB), operating under a charter. The DSMB will review the data to assess whether the study should be closed to future enrollment, and to determine whether blinding must be broken. While the DSMB is reviewing the data, recruitment may be temporarily halted until a decision is rendered.

**Data Safety Monitoring Committee**
Safety oversight will be conducted by a DSMB that is an independent group of experts that monitors subject safety. The DSMB members will be separate and independent of study personnel participating in this trial and should not have scientific, financial, or other conflict of interest related to this trial. Details of the composition, responsibilities, process, and frequency of review of safety data as well as meetings will be in the Data and Safety Monitoring Board (DMSB) charter.

##### 11.2.2 Data handling and Record Keeping

**Data Collection and Management Responsibilities**
Data collection is the responsibility of the study personnel at the participating clinical study site under the supervision of the site PI. The site PI must maintain complete and accurate source documentation.

Clinical research data from source documentation (including, but not limited to, AEs, SAEs, concomitant medications, medical history, clinical laboratory data, CT scan, etc.) will be entered by the clinical study site into eCRFs (electronic Case Report Forms). The data system includes password protection. AEs and concomitant medications will be coded according to the most current versions of MedDRA and WHODrug, respectively.

AEs will be coded according to the MedDRA dictionary version 23.0 or higher.

**Study Record Retention**

Study-related records, including the regulatory file, study product accountability records, consent forms, subject source documents and electronic records should be maintained for a period of 2 years following the date a marketing application is approved for the investigational product for the indication for which it is being investigated; or, if no application is to be filed or if the application is not approved for such indication, until 2 years after the investigation is discontinued. These documents should be retained for a longer period, however, if required by local policies or regulations. No records will be destroyed without the written consent of the study PI. Consent forms with specimen retention linked to identifiable specimens will be maintained for as long as the specimens remain in identifiable format, and a minimum of three years after use of the identifiable specimens in non-exempt human subject research.

**Source Records**

Source data are all information in original records (and certified copies of original records) of clinical findings, observations, or other activities in a clinical trial necessary for the reconstruction and evaluation of the trial. Source data should be attributable, legible, contemporaneous, original, accurate, and complete. Each participating site will maintain appropriate medical and research records for this study, in compliance with regulatory and institutional requirements. Data recorded in the eCRF derived from source documents should be consistent with the data recorded on the source documents.

Interview of subjects is sufficient for obtaining medical history. Solicitation of medical records from the subject’s primary care provider are not required.

##### 11.2.3 Protocol Deviations

A protocol deviation is any noncompliance with the clinical trial protocol.

The noncompliance may be either on the part of the subject, the investigator, or the study site staff. Following a deviation(s), corrective actions should be developed by the site and implemented promptly. All individual protocol deviations will be addressed in subject study records.

It is the responsibility of the site PI and personnel to use continuous vigilance to identify and report deviations. All deviations must be recorded on the protocol deviation log. Protocol deviations should be sent to the local IEC per their guidelines. The site PI and personnel are responsible for knowing and adhering to their IEC requirements. A completed copy of the Protocol Deviation Form must be maintained in the Regulatory File, as well as in the subject's chart if the deviation is subject specific.

##### 11.2.4 Publication and Data Sharing Policy

To avoid premature release of data, this Protocol specifies that efficacy data that has not yet been completed due to insufficient enrollment should not be released.

##### 11.2.5 Publication

Following completion of the study, the PI may publish the results of this research in a scientific journal.

##### 11.2.6 Conflict of Interest Policy

The independence of this study from any actual or perceived influence, such as by the pharmaceutical industry, is critical. Therefore, any actual conflict of interest of persons who have a role in the design, conduct, analysis, publication, or any aspect of this study will be disclosed and managed. Furthermore, persons who have a perceived conflict of interest will be required to have such conflicts managed in a way that is appropriate to their participation in the design and conduct of this study.

### 12 Additional Considerations

#### 12.1 Research Related Injuries

For any potential research related injury, the site PI or designee will assess the subject. Study personnel will try to reduce, control, and treat any complications from this study. Immediate medical treatment may be provided by the participating study site.

As needed, referrals to appropriate health care facilities will be provided to the subject. The site PI should then determine if an injury occurred as a direct result of the tests or treatments that are done for this study.

If it is determined by the participating site PI that an injury occurred to a subject as a direct result of the tests or treatments that are done for this study, then referrals to appropriate health care facilities will be provided to the subject.

Study personnel will try to reduce, control, and treat any complications from this study. Immediate medical treatment may be provided by the participating site, such as giving emergency medications to stop immediate allergic reactions.

### 13 ABBREVIATIONS

| | |
|---|---|
| AE | Adverse Event |
| AGEP | Acute Generalized Exanthematous Pustulosis |
| AIDS | Acquired Immunodeficiency Syndrome |
| ALT | Alanine Transaminase |
| ARDS | Adult Respiratory Distress Syndrome |
| AST | Aspartate Transaminase |
| BMI | Body Mass Index |
| CBC | Complete Blood Count |
| Cf-DNA/NET | Neutrophil-derived circulating free DNA |
| CoVs | Coronavirus |
| CT | Computed Tomography |
| CRF | Case Report Form |
| CSR | Clinical Study Report |
| DNA | Deoxyribonucleic Acid |
| DSMB | Data and Safety Monitoring Board |
| ECG | Electrocardiogram |
| E-CRFs | Electronic Case Report Forms |
| EEG | Electroencephalogram |
| ECMO | Extracorporeal Membrane Oxygenation |
| FDA | Food and Drug Administration |
| GCP | Good Clinical Practice |
| GFR | Glomerular Filtration Rate |
| HIV | Human Immunodeficiency Virus |
| IB | Investigator’s Brochure |
| ICF | Informed Consent Form |
| ICH | International Council for Harmonisation |
| ICU | Intensive Care Unit |
| IEC | International Ethics Committee |
| IHR | International Health Regulations |
| IL | Interleukin |
| KG | Kilogram |
| KM | Kaplan-Meier |
| LDH | Lactate Dehydrogenase |
| MedDRA | Medical Dictionary for Regulatory Activities |
| MERS | Middle East Respiratory Syndrome |
| MERS-CoV | Middle East Respiratory Syndrome Coronavirus |
| MG | milligrams |
| N | Number (typically refers to subjects) |
| NET | Neutrophil Extracellular Traps |
| PCR | Polymerase Chain Reaction |
| PHI | Protected Health Information |
| PI | Principal Investigator |
| PBMC | Peripheral Blood Mononuclear Cell |

| | |
|---|---|
| PO | Oral |
| PP | Per Protocol |
| RNA | Ribonucleic Acid |
| SAE | Serious Adverse Event |
| SAP | Statistical Analysis Plan |
| SARS | Severe Acute Respiratory Syndrome |
| SARS-CoV | Severe Acute Respiratory Syndrome Coronavirus |
| SOA | Schedule of Assessments |
| SOP | Standard Operating Procedure |
| SP | Safety Population |
| $spO_2$ | Oxygen Saturation |
| SUSAR | Suspected Unexpected Serious Adverse Reaction |
| UP | Unanticipated Problem |
| US | United States |
| WBC | White Blood Cell |
| WHO | World Health Organization |
