## Appendix 2: SAP for "Safety and Efficacy of Disulfiram in Hospitalized Patients with Moderate COVID-19: A Randomized, Double-Blind, Placebo-Controlled Trial"

DocuSign Envelope ID: 8F061211-20AA-42BB-A5E5-CF9CAC78AF08

ETICA and Spring Research Foundation

Augusto Mota, MD, PhD
Rua Altino Serbeto de Barros, 119
14º Andar Sala 1408
Pituba Salvador-Ba
CEP 41830-492

### A Randomized, Double-blind, Placebo-controlled Safety and Clinical Outcomes Study of Disulfiram in Subjects with Moderate COVID-19

SPR-001-201

Statistical Analysis Plan

Final Version: 1.0
Date: 29 July 2021

DocuSign Envelope ID: 8F061211-20AA-42BB-A5E5-CF9CAC78AF08

### SIGNATURE PAGE

This document is prepared by:

DocuSigned by:
Munish Mehra
5D39B22913DD4A2...

Date: 8/4/2021

Munish Mehra, MSc., MS, PhD
Lead Biostatistician
Tigermed

This document is reviewed by:

DocuSigned by:
Mark Marino
74E128FB0BD5487...

Date: 8/7/2021

Mark T Marino, MD
Medical Monitor
Mark T. Marino Consulting LLC

This document is approved by:

DocuSigned by:
Augusto Mota
69B552698E17483...

Date: 8/4/2021

Augusto Mota, MD, PhD
Study Sponsor and Principal Investigator
ETICA/Clinica AMO

DocuSigned by:
5395726981D04BE...

Date: 8/5/2021

Wendy Cousin, PhD
Study Director, Board Member
Spring Research Foundation

| MODIFICATION HISTORY | | | |
|---|---|---|---|
| **VERSION** | **VERSION DATE** | **AUTHOR** | **DESCRIPTION** |
| 1.0 | 29-JULY-2021 | MUNISH MEHRA | FIRST VERSION |

DocuSign Envelope ID: 8F061211-20AA-42BB-A5E5-CF9CAC78AF08

 Confidential

DocuSign Envelope ID: 8F061211-20AA-42BB-A5E5-CF9CAC78AF08

### List of Tables

Table 1. Objectives and Endpoints

Table 2. Study Procedures and Assessments

Table 3. Lists of Visits

Table 4. Risk Categories based on Age and Comorbidities

Table 5. Vital Signs Abnormalities

Table 6. Laboratory Tests Abnormalities

### ABBREVIATIONS

#### List of Abbreviations

| Abbreviation | Specification |
|---|---|
| AE | Adverse Event |
| ALT | Alanine Transaminase |
| ANCOVA | Analysis of Covariance |
| AST | Aspartate Transaminase |
| BMI | Body Mass Index |
| CBC | Complete Blood Count |
| Cf-DNA/NET | Neutrophil-derived circulating free DNA |
| CI | Confidence Interval |
| COVID-19 | Coronavirus Disease 2019 |
| CoVs | Coronavirus |
| CRF | Case Report Form |
| CSR | Clinical Study Report |
| CT | Computed Tomography |
| CTCAE | Common Terminology Criteria for Adverse Events |
| DNA | Deoxyribonucleic Acid |
| DSMB | Data and Safety Monitoring Board |
| ECG | Electrocardiogram |
| eCRFs | Electronic Case Report Forms |
| FDA | Food and Drug Administration |
| GCP | Good Clinical Practice |
| GFR | Glomerular Filtration Rate |
| HIV | Human Immunodeficiency Virus |
| IB | Investigator's Brochure |
| ICF | Informed Consent Form |
| ICH | International Council for Harmonisation |
| ICU | Intensive Care Unit |
| IHR | International Health Regulations |
| IL | Interleukin |
| KM | Kaplan-Meier |
| KG | Kilogram |
| LDH | Lactate Dehydrogenase |
| LLN | Lower Limit Normal |
| MedDRA | Medical Dictionary for Regulatory Activities |
| MERS | Middle East Respiratory Syndrome |
| MERS-CoV | Middle East Respiratory Syndrome Coronavirus |
| MG | Milligrams |
| mITT | Modified Intention-to-Treat |
| ML | Maximum Likelihood |
| MMRM | Mixed Model Repeated Measures |
| N | Number (typically refers to subjects) |
| NET | Neutrophil Extracellular Traps |
| PCR | Polymerase Chain Reaction |
| PHI | Protected Health Information |
| PI | Principal Investigator |
| PO | Oral |
| PP | Per Protocol Population |
| PT | Preferred Term |

DocuSign Envelope ID: 8F061211-20AA-42BB-A5E5-CF9CAC78AF08

| | |
|---|---|
| RNA | Ribonucleic Acid |
| SAE | Serious Adverse Event |
| SAP | Statistical Analysis Plan |
| SARS | Severe Acute Respiratory Syndrome |
| SARS-CoV | Severe Acute Respiratory Syndrome Coronavirus |
| SOA | Schedule of Assessments |
| SOP | Standard Operating Procedure |
| SP | Safety Population |
| $SpO_2$ | Oxygen Saturation |
| SUSAR | Suspected Unexpected Serious Adverse Reaction |
| TEAE | Treatment Emergent Adverse Event |
| ULN | Upper Limit Normal |
| UP | Unanticipated Problem |
| US | United States |
| WBC | White Blood Cell |
| WHO | World Health Organization |

DocuSign Envelope ID: 8F061211-20AA-42BB-A5E5-CF9CAC78AF08

### 1. INTRODUCTION

Coronaviruses (CoVs) are positive-sense single-stranded enveloped RNA viruses, many of which are commonly found in humans and cause only mild symptoms. Over the past two decades, emerging pathogenic CoVs capable of causing life-threatening disease in humans and animals have been identified, namely Severe Acute Respiratory Syndrome coronavirus (SARS-CoV) and Middle East Respiratory Syndrome coronavirus (MERS-CoV).

Coronavirus Disease 2019 (COVID-19) emerged in Wuhan, China in late 2019 and is caused by Severe Acute Respiratory Syndrome coronavirus 2 (SARS-CoV-2), which is a highly infectious zoonotic virus. Clinically, individuals with COVID-19 present with a myriad of symptoms, clinical course, and outcome. In order to develop effective treatments against COVID-19, it is necessary to understand the nature of the disease and the immunological response that occurs in these varying presentations. While the immune response - to pathogens has been well categorized (Type-1 Immunity, Type-2 Immunity, and Type-3 Immunity) and despite the numerous analyses that have been carried out to date, the correlation between an individual's immune response to a SARS-CoV-2 infection and their clinical outcome remain unclear.

COVID-19 is a respiratory disease caused by a novel coronavirus (SARS-CoV-2) and causes substantial morbidity and mortality. As of the start of this trial there was no vaccine to prevent COVID-19 or infection with SARS-CoV-2 or therapeutic agent to treat COVID-19. This clinical trial is designed to evaluate disulfiram for the treatment of adult subjects hospitalized with COVID-19.

This phase 2 study is being completed to evaluate the safety and clinical outcomes of disulfiram as compared to placebo in hospitalized subjects with moderate COVID-19 as well as the effect of disulfiram on selected biomarkers.

This statistical analysis plan (SAP) is based on protocol SPR-001-201 - A Randomized, Double-blind, Placebo-controlled Safety and Clinical Outcomes Study of Disulfiram in Subjects with Moderate COVID-19 (version 1.4, 09 July 2021). In this document, the contents and methods of statistical analysis will be described in detail).

### 2. STUDY OBJECTIVES

The overall objective of the study is to evaluate the clinical outcomes, safety, and the effect of disulfiram on selected biomarkers as compared to placebo in hospitalized subjects with moderate COVID-19. These objectives are detailed in Table 1.

#### Table 1. Objectives and Endpoints

| **OBJECTIVES** | **ENDPOINTS (OUTCOME MEASURES)** |
|---|---|
| **Primary** | |
| The primary objective is to assess the effect of disulfiram on clinical improvement. | To assess the time to clinical improvement, defined as the time from baseline to the first post-baseline assessment with an improvement in WHO score of ≥1 point. |
| **Secondary** | |
| To assess the effect of disulfiram on clinical outcomes | Key secondary:<br>1. To assess the mean number of days of supplemental oxygen (WHO Score ≥4).<br>2. To assess the time from baseline to discharge from the hospital.<br>3. To assess the percentage of subjects that are discharged by Day 8.<br>4. To assess the percentage of subjects that worsened 1 or more points on the WHO Ordinal Scale from baseline to any post baseline assessment through Day 28.<br>5. To assess the mean number of days of non-invasive ventilation or high flow oxygen devices or invasive mechanical ventilation (WHO Score 5 or 6) over the 28-day period.<br>6. To assess the mean number of days subjects were in the Intensive Care Unit (ICU).<br>7. To assess the percentage of subjects that were on non-invasive or high flow oxygen devices or invasive mechanical ventilation (WHO Score 5 or 6) over the 28-day period.<br>8. To assess the 28-day mortality. |
| To assess the effect of disulfiram on other measures of efficacy and various biomarkers. | Other Secondary:<br>1. To assess the mean change and percent change from baseline to Day 8 and Day 15 for Cytokine IL-18.<br>2. To assess the percentage of subjects requiring supplemental oxygen (WHO Score ≥4) by Day 8, 15, and 28.<br>3. To assess the percentage of subjects that are discharged by Day 15 and Day 28 |

| OBJECTIVES | ENDPOINTS (OUTCOME MEASURES) |
|---|---|
| | 4. To assess the percentage of subjects that worsened 1 or more points on the WHO Ordinal Scale from baseline through Day 8 and Day 15.<br>5. To assess the percentage of subjects admitted to the Intensive Care Unit.<br>6. To assess the percentage of subjects that improved 1 or more points on the WHO Ordinal Scale from baseline to Day 8, 15, and 28.<br>7. To assess the change and percent change in neutrophil count from baseline to Day 8 and 15.<br>8. To assess the change and percent change in total lymphocyte count from baseline to Day 8 and 15.<br>9. To assess the change and percent change from baseline to Day 8 and 15 for neutrophil-derived circulating free DNA (cf-DNA/NETs).<br>10. To assess the mean change and percent change from baseline to Day 8 and 15 for:<br>a. Cytokine TNF-α<br>b. Cytokine IL-1β<br>c. Cytokine IL-1RA<br>d. Cytokine IL-6<br>e. Cytokine IL-8<br>f. Cytokine IL-10<br>g. Lactate dehydrogenase (LDH)<br>h. D-dimer<br>11. To assess the association between baseline and worst post-baseline WHO score. |
| Safety | Incidence of Adverse Events (AEs) and Serious Adverse Events (SAEs). Vital Signs and Laboratory data will be collected during the study period. |

### 3. STUDY DESIGN

#### 3.1 General Design

This is a prospective, randomized, double-blind, placebo-controlled study to evaluate the safety and clinical outcomes of disulfiram administered to subjects diagnosed with moderate COVID-19. The multi-center study will be conducted up to 8 clinical sites in Brazil. The study is a 2-arm comparison between disulfiram and placebo. Eligible subjects will be enrolled and randomized (1:1) to either receive disulfiram (study drug) or placebo, orally (po) once daily for fourteen (14) days, stratified by the baseline risk factor specified in Table 4 below. Given the severity of illness in COVID-19, there are no pre-specified study stopping rules for safety.

DocuSign Envelope ID: 8F061211-20AA-42BB-A5E5-CF9CAC78AF08

The Data Safety Monitoring Board (DSMB), which will operate under a charter and review safety data. This SAP does not consider any analyses conducted by the DSMB.

The study schema is presented below:

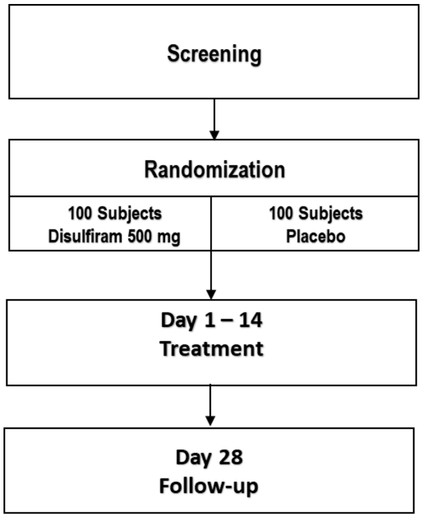

Visit-specific procedures and assessments are outlined in Table 2.

#### Table 2. Study Procedures and Assessments

| | | Treatment | | | | | Follow-up |
|---|---|---|---|---|---|---|---|
| **Activity** | **Screening Day -2 to 1**[1] | **Baseline Day 1** | **Days 2-7**[2] | **Day 8 ±2 days** | **Days 9-14**[2] | **Day 15 ±3 days** | **Day 28**[12] **±7 days** |
| **Informed Consent** | x | | | | | | |
| **Confirmation Eligibility** | | x | | | | | |
| **Demographics & Medical History** | x | | | | | | |
| **Concomitant Meds** | x | x | x | x | x | x | x |
| **Physical Exam** | x | x | | | | | |
| **Weight and Height** | x | | | | | | |
| **Vital Signs** | x | x | x | x | x | x | |
| **SpO2 (pulse oximetry)** | x | x | x | x | x | x | |
| **12 Lead ECG** | | x[3] | | | | | |
| **Supplemental Oxygen Use Assessment** | x | x | x | x | x | x | x |
| **Need for Invasive or Non-invasive Ventilation Assessment** | | | x | x | x | x | x |

| | | Treatment | | | | | Follow-up |
|---|---|---|---|---|---|---|---|
| **Activity** | **Screening Day -2 to 1[1]** | **Baseline Day 1** | **Days 2-7[2]** | **Day 8 ±2 days** | **Days 9-14[2]** | **Day 15 ±3 days** | **Day 28[12] ±7 days** |
| **ICU Admission Assessment** | | | x | x | x | x | x |
| **CBC and Serum Chemistry** | x | x | | x | | x | |
| **ALT, AST** | x[4] | | | | | | |
| **Estimated GFR** | x | | | | | | |
| **SARS-CoV-2 PCR or rapid antigen** | x[5] | | | | | | |
| **hCG Test** | x [6] | | | | | | |
| **CT Scan** | | x [7] | x [8] | x [8] | x [8] | x [9] | |
| **Cytokine levels[10]** | | x | | x | | x | |
| **LDH** | | x | | x | | x | |
| **cf-DNA/NETs[11]** | | x | | x | | x | |
| **D-dimer** | | x | | x | | x | |
| **WHO Ordinal Assessment** | | x | x | x | x | x | x |
| **Dispense Study Drug** | | x | | x | | | |
| **Study Drug Accountability** | | | | x | | x | |
| **AE** | | | x | x | x | x | x |

[1] The screening and baseline visit can be combined if all eligibility criteria are met.
[2] Visits only if hospitalized.
[3] If the subject had ECG already performed for this hospitalization, it can be used.
[4] If ALT and AST levels are available from this hospitalization, they do not need to be repeated.
[5] SARS-CoV-2 PCR or rapid antigen results from a test performed within 7 days of screening can be used.
[6] hCG pregnancy test, either urine or blood to be performed in female subjects of childbearing potential
[7] If subject had CT scan performed within 3 days of enrollment this can be used.
[8] If CT scan occurs as standard of care, capture in the source documents.
[9] The Day 15 CT scan only occurs if the subject is in the hospital on this visit day.
[10] Cytokine levels (Cytokine IL-18, Cytokine TNF-α, Cytokine IL-1β, Cytokine IL-6, Cytokine IL-10, Cytokine IL-1RA, and Cytokine IL-8)
[11] Circulating neutrophil DNA / neutrophil extracellular traps (cf-DNA/NETs)
[12]May be conducted remotely

DocuSign Envelope ID: 8F061211-20AA-42BB-A5E5-CF9CAC78AF08

##### 3.2 Sample Size and Power Considerations

Since the efficacy of disulfiram on COVID-19 infected patients is not known, the sample size for this study is not based on any formal calculations.

The sample size of 200 subjects and formal statistical analyses detailed in this SAP with control for multiplicity will allow testing for the primary and key secondary endpoints with an overall Type I error of < 0.05.

##### 3.3 Study Population

The study population consists of hospitalized adult male and female subjects aged 35 years or older with moderate COVID-19. Inclusion and exclusion criteria can be found in Sections 5.1 and 5.2 of the Protocol.

#### 4 ANALYSIS POPULATION

Approximately 200 subjects hospitalized with moderate COVID-19 who meet all eligibility criteria will be enrolled at up to 8 clinical sites in Brazil.

The estimated time from screening (Day -2 to 1) to end of study (Day 28 ± 7 days) for an individual subject is approximately 37 days.

##### 4.1 Randomized Population

The randomized population includes all subjects who were randomized. This is also referred to as the Full Analysis Set (FAS) or Intent-to-treat (ITT) population. Currently minimal to no difference is expected between this population and the mITT or Safety population hence no analyses are planned on this population. If at the end of the study, there are differences warranting further investigation additional analyses may be performed on this ITT population. For this population subjects are allocated based on treatment group they were randomized to regardless of which treatment they actually received.

##### 4.2 Modified Intent-to-Treat (mITT) Population

The modified intent-to-treat (mITT) population will include all randomized subjects who receive a dose of study drug and have WHO score measured both at the baseline visit and at least one post-baseline time point. The subjects will be grouped as randomized irrespective of the treatment they received. The mITT population will be used for all efficacy analyses.

DocuSign Envelope ID: 8F061211-20AA-42BB-A5E5-CF9CAC78AF08

##### 4.3 Per Protocol (PP) Population

The per protocol (PP) population is a subset of the mITT population and includes all subjects who did not have any important protocol deviations significantly impacting efficacy assessment, the subject's rights, or their safety. For this population, subjects will be grouped based on treatment they received. The per protocol population will be used for sensitivity analysis of the primary efficacy endpoints and may also be used for selected secondary endpoints.

##### 4.4 Safety Population (SP)

The safety population (SP) will consist of all subjects randomized and receiving at least one dose of study drug. The subjects will be grouped according to the treatment they actually received. The safety population will be used for safety analyses.

#### 5 GENERAL CONSIDERATIONS FOR DATA HANDLING AND ANALYSES

##### 5.1 Data Management

Clinical research data from source documentation (including, but not limited to, WHO Scale, AEs, SAEs, concomitant medications, medical history, clinical laboratory data, etc.) will be entered by the clinical study sites into a US FDA 21 CFR Part 11 compliant validated EDC system.

AEs will be coded according to the MedDRA Dictionary, Version 23.1, while concomitant medications will be coded according to the WHO Drug Global Dictionary, Version September 1, 2020, B3. Medical history will not be coded or tabulated.

Derived dataset, statistical programming and analyses will be performed using SAS® Version 9.4 or higher.

##### 5.2 Data Standard

When data such as for Biomarkers are skewed and transformations are necessary, data will be transformed using the log base 2 function. A unit change in the log base 2 transformed value is equivalent to a doubling of the value of the biomarker. In this case, if a particular observation takes the value of 0 or OOR<, then it should be replaced by the value of 0.0001, before taking the log.

If data values are in the form of "≤x", "<x", ">x", or "≥x", then the value of x will be used for analyses, but the reported value will be listed. For example, a value reported as ">200" will be listed as such but for analyses (e.g., when reporting the mean), then 200 will be used. Other approximations for Biomarkers below level of quantification such as taking the mid-point between lower limit of quantification and 0 may also be used.

#### 5.3 Baseline Definition

The baseline value for efficacy and safety analyses is the last non-missing observed data prior to the first dose of study drug.

#### 5.4 Visits

There are 5 main visits as indicated in Table 3 below. During hospitalization there are additional required visits, every day of hospitalization through to Day 15. There may be unscheduled visits at which data are collected. For analysis of efficacy data only scheduled visits will be used. For summaries of abnormalities in Vital Signs or Laboratory data from all unscheduled and scheduled visits will be used.

For analysis of safety and efficacy at a given timepoint such as change from baseline to a post baseline visit or proportion of subjects discharged by a certain visit, results will only be presented at main visit.

**Table 3. Lists of Visits**

| Scheduled Visits | Relative Target Day | Main Visits |
|---|---|---|
| Screening[1] | -2 to 1 | Yes |
| Day 1 (Baseline) | 1 | Yes |
| Day 2-7 | | No |
| Day 8 Visit | 6 to 10 | Yes |
| Day 9-14 | | No |
| Day 15 Visit | 12 to 18 | Yes |
| Day 28 Visit | 21 to 35 | Yes |

[1] The Screening and Baseline visit can be combined if all eligibility criteria are met.

In listings data collected at all scheduled and unscheduled visits will be presented.

#### 5.5 Derivation and Transformation of Data

##### 5.5.1 Baseline Age

Variable Age is captured on the eCRF, so no derivation is necessary.

##### 5.5.2 Change and Percent (%) Change from Baseline

Absolute change from baseline is calculated as (post-baseline result – baseline result).

DocuSign Envelope ID: 8F061211-20AA-42BB-A5E5-CF9CAC78AF08

Percentage change from baseline is calculated as [100*(post-baseline value – baseline value) / (absolute value at baseline)].

If either the baseline or the post-baseline result is missing, the change from baseline is set to missing as well. If the baseline result is missing or 0, or if the post-baseline result is missing, then % Change from Baseline will be set to missing at that visit.

##### 5.5.3 Number of Days from Baseline

The number of days from baseline to a pre-baseline event is calculated as:

Date of baseline - Date of the pre-baseline event

The number of days from baseline to a post-baseline event is calculated as:

(Date of the post-baseline event – Date of baseline) + 1

The day of treatment is considered Day 1. There is no Day 0.

##### 5.5.4 Duration of an Event in Number of Days

The length of an event, as measured by number of days, is defined as:

(Last Date of the event – First Date of the event) + 1

The number of days is calculated irrespective of the time the event started/ended.

#### 5.6 Subjects in Study

##### 5.6.1 Subject Disposition

The number of subjects who signed an informed consent but were not randomized (screen failures) will be summarized along with the reason for being a screen failure.

The number of subjects treated (subject received at least one dose of study drug), completed through 28 and discontinued from the study will be tabulated by treatment group using the randomized population. For subjects who prematurely discontinue the study, the number and percentage of subjects that discontinue and the reasons for doing so will be tabulated. The reasons will be ordered in decreasing frequency of all subjects.

##### 5.6.2 Protocol Deviation

As per ICH E3 guideline Section 10.2, All important deviations related to study inclusion or exclusion criteria, conduct of the trial, patient management or patient assessment should be summarized by site and grouped into different categories, such as:

- those who entered the study even though they did not satisfy the entry criteria
- those who developed withdrawal criteria during the study but were not withdrawn

DocuSign Envelope ID: 8F061211-20AA-42BB-A5E5-CF9CAC78AF08

- those who received the wrong treatment or incorrect dose
- those who received an excluded concomitant treatment

Additional details of reviewing and classifying protocol deviations as “Important” or “Not Important” are detailed under Section 8 -Protocol Deviations of Data Management Plan final version 1.0 dated 30th Oct 2020.

Final categorization of protocol deviations will be done prior to database lock and unblinding and documented.

Any protocol deviations and important protocol deviations will be summarized by treatment group and site.

##### 5.6.3 Handling of Missing or Partial Start and Stop Dates for Prior and Concomitant Medication

The following imputation rules will be used in this study to classify medications as prior or concomitant medications:

- Partial or missing start date for concomitant medication
    1. Incomplete or partial start date will not be imputed if end date of concomitant medication is on or before treatment start date. These are prior medications.
    2. Incomplete or missing start date will be imputed as mentioned below if end date of concomitant medication falls after treatment start date (or if medication is ongoing at end of study for treated subjects). These are concomitant medications that may also be prior medications based on imputation rules below.
- Partial Start Date
    1. Missing day: impute the 1st of the month; if month and year are the same as month and year of the first dose of study drug then impute the first dose date.
    2. Missing day and month: impute 1st January; if year is the same as first dose date, then impute first dose date.
- Partial End Date
    1. Missing day: impute the last day of the month; if month and year are the same as month and year of the last dose of study drug then impute the last dose date.
    2. Missing day and month: impute 31st December; if month and year are the same as the last dose date then impute the last dose date.

If a medication end date is completely missing or it is unclear whether the medication was taken prior to or in the treatment period, the medication will be assumed to be taken both as a prior and concomitant medication.

##### 5.6.4 Missing Start and Stop Dates for Adverse Events

Due to the short duration of the study the amount of partial or missing start and stop dates in this study is expected to be minimal. The following imputation rules will be used in this study for determining when Adverse Events started and ended and if they were treatment emergent.

- Partial or missing start date
    1. Missing day: impute the 1st of the month; if month and year are the same as month and year of the first dose of study drug then impute the first dose date.
    2. Missing day and month: impute the 1st of January; if year is the same as the first dose date, then impute the first dose date.
- Partial or missing end date
    1. Missing day: impute the last date of the month; if month and year are the same as month and year of the last dose of study drug, then impute the last dose date.
    2. Missing day and month: impute 31st December; if year is the same as the last dose date, then impute the last dose date.

If an AE start date is completely missing or it is unclear whether the event occurred prior to or in the treatment period, the AE will be assumed to be treatment emergent (after initiation of first treatment).
The above is a conservative approach to categorize any potential AE as a TEAE.

#### 5.7 Early Termination Visit

For efficacy analyses the early termination visit observations will be allocated to the nearest visit for which that endpoint was planned to be collected. If data for the endpoint was collected at that visit, then the early termination visit will be assigned to the next visit for which that endpoint was planned to be collected.

#### 5.8 Demographic and Baseline Characteristics

Subject demographics and other baseline characteristics will be summarized by treatment group and overall.

Continuous variables such as age, weight, height, and BMI will be described by their mean, standard deviation, median, minimum, maximum, 25$^{th}$ and 75$^{th}$ percentiles and number of missing values per treatment group and overall.

Categorical variables (e.g., race) will be summarized using counts and percentages. Percentages are calculated using the total subjects per treatment group with available data (=100%). Percentages will be rounded to one decimal place and therefore, there may be occasions when the total of the percentages does not exactly equal 100%. Denominators used for computation of percentages will be indicated in footnotes of tables.

Missing categories will be presented when relevant.

The following demographic variables along with the subgroups mentioned in Section 6.2.3 will be descriptively summarized at baseline.

- Sex (male or female)
- Age in years

- Age group
- Weight (kg)
- Height (cm)
- Body Mass Index (BMI)
- Hypertension
- Diabetes
- Race
- Baseline WHO scores
- Baseline risk categories

#### 5.9 Comorbidities

The number and the percentage of subjects for each risk group (number of comorbidities and age group) will be summarized by treatment group and overall, per Table 4.

**Table 4. Risk Categories based on Age and Comorbidities**

| | **Comorbidities:** Hypertension (on any prescription anti-hypertension medication), Diabetes (on oral or insulin injections for diabetes), BMI ≥ 35 | | | |
|---|---|---|---|---|
| **Age** | 0 | 1 | 2 | 3 |
| 35-59 | Low | Low | Low | Low |
| 60-69 | Low | Medium | Medium | Medium |
| 70-79 | Medium | High | High | High |
| ≥ 80 | High | Very High | Very High | Very High |

##### 5.9.1 Medical History

Medical history for each subject will only be presented in listings.

##### 5.9.2 Prior and Concomitant Medications

All medications (e.g., both prior and concomitant) will be coded using the WHO Drug Global Dictionary, Version September 1, 2020, B3. Anatomical Therapeutic Chemical drug class 2 will be used. The incidence of medications will be summarized by standard medication class and standardized medication. Subjects are counted only once in each standard medication class category, and only once in each standardized medication category. Prior medications will include all medications taken prior to a subject taking the first dose of study drug. Any medication given at least once on or after the date of first day of study drug treatment will be defined as a concomitant medication.

The standard medication class will be ordered by decreasing frequency of all subjects and then alphabetically. The standardized medication will be ordered by decreasing frequency of all subjects within the standard medication class then alphabetically.

#### 5.10 Data Presentation Conventions and General Approach

All data collected in the study will be presented in individual data listings. All listings will include the Subject ID and treatment group. Where applicable, listings will also include visit number, visit date, and days relative to the initiation of treatment. Individual subject data listings for data collected by visit are sorted by Subject ID and Visit.

When missing or partial dates are imputed, the listings will display the original and imputed dates, and if space allows a flag that indicated if the date has been derived.

Tables and listings should be self-contained, complete, accurate and concise; all relevant information should either be in the titles, columns, row headers, or in the footnotes; units must be always specified for all appropriate data unless obvious (either in the title, footnotes or body of a table or listing.).

If possible, variables being summarized, and statistics reported should appear in the first column on the left of a table. The next column for treatment group should report data from left to right for the Disulfiram, Placebo and (depending on the table) overall treated subjects, respectively.

In each table, denominators used to compute percentages will be indicated in the footnotes of the table or should be clear in the title or in the body of the table; the summary table will clearly indicate the population based for the analysis, number of subjects to which the data apply along with an indication for the number of subjects with missing data. Missing data are included in the calculation of percentages, unless otherwise specified.

Row entries in summary tables are always displayed for every category even if no data exist for any subjects (for example a row with all zeros will be displayed), unless otherwise specified.

The “Missing” entry for a categorical variable should be the last item reported in the Table. The “Other” entry of a categorical variable, if applicable, should not be listed alphabetically, but presented at the end of the list, just before the “Missing” category. More information can be found in the relevant Table Shells.

Actual tables and listings final outputs might differ slightly from tables and listings mock shells.

The following conventions are applicable in all data presentations and summaries:

1. For continuous variables such as mean, 25$^{th}$ and 75$^{th}$ percentile, and median values are formatted to one more decimal place than the measured value. Standard deviation values are formatted to two more decimal places than the measured type unless there are space limitations or enough significant digits in which case one more decimal place is sufficient. Minimum and maximum value are presented to the same accuracy to which the variable was recorded and presented. This rule may be modified to fit data in a given Table as long as the data displays relevant decimal places for interpretation of results.
2. For categorical variables, the number and percentages are presented in this form: XX (XX.X), with the percentage within the parenthesis rounded to 1 decimal place. If the percentage is 8.7 then 8.7 should be reported (rather than 08.7), with aligned parenthesis across the table.

3. Where possible data should be center, right, or left aligned as appropriate to display the data and column headers in the best possible way. Parenthesis will also be aligned appropriately for the best display for that particular table. See the mock shells for further details.
4. P-values, when applicable, will be presented to 3 decimal places. If the p-value is less than 0.001 then it will be displayed as <0.001. If the rounded result is 1.000 then it will be displayed as >0.999.

Summary tables and listings will include a “footer” providing explanatory notes that indicate as a minimum:

- Date and time of output generation
- SAS program name, including the path that generates the output
- Any other output specific details that require further elaboration
- Page number in the format of Page X of Y

The default convention is to number tables and listings using a decimal system to reflect main levels of unique tables and listings and sub-levels of replicate tables and listings with a maximum of 4 numbers separated by 3 decimal digits (for example Table WW.XX.YY.ZZ).

Subject accounting, final disposition and baseline and demographic profiles should appear in series of tables starting with 14.1; efficacy analyses should come next in the 14.2 series; safety should correspond to series 14.3.

### 6 EFFICACY EVALUATIONS

#### 6.1 General Considerations

The primary efficacy endpoint and key secondary efficacy endpoints will be tested sequentially in the order specified in Table 1, to account for multiplicity and preserve the overall Type I error at 0.05. No adjustments will be made for multiple comparisons in testing other secondary or exploratory efficacy endpoints.

Each statistical test will be performed at Type I error $\alpha$=0.05 (Two-sided).

Summary tables for each efficacy endpoint will be presented for each post baseline visits (Day 8, Day 15, and Day 28 visits when relevant).

Individual components of composite endpoints will be summarized when applicable.

Descriptive statistics for continuous and categorical variables will be presented as described in Section 5.10

#### 6.2 Efficacy Analysis

##### 6.2.1 Analysis on Primary Efficacy Endpoint

The primary efficacy endpoint of this study is to assess the time to clinical improvement of one or more points on the WHO 7-point ordinal scale using the mITT population.

The WHO 7-point ordinal scale (or WHO score) is an assessment of the clinical status. The scale is as follows:

1. Not hospitalized, no limitations on activities
2. Not hospitalized, limitation on activities
3. Hospitalized, not requiring supplemental oxygen
4. Hospitalized, requiring supplemental oxygen
5. Hospitalized, on non-invasive ventilation or high flow oxygen devices
6. Hospitalized, on invasive mechanical ventilation or ECMO
7. Death

Improvement will be defined as a decrease of at least one point on the WHO score compared to the baseline value (e.g., from 4 to 3; from 4 to 2 etc.). The time to improvement will be defined as the time (in days) from baseline to the earliest day of improvement at any post baseline WHO assessment.

The hypotheses test for testing the difference between Disulfiram and Placebo in the primary efficacy endpoint of time to improvement through Day 28 will be performed using a log rank test.

The null hypothesis assumes no difference in Kaplan-Meier (KM) curves of time to improvement (identical) in the active treatment group (Disulfiram) compared to the placebo group. The alternative hypothesis is that there is a difference in KM curves between time to improvement in the active treatment group compared to the placebo group.

Both hypotheses are represented below:

**$H_0$**: The two time to improvement KM curves are identical (or $S_{t1} = S_{t2}$)

**$H_A$**: The two time to improvement KM curves are not identical (or $S_{t1} \neq S_{t2}$, at any time t)

Where $S_{t1}$ is the KM curve for time to improvement of ≥1 point from Baseline to Day 28 visit for subjects randomized to Disulfiram (Active) treatment group and St2 is the KM curve for time to improvement of ≥1 point from Baseline to Day 28 visit for subjects randomized to the placebo group.

Subjects who are lost to follow-up or terminate the study prior to Day 28 visit and prior to observing the event will be censored at the time of their last observed assessment. Subjects who die will be assumed to have been censored at Day 28 with respect to time to improvement, giving deaths the worst rank. The Kaplan-Meier estimates for the time to clinical improvement will be presented along with the proportions of subjects who had clinical improvements by days 8, 15, and 28.

##### 6.2.2 Sensitivity Analysis on Primary Efficacy Endpoint

Below sensitivity analyses will be performed for the primary efficacy endpoint.

- Using the PP population.
- With baseline risk factors as a stratum using stratified log rank test with mITT population. If there are not sufficient events in an individual risk category, categories will be combined (i.e., with adjacent level) and then analyzed.
- Using Cox proportional hazards models to estimate the hazard ratio and CIs on mITT population

##### 6.2.3 Subgroup Analyses

Subgroup analyses for the primary and all key secondary endpoints will evaluate the homogeneity of treatment effect across the following subgroups:

- Sex (Male; Female)
- Age group (< vs ≥ Median)
- Baseline risk categories (Low; Medium; High; Very High)
- Comorbidities at baseline
    - Hypertension (Yes/No)
    - Diabetes (Yes/No)
    - BMI (< vs ≥ 35)
    - BMI (< vs ≥ 40)
- Subgroup by hyperinflammation state at baseline
    - NLR ratio < vs ≥ 6.1. (Defined as dividing the absolute neutrophil count by the absolute lymphocyte count)
    - Biomarker levels (IL-18) (< vs ≥ Median)
    - Biomarker levels (IL-18) (< vs ≥ 400 pg/mL)
    - D-dimer level (< vs ≥ 1.0 μg/mL)
- Subgroup by clinical status (Screening WHO score (3 vs 4))
- Other parameters at baseline associated with increased mortality:
    - Absolute Lymphocytes (< vs ≥ 0.8 x10^9/L)
    - Absolute Neutrophils (< vs ≥ 8 x10^9/L)
    - Platelets Count (< vs ≥ 150 x10^9/L)
    - WBC (< vs ≥ 10 x10^9/L)

Additional subgroup analyses might be conducted, following the same approach. A forest plot will display confidence intervals across subgroups. Hypothesis tests will be conducted on the interaction terms to determine whether the effect of treatment varies by subgroup.

To assess statistical significance of interaction term the following models will be used.

- Time to event endpoints - cox regression
- Categorical endpoints - logistics regression
- Continuous endpoints - ANCOVA

#### 6.3 Analysis on Secondary Efficacy Endpoints

##### Key Secondary Endpoints

As stated in Section 6.1, testing of the statistical hypothesis will be conducted hierarchically (in the order shown in Table 1) for the key secondary endpoints.

Significance testing will be done sequentially for each key secondary endpoint using α = 0.05 (two-sided), to preserve the overall Type I error. As soon as a test yields a p-value greater than 0.05 no further tests following it will be considered statistically significant, however p-values will be presented and considered nominal.

Summary tables with descriptive statistics and p-value will be prepared for all key and non-key secondary efficacy endpoints.

##### General Considerations

Continuous endpoints will be analyzed using the raw values, however if data are skewed and model assumptions might be violated, then the log or other transformations may be performed.

Categorical endpoints will compare proportions between the active and placebo groups using a binomial logistic regression model adjusted for baseline risk category. If logistic regression cannot be appropriately fit based upon the distribution of covariates between treatment groups a Chi-squared and/or Fisher's exact test, unless otherwise stated, will be used to test the hypothesis.

##### 6.3.1 To assess the mean number of days of supplemental oxygen (WHO Score ≥4)

The hypothesis testing to compare the mean number of days of supplemental oxygen between two treatment groups will be tested using an analysis of covariance (ANCOVA) model with treatment as a fixed effect; adjusted for covariates: baseline risk category and the interaction between treatment groups and baseline risk categories. Death at any time point will be assigned a score of 28 days.

Significance testing will be based on Least Square (LS) Means. LS means (95% CI of LS means), Standard Error (SE), LS mean vs placebo, 95% CI and p-value from above mentioned ANCOVA model will be presented.

##### 6.3.2 To assess the time from baseline to discharge from the hospital

Time from baseline to discharge will be analyzed similar to the primary efficacy endpoint as described above. The time to discharge will be defined as the time (in days) from baseline to when the subject is first discharged from hospital.

##### 6.3.3 To assess the percentages of subjects that are discharged by Day 8

The hypothesis testing to compare the percentage of responders between the two treatment groups will be tested using binomial logistic regression. The dependent variable for logistic

DocuSign Envelope ID: 8F061211-20AA-42BB-A5E5-CF9CAC78AF08

regression will be whether a subject is a responder or not. The independent variables in the model will be treatment group and baseline risk category.

The placebo group will be the reference treatment and the PARAM option in the CLASS statement will be used. Subject discharged at Day 8 visit will be defined as the event. The odds ratio together with the Wald's two-sided 95% confidence interval and p-value will be reported using logistic regression model.

In addition to above analysis, the frequency and percentage of subjects will be presented by treatment group together with the two-sided 95% Clopper-Pearson confidence interval.

##### 6.3.4 To assess the percentage of subjects that worsened 1 or more points on the WHO Ordinal Scale from baseline to any post baseline assessment through Day 28.

Subjects that worsened in WHO 7-point scale from baseline to any post baseline visit. Worsening will be defined as an increase of at least one point on the WHO 7-point scale compared to the baseline value (e.g., from 3 to 4; from 4 to 6 etc.). This efficacy endpoint will be evaluated with the same approach mentioned in Section 6.3.3.

##### 6.3.5 To assess the mean number of days of non-invasive ventilation or high flow oxygen devices or invasive mechanical ventilation (WHO Score 5 or 6) over 28 days period

For this endpoint, the following WHO scores will be considered:

- Subjects hospitalized, on non-invasive ventilation (5) or on invasive mechanical ventilation (6) or Death (7).
- Subject hospitalized, on non-invasive ventilation or high flow oxygen devices (5)
- Subjects hospitalized, on invasive mechanical ventilation or ECMO (6)

Although significance testing will be conducted for all 3 approaches, only the first one will be considered for hierarchical testing. This efficacy endpoint will be evaluated with same approach stated in Section 6.3.1.

##### 6.3.6 To assess the mean number of days of subjects were in the Intensive Care Unit (ICU)

This efficacy endpoint will be evaluated with same approach stated in Section 6.3.1.

##### 6.3.7 To assess the percentage of subjects that are on non-invasive ventilation or high flow oxygen devices or invasive mechanical ventilation (WHO Score 5 or 6) over 28 days period

Not requiring non-invasive ventilation and invasive mechanical ventilation at Day 28 visit will be defined as the event.

For this objective, results will also be presented based on different values of WHO score:

- Subjects hospitalized, on non-invasive ventilation (5) or on invasive mechanical ventilation (6) or Death (7).

- Subject hospitalized, on non-invasive ventilation or high flow oxygen devices (5)
- Subjects hospitalized, on invasive mechanical ventilation or ECMO (6)

Although significance testing will be conducted for all 3 approaches, only the first one will be considered for hierarchical testing. This efficacy endpoint will be evaluated with same approach stated in Section 6.3.3.

##### 6.3.8 To assess the 28-day mortality

To test the difference in proportion of subjects dying from any cause over an assessment period from start of study treatment until Day 28 visit between the treatment groups Chi-Square test will be used. In case expected cell frequency is less than 5 then Fisher's exact test will be used for analysis and the corresponding confidence interval for the difference in proportion will be calculated.

#### 6.4 Other Efficacy Endpoints

Data will be summarized with descriptive statistics for each planned visit, when relevant, and changes from Baseline to Day 8, 15 and 28. Hypothesis tests will be conducted for non-key efficacy endpoints similar to key secondary endpoints. No adjustment will be made for multiplicity for other efficacy endpoints.

No sensitivity or subgroup analyses are planned for non-key secondary endpoints.

1. The mean change and percent change from baseline to Day 8 and Day 15 for Cytokine IL-18. Below analysis approach will be followed.

   For this endpoint analysis, no imputation will be used, and a Mixed Model Repeated Measures (MMRM) analysis will be used on Log transformed data to estimate the treatment effect of change from Baseline to Day 8 and Day 15 as stated in Section 5.2.

   The MMRM model will include change from baseline as the dependent variable and treatment group, visit, the baseline value and, baseline risk category as the main effects and the interaction between treatment groups and visit.

   Since some risk categories might have low number of subjects, it may be necessary to combine risk categories. This will only be done for adjacent categories (e.g., low with medium, high with very high).

   The Placebo treatment group and the low category will be used as the reference level for treatment and baseline risk, respectively.

   The Kenward-Roger approximation of degrees of freedom will be used.

   If the model with the unstructured correlation matrix fails to converge and/or the Hessian matrix is not positive definite then following covariance structures will be tested in order below. The first covariance structure that converges will be used.

   1. Toeplitz with heterogeneity (TOEPH)
   2. Autoregressive with heterogeneity (ARH (1))

DocuSign Envelope ID: 8F061211-20AA-42BB-A5E5-CF9CAC78AF08

3. Compound symmetry with heterogeneous variances (CSH)
4. Toeplitz (TOEP)
5. Autoregressive (AR (1))
6. Compound symmetry without heterogeneous variances (CS)

Analysis of IL-18 and other Biomarkers all inference to compare Disulfiram vs placebo on Day 8 will be based on log transformed data using above MMRM methodology. To review results on Biomarkers in units collected (non-log-transformed data) results will be converted using exponentiation and presented as the geometric mean and coefficient of variation.

2. The frequency and percentages of subjects requiring supplemental oxygen (WHO Score ≥4) overall and with individual WHO scale of 4, 5, and 6 through Day 8, Day 15 and 28. Same analysis approach mentioned in Section 6.3.3 will be followed.
3. The frequency and percentages of subjects that are discharged by Day 15 and 28. Same analysis approach mentioned in Section 6.3.3 will be followed.
4. The frequency and percentages of subjects that worsened 1 or more points on the WHO score from baseline through Day 8, 15 on the WHO score. Same analysis approach mentioned in Section 6.3.3 will be followed.
5. The frequency and percentages of subjects that are admitted to the Intensive Care Unit (ICU). Same analysis approach mentioned in Section 6.3.3 will be followed.
6. The frequency and percentages of subjects that improved from baseline to Day 8, 15 and 28 with improvement of 1 or more points on the WHO score. Same analysis approach mentioned in Section 6.3.3 will be followed.
7. The change and percentage change from baseline in neutrophil count to Day 8 and Day 15. Same analysis approach mentioned in above point 1 will be followed.
8. The change and percentage change from baseline in total lymphocyte count to Day 8 and Day 15. Same analysis approach mentioned in above point 1 will be followed.
9. The change and percentage change from baseline in neutrophil-derived circulating free DNA (cf-DNA/NETs) to Day 8 and Day 15. Same analysis approach mentioned in above point 1 will be followed.
10. The mean change and percentage change from baseline to Day 8 and Day 15 for the following biomarkers with observed and log transformed values will be assessed appropriately: Cytokine TNF-α, Cytokine IL-1β, Cytokine IL-1RA, Cytokine IL-6, Cytokine IL-8, Cytokine IL-10, Lactate dehydrogenase (LDH) and D-dimer. Same analysis approach mentioned in above point 1 will be followed.
11. To assess the association between baseline and worst post-baseline WHO score a shift table for changes in the WHO score from baseline to worst post-baseline score will be presented for each treatment arm. The hypothesis testing to measure the ordinal association between the baseline WHO score (3 and 4) and worst post-baseline WHO score (1-7) variables of the contingency table will be done using non-parametric test Goodman-Kruskal's Gamma within each treatment group. Significance testing will be based on gamma statistics, Gamma value, 95% CI and p-value from the above-mentioned test will be presented.

DocuSign Envelope ID: 8F061211-20AA-42BB-A5E5-CF9CAC78AF08

### 7 EXPLORATORY EFFICACY ANALYSES

Exploratory efficacy analyses are not covered in this SAP. Any exploratory analyses performed to support study results which are not identified in this SAP, will be documented, and reported in the CSR.

### 8 SAFETY ANALYSIS

#### 8.1 General

The safety population will be used for all safety analyses. Summaries will be presented by the treatment actually received.

All safety data will be summarized with descriptive statistics with no hypothesis testing.

#### 8.2 Study Drug Administration

The exposure to the study drug is defined as the number of administered doses which is the number of doses dispensed minus the number of doses returned. Exposure will be summarized by treatment group and overall.

Compliance (%), defined as 100*(number of doses taken divided by number of doses supposed to be taken over 14 days), will be summarized by treatment group and overall.

Treatment duration (days) is defined as the last date study drug was taken minus the date of the first study drug intake plus 1.

#### 8.3 Adverse Events

The safety analysis will be conducted on the safety population. All safety data will be summarized by descriptive statistics, with no hypothesis testing.

Adverse events (AEs) will be coded using MedDRA, Version 23.1. A treatment emergent adverse event (TEAE) is defined as an AE that occurs for the first time after initiation of treatment or if it had occurred prior to treatment, it worsens in severity after initiation of treatment. If AE dates are incomplete and it is not known if the AE was TEAE or not, then it is assumed to be a TEAE.

When reporting an AE by severity only the worst severity will be used.

AEs which are not TEAEs will be only included in listings.

A summary table will show the total number of TEAE and the frequency and percentage (within the treatment group) of subjects by treatment group with at least one AE of the following:

- All TEAE
- Serious TEAE

DocuSign Envelope ID: 8F061211-20AA-42BB-A5E5-CF9CAC78AF08

- Serious treatment related TEAE
- TEAE by severity
- Treatment related TEAE
- Severe Adverse Events
- AE leading to study discontinuation
- AE leading to treatment discontinuation
- AE leading to death
- Suspected Unexpected Serious Adverse Reactions (SUSAR)

Separate tables showing the System Organ Class (SOC) and Preferred Team (PT) will show the following AEs:

- All TEAE
- Serious TEAE
- Serious treatment related TEAE
- TEAE by worst severity
- Severe Adverse Events
- Treatment related TEAE
- AE leading to study discontinuation
- AE leading to treatment discontinuation
- AE leading to death
- Suspected Unexpected Serious Adverse Reactions (SUSAR)

The SOC will be sorted by decreasing frequency of all subjects then alphabetically. PTs will be sorted within SOC by decreasing frequency of all subjects then alphabetically. Subjects will be counted at most once per SOC and at most once per PT.

#### 8.4 Vital Signs

Vital signs values, their change from baseline and frequency of vital signs abnormalities will be summarized by treatment and visit. All vital signs measurements will be listed.

The following parameters are collected: temperature, heart rate, systolic blood pressure, diastolic blood pressure, oxygen saturation and respiratory rate. The thresholds in Table 5 are considered clinically significant abnormalities for each vital sign.

DocuSign Envelope ID: 8F061211-20AA-42BB-A5E5-CF9CAC78AF08

##### Table 5. Vital Signs Abnormalities

| Vital Sign | Abnormality Values |
|---|---|
| Temperature (°C)[1] | ≤ 32°C<br>> 40°C |
| Heart Rate (beats per minute)[1] | < 60 bpm<br>> 100 bpm |
| Systolic Blood Pressure (mm Hg) | ≤ 90 mm Hg2<br>≥ 160 mm Hg1 |
| Diastolic Blood Pressure (mm Hg) | ≤ 60 mm Hg2<br>≥ 100 mm Hg1 |
| Oxygen Saturation - $SpO_2$ (%)[1] | ≤ 88% |
| Respiratory Rate (breaths / min)[3] | ≤ 12 breaths / min<br>≥ 25 breaths / min |

[1]Abnormalities are based on Grade 3 or higher of Common Terminology Criteria for Adverse Events (CTCAE) V5.0.
[2] https://www.mayoclinic.org/diseases-conditions/low-blood-pressure/symptoms-causes/syc-20355465#:~:text=In%20severe%20cases%2C%20low%20blood,generally%20considered%20low%20blood%20pressure
[3] https://my.clevelandclinic.org/health/articles/10881-vital-signs

#### 8.5 Laboratory Test Results

Clinical laboratory tests will be converted to consistent units according to the International System of Units (SI) before summarization.

Clinical laboratory values and their change from baseline will be summarized by visit. Since visits date are not available in laboratory file, data will be sorted by date and visit ID will be assigned sequentially based on the actual date.

Shift tables for laboratory tests between baseline and each post-baseline visit based on a classification of values as low, normal, or high with respect to the reference range will be summarized for subjects with values for both baseline and the relevant visit. The percentage will be based on the number of treated subjects in each treatment group.

All laboratory values will be listed, alongside abnormalities. Table 6 below, shows a list of laboratory tests and their abnormality thresholds (for continuous variables), based on Grade 3 or higher of CTCAE V5.0.

##### Table 6. Laboratory Tests Abnormalities

| Category | Laboratory Test (Reported Unit) | Abnormality Values – Grade 3 or higher of CTCAE V5.0 |
|---|---|---|
| Chemistry | Alanine aminotransferase - ALT (U/L) | > 5.0 x ULN |

DocuSign Envelope ID: 8F061211-20AA-42BB-A5E5-CF9CAC78AF08

| **Category** | **Laboratory Test (Reported Unit)** | **Abnormality Values – Grade 3 or higher of CTCAE V5.0** |
|---|---|---|
| Chemistry | Aspartate aminotransferase - AST (U/L) | > 5.0 x ULN |
| Chemistry | Creatinine (umol/L) | > 3.0 x ULN |
| Chemistry | Gamma-Glutamyl Transferase - GGT (nkat/L) | > 5.0 x ULN |
| Chemistry | Glucose (mmol/L) | < 40 mg/dL |
| Chemistry | Total Bilirubin (umol/L) | > 3.0 x ULN |
| Hematology | Absolute lymphocytes (10^9/L) | < 0.5 x 10^3/uL or > 20 x 10^3/uL |
| Hematology | Absolute neutrophils (10^9/L) | <1 x 10^3/uL |
| Hematology | Hemoglobin (g/L) | Increase of > 4 g/dL compared to Baseline |
| Hematology | Platelet count (10^9/L) | < 50 x 10^9/L |
| Hematology | White Blood Cell - WBC (10^9/L) | <2 x 10^9/L |

The following tests do not have a clinically significant threshold in CTCAE 5.0 so will only be reported in the relevant shift tables based on LLN and ULN:

- Chemistry: BUN (blood urea nitrogen) (mg/dL), Conjugated bilirubin (umol/L), Glomerular Filtration Rate - GFR (mL/min), Lactate Dehydrogenase - LDH (nkat/L), Unconjugated bilirubin (umol/L).
- Hematology: Absolute basophils (10^9/L), Absolute eosinophils (10^9/L), Absolute monocytes (10^9/L), Atypical lymphocytes (%), Basophiles (%), D-Dimer (ug/L), Eosinophils (%), Hematocrit (%), Lymphocytes (%), Monocytes (%), Neutrophils (%).

They will not be in the table summarizing clinically significant abnormalities.

#### 8.6 Physical Examination

The physical examination data will only be listed by subject for each visit, with results graded as normal or abnormal.

DocuSign Envelope ID: 8F061211-20AA-42BB-A5E5-CF9CAC78AF08

#### 8.7 Oxygen Saturation

Oxygen saturation is measured by pulse oximetry ($SpO_2$) and is taken at every planned visit. Abnormalities of saturation levels will be summarized by treatment arms. All values will be listed.

### 9 CHANGES FROM PROTOCOL

The original protocol specifies that scheduled visits are taken at Day 7 and Day 14, but this was subsequently amended to Day 8 and Day 15 visits as per protocol amendment 1.3. These SAP and table shells are written according to the new definition of scheduled visits.

DocuSign Envelope ID: 8F061211-20AA-42BB-A5E5-CF9CAC78AF08
